# Evaluation of the impact of COVID-19 pandemic on hospital admission related to common infections

**DOI:** 10.1101/2023.07.16.23292723

**Authors:** Ali Fahmi, Victoria Palin, Xiaomin Zhong, Ya-Ting Yang, Simon Watts, Darren M Ashcroft, Ben Goldacre, Brian Mackenna, Louis Fisher, Jon Massey, Amir Mehrkar, Seb Bacon, OpenSAFELY collaborative, Kieran Hand, Tjeerd Pieter van Staa

## Abstract

**Background:** Antimicrobial resistance (AMR) is a multifaceted global challenge, partly driven by inappropriate antibiotic prescribing. The COVID-19 pandemic impacted antibiotic prescribing for common bacterial infections. This highlights the need to examine risk of hospital admissions related to common infections, excluding COVID-19 infections during the pandemic.

**Methods:** With the approval of NHS England, we accessed electronic health records from The Phoenix Partnership (TPP) through OpenSAFELY platform. We included patients with primary care diagnosis of common infections, including lower respiratory tract infection (LRTI), upper respiratory tract infections (URTI), and lower urinary tract infection (UTI), from January 2019 to August 2022. We excluded patients with a COVID-19 record 90 days before to 30 days after the infection diagnosis. Using Cox proportional-hazard regression models, we predicted risk of infection-related hospital admission in 30 days follow-up period after the diagnosis.

**Results:** We found 12,745,165 infection diagnoses from January 2019 to August 2022. Of them, 80,395 (2.05%) cases were admitted to hospital in the follow-up period. Counts of hospital admission for infections dropped during COVID-19, e.g., LRTI from 3,950 in December 2019 to 520 in April 2020. Comparing those prescribed an antibiotic to those without, reduction in risk of hospital admission were largest with LRTI (adjusted odds ratio (OR) of 0.35; 95% CI, 0.35-0.36) and UTI (adjusted OR 0.45; 95% CI, 0.44-0.46), compared to URTI (adjusted OR 1.04; 95% CI, 1.03-1.06).

**Conclusion:** Large effectiveness of antibiotics in preventing complications related to LRTI and UTI can support better targeting of antibiotics to patients with higher complication risks.

**Key messages:**

- The main drivers of infection-related hospital admission are age, Charlson comorbidity index, and history of prior antibiotics.
- Antibiotics are more effective in preventing hospital admission related to infections such as lower respiratory tract infection and urinary tract infection, rather than upper respiratory tract infection.
- Common antibiotic types are associated with more reduction in the risk of infection-related hospital admission.

## Introduction

Antimicrobial resistance (AMR) is a multifaceted global challenge that needs to be managed through antimicrobial stewardship interventions.^1, 2^ Antibiotics are prescribed to prevent infections, but if prescribed inappropriately or excessively, antibiotics use can drive AMR.^3^ Prescribing of antibiotics declined between the end of 2019 and 2021 compared to previous years as an indirect impact of the COVID-19 pandemic,^2^ mainly due to reduced social mixing and spread of infections.

Few studies have evaluated the risk of hospital admissions related to common infections and antibiotic prescribing during the COVID-19 pandemic. During the pandemic, Zhu et al. found a reduction in community antibiotics prescribing in northwest London.^4^ Silva et al. evaluated the impact of the pandemic on the trend of antibiotics prescribing in outpatient care in Portugal and found a significant reduction in antibiotic prescribing in outpatient care.^5^ Several pre-pandemic studies have investigated the link between using antibiotics and developing complications, for example, Mistry et al. evaluated the risk of incident complication related to urinary tract infection (UTI), upper respiratory tract infection (URTI), and lower respiratory tract infection (LRTI).^3^ van Bodegraven et al. found an association between lower rates of antibiotics prescribing and higher risk of infection-related complications.^6^ Whilst these studies are informative, there is a need to understand the impact of the pandemic on outcomes after common infections. The aims of this study were to evaluate the impact of the COVID-19 pandemic on the primary care treatment with antibiotic for common infections in England and to develop and validate risk prediction models for infection-related complications. Risk prediction models are statistical models that aim to predict the probability of future events, such as whether a patient will develop a disease or not.

## Methods

In this cohort study, we used data from the OpenSAFELY platform (https://opensafely.org/) that focuses on urgent research into COVID-19 pandemic and securely links, pseudonymises, stores, and analyses EHR on behalf of the National Health Service pandemic.^7, 8^ This platform provides almost 24 million people’s pseudonymised patient-level data of primary care from The Phoenix Partnership (TPP). These data are linked to external databases such as the Hospital Episode Statistics (HES)^9^ and Office for National Statistics (ONS)^10^ using industry standard cryptographic hashing techniques.^11^ The OpenSAFELY data include patient-level demographics (age, sex, body mass index (BMI), ethnicity, and smoking status), clinical diagnoses history, medication history, and vaccination history. These data are linked to the hospital admission records through the Admitted Patient Care (APC) dataset.^12^

## Study population

This study included adult patients (18 years old or more) registered with a GP during study period (January 2019 to August 2022). We identified patients who were diagnosed with a common infection, namely LRTI, URTI (including specific URTI, cough, cold with cough, and sore throat), lower UTI (not including renal infections), sinusitis, otitis media, and otitis externa. We extracted 20 different infection diagnoses which created the cohorts for common infections. We investigated the records of hospital admission that occurred during the 30 days after the date of infection diagnosis (i.e., follow-up). To extract hospital admission records, we used the codes of the 10th revision of International Classification of Diseases (ICD10) for the primary admission diagnosis for a broad set of infection-related complications. These codes are available at opencodelists website.^13^ We excluded patients who had a record of COVID-19 diagnosis that occurred 90 days before or 30 days after the date of infection diagnosis. For COVID-19 records, we combined the records of the Second Generation Surveillance System (SGSS)^14^ and GP records of COVID-19 diagnosis in primary care. The study definition scripts for each common infection can be found in the hospitalisation branch of GitHub repository of the BRIT2 project.^15^ Figure 1 shows the study design.

**Figure 1.**
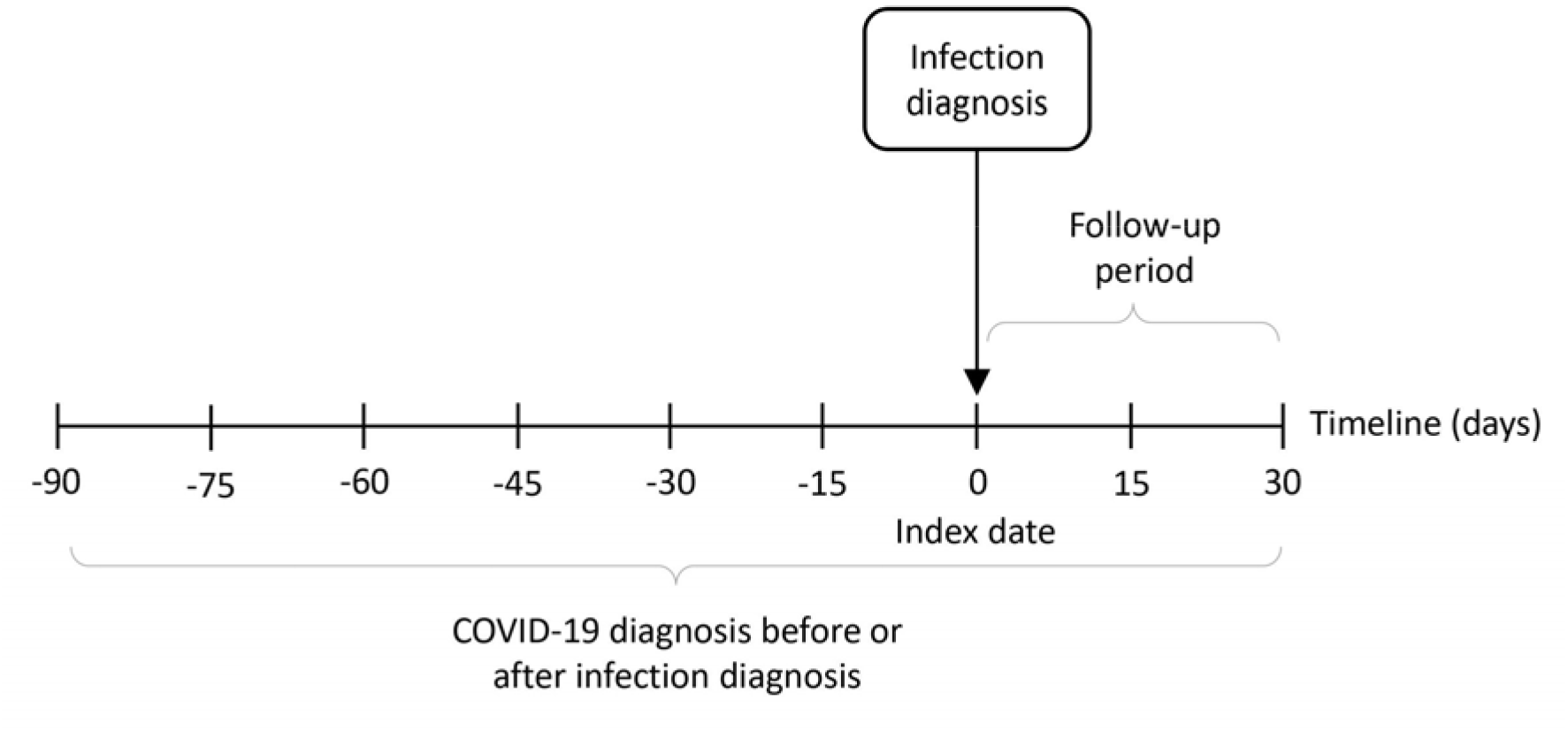
Diagram of study design.

## Variables

We extracted predictor variables that could potentially have an association with the risk of hospital admission related to common infections. These variables were age, sex, ethnicity, smoking status, socioeconomic class measured with the Index of Multiple Deprivation (IMD), region in England, BMI, comorbidities measured with the Charlson Comorbidity Index (CCI),^16^ season of infection diagnosis, flu vaccination in the one year before, and history of prior antibiotics measured by the count of antibiotic prescriptions in the one year before.

## Statistical methods

The common infection cohorts were split into four sub-datasets: 1) incident infection with no prescribed antibiotics, 2) incident infection with prescribed antibiotics, 3) prevalent infection with no prescribed antibiotics, and 4) prevalent infection with prescribed antibiotics. Incident common infections refer to having no diagnosis record for the same infection within 42 days prior to the diagnosis of common infection. Prevalent common infections were any infections that were not incident. Prescribed antibiotics were given in 5 days after the date of infection diagnosis. DMD codes were used to identify antibiotics.^17^ We analysed the count and rate of infection-related hospital admissions for each sub-dataset. To calculate rates, we divided the count of infection-related hospital admission cases (numerator) by the count of infection diagnosis (denominator). A Cox proportional-hazards regression model was fitted to each sub-dataset. Censored patients were those who died or deregistered from the practice after being hospitalised (within the follow-up period). Patients with a missing value for ethnicity, smoking status, IMD, and BMI variables were given a missing indicator called Unknown. We dropped patients who had no record for the region since they were very rare.

Each sub-dataset for each common infection was randomly split into development (75%) and validation (25%) cohorts and used to develop and validate a set of Cox models for common infections, namely LRTI, URTI, UTI, sinusitis, otitis media, and otitis externa. Similarly, a set of Cox models were developed and validated for the components of the URTI: specific URTI, cough, cold with cough, and sore throat. To assess the effect of prescribed antibiotics on hospital admission related to incident or prevalent common infections, we developed Cox models with an additional predictor binary variable for prescribed antibiotics given within 5 days after diagnosis. We also built Cox models with an additional categorical variable of antibiotic type (most prescribed, second most prescribed, others, and none) to estimate the effect of antibiotic type on hospital admission related to incident or prevalent common infections.

For evaluation of impact of the COVID-19 pandemic on antibiotic treatment of common infections, we analysed the count of infection-related hospital admission considering four periods regarding COVID-19 status: 1) pre-pandemic (January 2019 to December 2019), 2) beginning of the pandemic (January 2020 to April 2020), 3) during the pandemic (May 2020 to April 2021), and 4) after the second national lockdown (May 2021 to August 2022). We built Cox models with stratified data by age categories, sex categories, and period categories (pre-pandemic, beginning and during pandemic, and after second lockdown) to predict infection-related hospital admission. The models with stratified period categories can indicate the effect of antibiotics use on infection-related hospital admission in each period.

The performance of Cox models with overall data and pre-pandemic data was evaluated with concordance value or C-statistic, which measures the models’ ability to discriminate patients who developed complication and those who did not. Calibration of models was calculated by comparing the observed and predicted risks for deciles of predicted risks of infection-related hospital admission. The TRIPOD checklist was completed for the development of prediction models. The models and their performance analyses were done in Jupyter notebooks with Python 3.8.8.^18^ The notebooks are available in the hospitalisation branch of GitHub repository of BRIT2 project.^15^ The lifelines package version 0.26.4 was used to develop Cox models and validate them.^19^

## Results

A total of 12,745,165 diagnoses of common infections was found from January 2019 to August 2022, of which 11,455,025 (89.88%) were incident and 1,290,140 (10.12%) were prevalent. Of incident common infections, 7,539,015 (65.81%) were prescribed antibiotics (86.33% received antibiotics for incident LRTI, 55.93% for URTI, and 86.79% for UTI). Of prevalent common infection, 789,340 (61.18%) were prescribed antibiotics. Table 1 shows the main baseline characteristics of the cohort of incident common infections (namely LRTI, URTI, and UTI) without prescribed antibiotics. Supplementary Tables 1-4 display remaining baseline characteristics of these cohorts as well as those for other infections (sinusitis, otitis media, otitis externa, specific URTI, cough, cold with cough, and sore throat). In all cohorts, BMI, ethnicity, smoking status, and IMD had missing values of 19.81%, 34.76%, 0.50%, and 1.63%, respectively.

**Table 1.** Baseline characteristics of the cohorts of incident common infections in patients not prescribed antibiotics (using data from January 2019 to August 2022).

|  | LRTI | URTI | UTI |
| --- | --- | --- | --- |
| **Total, N infections** | 252,975 | 2,567,235 | 294,325 |
| **Age, N (%)** |  |  |  |
| 18-24 | 10,150 (4.01) | 158,300 (6.17) | 25,545 (8.68) |
| 25-34 | 20,185 (7.98) | 263,190 (10.25) | 35,195 (11.96) |
| 35-44 | 22,390 (8.85) | 267,840 (10.43) | 28,180 (9.57) |
| 45-54 | 31,215 (12.34) | 372,565 (14.51) | 33,920 (11.52) |
| 55-64 | 39,005 (15.42) | 480,850 (18.73) | 36,890 (12.53) |
| 65-74 | 50,195 (19.84) | 548,215 (21.35) | 48,330 (16.42) |
| 75+ | 79,830 (31.56) | 476,275 (18.55) | 86,270 (29.31) |
| **Sex, N (%)** |  |  |  |
| Male | 108,850 (43.03) | 1,116,735 (43.50) | 83,720 (28.45) |
| Female | 144,125 (56.97) | 1,450,500 (56.50) | 210,605 (71.55) |
| **Ethnicity, N (%)** |  |  |  |
| White | 145,930 (57.69) | 1,519,395 (59.18) | 168,765 (57.34) |
| Non-White | 15,905 (6.29) | 190,065 (7.40) | 19,250 (6.54) |
| Unknown | 91,135 (36.03) | 857,780 (33.41) | 106,310 (36.12) |
| **CCI<sup>1</sup>, N (%)** |  |  |  |
| Very low (=0) | 123,435 (48.79) | 1,499,380 (58.40) | 176,150 (59.85) |
| Low (=1) | 95,705 (37.83) | 846,450 (32.97) | 85,315 (28.99) |
| Medium (=2) | 26,145 (10.33) | 179,440 (6.99) | 25,080 (8.52) |
| High (=3 and =4) | 5,645 (2.23) | 32,810 (1.28) | 5,900 (2.00) |
| Very high (≥5) | 2,040 (0.81) | 9,160 (0.36) | 1,875 (0.64) |
| **Flu vaccination, N (%)** |  |  |  |
| Yes | 134,495 (53.17) | 1,237,915 (48.22) | 133,530 (45.37) |
| No | 118,475 (46.83) | 1,329,320 (51.78) | 160,800 (54.63) |
| **Period** |  |  |  |
| Pre-pandemic | 103,175 (40.78) | 990,915 (38.60) | 91,650 (31.14) |
| Beginning and during pandemic | 67,095 (26.52) | 723,000 (28.16) | 91,120 (30.96) |
| After 2<sup>nd</sup> lockdown | 82,705 (32.69) | 853,320 (33.24) | 111,555 (37.90) |
| **Count of antibiotic prescription in the one year before, mean (SD<sup>2</sup>)** | 2.20 (2.86) | 1.42 (2.09) | 2.30 (3.09) |

<sup>1</sup> CCI, Charlson Comorbidities Index, measured from 17 weighted conditions, including myocardial infarction, congestive heart failure, peripheral vascular disease, cerebrovascular disease, dementia, chronic pulmonary disease, Connective tissue disease, ulcer disease, mild liver disease, diabetes, hemiplegia, moderate or severe renal disease, diabetes with complications, any malignancy (including leukaemia and lymphoma), moderate or severe liver disease, metastatic solid tumour, and AIDS.
<sup>2</sup> SD, standard deviation.

Table 2 and Supplementary Tables 5-8 show the counts and rates of hospital admission related to incident common infections in 1,000 patients, from January 2019 to August 2022. We found 268,805 cases of infection-related hospital admission within 30-day follow-up after infection diagnosis. Rates were highest in patients with the highest CCI (rate of 149.5 in LRTI, 58.4 in URTI, and 168.0 in UTI). Although the counts of LRTI and URTI dropped during the pandemic, the rates did not change substantially by COVID-19 status (Table 2). Rates of UTI, however, fluctuated; from 66.8 pre-pandemic to 56.7 in the beginning and during the pandemic periods, and then to 61.4 after the second lockdown. Figure 2 shows a reduction of infection-related hospital admission in the beginning of the pandemic, between January 2020 and April 2020, especially in respiratory tract infections (LRTI and URTI) which dropped by 87% and 77%, respectively, between January 2020 and April 2020.

**Figure 2.**
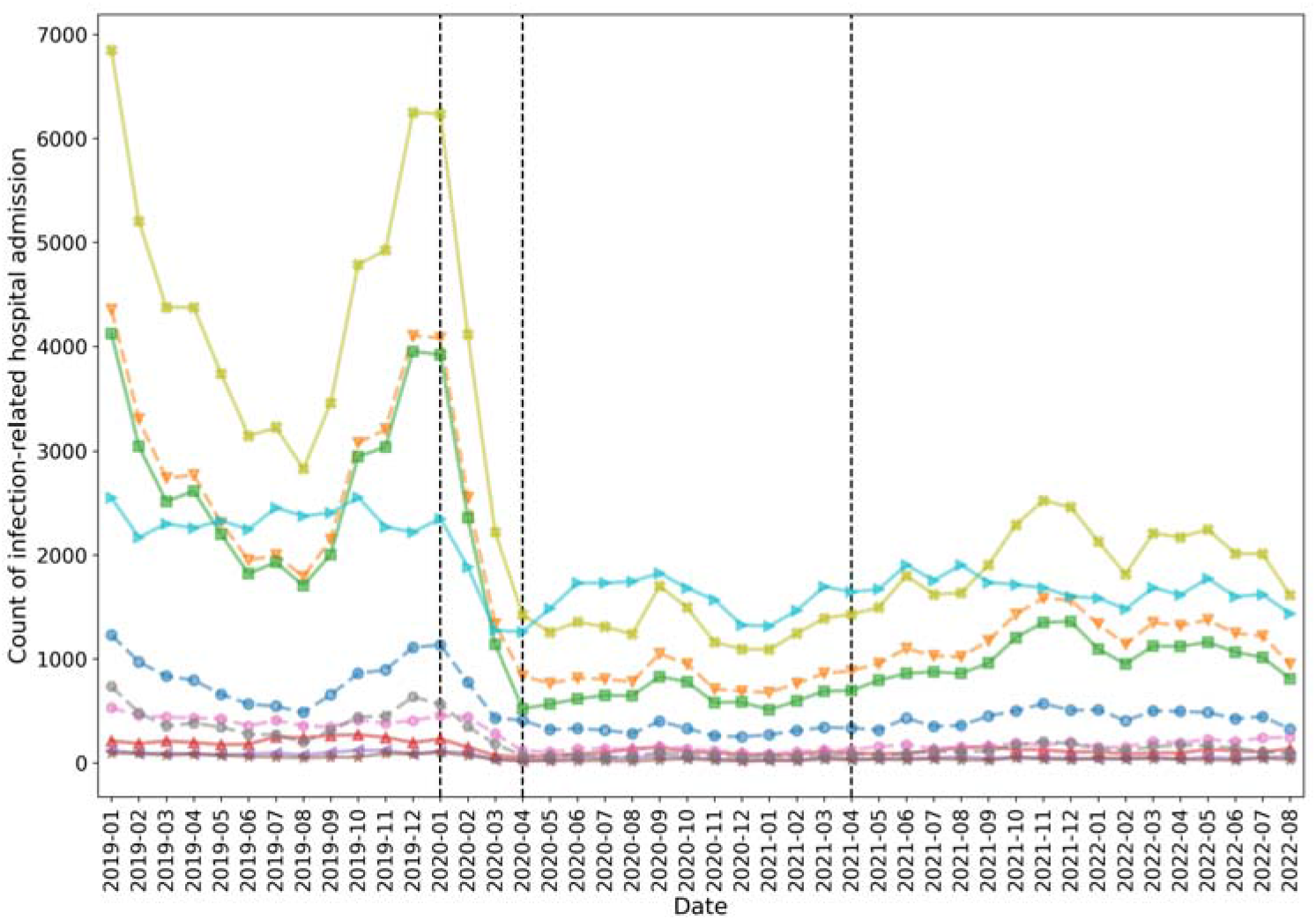
Count of infection-related hospital admission, where 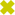 upper respiratory tract infections 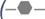 specific upper respiratory tract infection, 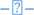 cough, 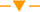 cold with cough, and 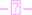 sore throat), 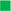 lower respiratory tract infection, 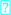 urinary tract infection, 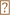 sinusitis, 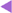 otitis media, and 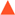 otitis externa.

**Table 2.** Count and rate of hospital admission related to incident common infections in patients not prescribed antibiotics (using data from January 2019 to August 2022).

|  | LRTI | URTI | UTI |
| --- | --- | --- | --- |
| **Total, N cases** | 17,915 | 40,035 | 18,140 |
| **Age, N (rate<sup>1</sup>)** |  |  |  |
| 18-24 | 235 (23.2) | 1,560 (9.9) | 455 (17.8) |
| 25-34 | 505 (25.0) | 2,150 (8.2) | 615 (17.5) |
| 35-44 | 670 (29.9) | 2,150 (8.0) | 565 (20.0) |
| 45-54 | 1,210 (38.8) | 3,180 (8.5) | 1,000 (29.5) |
| 55-64 | 2,030 (52.0) | 4,965 (10.3) | 1,545 (41.9) |
| 65-74 | 3,870 (77.1) | 8,495 (15.5) | 3,525 (72.9) |
| 75+ | 9,395 (117.7) | 17,540 (36.8) | 10,435 (121.0) |
| **Sex, N (rate<sup>1</sup>)** |  |  |  |
| Male | 8,690 (79.8) | 19,235 (17.2) | 8,160 (97.5) |
| Female | 9,225 (64.0) | 20,800 (14.3) | 9,980 (47.4) |
| **Ethnicity, N (rate<sup>1</sup>)** |  |  |  |
| White | 10,255 (70.3) | 22,810 (15.0) | 10,310 (61.1) |
| Non-White | 780 (49.0) | 2,120 (11.2) | 705 (36.6) |
| Unknown | 6,880 (75.5) | 15,105 (17.6) | 7,125 (67.0) |
| **CCI<sup>2</sup>, N (rate<sup>1</sup>)** |  |  |  |
| Very low (=0) | 6,520 (52.8) | 16,495 (11.0) | 7,210 (40.9) |
| Low (=1) | 7,400 (77.3) | 15,580 (18.4) | 6,790 (79.6) |
| Medium (=2) | 2,940 (112.4) | 5,845 (32.6) | 2,940 (117.2) |
| High (=3 and =4) | 750 (132.9) | 1,580 (48.2) | 880 (149.2) |
| Very high (≥5) | 305 (149.5) | 535 (58.4) | 315 (168.0) |
| **Flu vaccination, N (rate<sup>1</sup>)** |  |  |  |
| Yes | 11,765 (87.5) | 24,535 (19.8) | 11,660 (87.3) |
| No | 6,150 (51.9) | 15,500 (11.7) | 6,475 (40.3) |
| **Periods, N (rate<sup>1</sup>)** |  |  |  |
| Pre-pandemic | 7,375 (71.5) | 16,165 (16.3) | 6,125 (66.8) |
| Beginning and during pandemic | 4,575 (68.2) | 10,440 (14.4) | 5,170 (56.7) |
| After 2<sup>nd</sup> lockdown | 5,965 (72.1) | 13,430 (15.7) | 6,845 (61.4) |

<sup>1</sup> Rate is the number of cases per 1000 patients with common infection, calculated by dividing the count of infection-related hospital admission cases (numerator) by the count of infection diagnosis (denominator) and then multiplied by 1000
<sup>2</sup> CCI, Charlson Comorbidities Index, measured from 17 weighted conditions, including myocardial infarction, congestive heart failure, peripheral vascular disease, cerebrovascular disease, dementia, chronic pulmonary disease, Connective tissue disease, ulcer disease, mild liver disease, diabetes, hemiplegia, moderate or severe renal disease, diabetes with complications, any malignancy (including leukaemia and lymphoma), moderate or severe liver disease, metastatic solid tumour, and AIDS

C-statistics of the Cox models with development and validation datasets of incident infections with no antibiotics were respectively 0.68 and 0.67 for LRTI, 0.73 and 0.72 for URTI, and 0.73 and 0.73 for UTI. Supplementary Table 9 presents C-statistics of the Cox models with development and validation datasets for hospital admission related to all incident and prevalent infections with and without antibiotics. Infections included LRTI, URTI, UTI, sinusitis, otitis media, otitis externa, as well as the components of the URTI: URTI, cough, cold with cough, and sore throat, with and without antibiotics.

As presented in Table 3, age was strongly associated with infection-related complication with HRs of age category 75+ being 5.27 (95% CI 4.49 to 6.28) in LRTI, 3.54 (95% CI 3.31 to 3.78) in URTI, and 5.54 (95% CI 4.94 to 6.20) in UTI. CCI also had a high association with hospital admission related to incident infections as HRs of very high CCI were 2.03 (95% CI 1.78 to 2.31) for LRTI, 3.18 (95% CI 2.88 to 3.51) for URTI, and 2.21 (95% CI 1.94 to 2.52) for UTI. History of prior antibiotics influenced the risk of infection-related complication, especially in mild infections like URTI with HR of 1.14 (95% CI 1.13-1.14). Supplementary Tables 10-12 show HRs for these predictors for other infections.

**Table 3.**
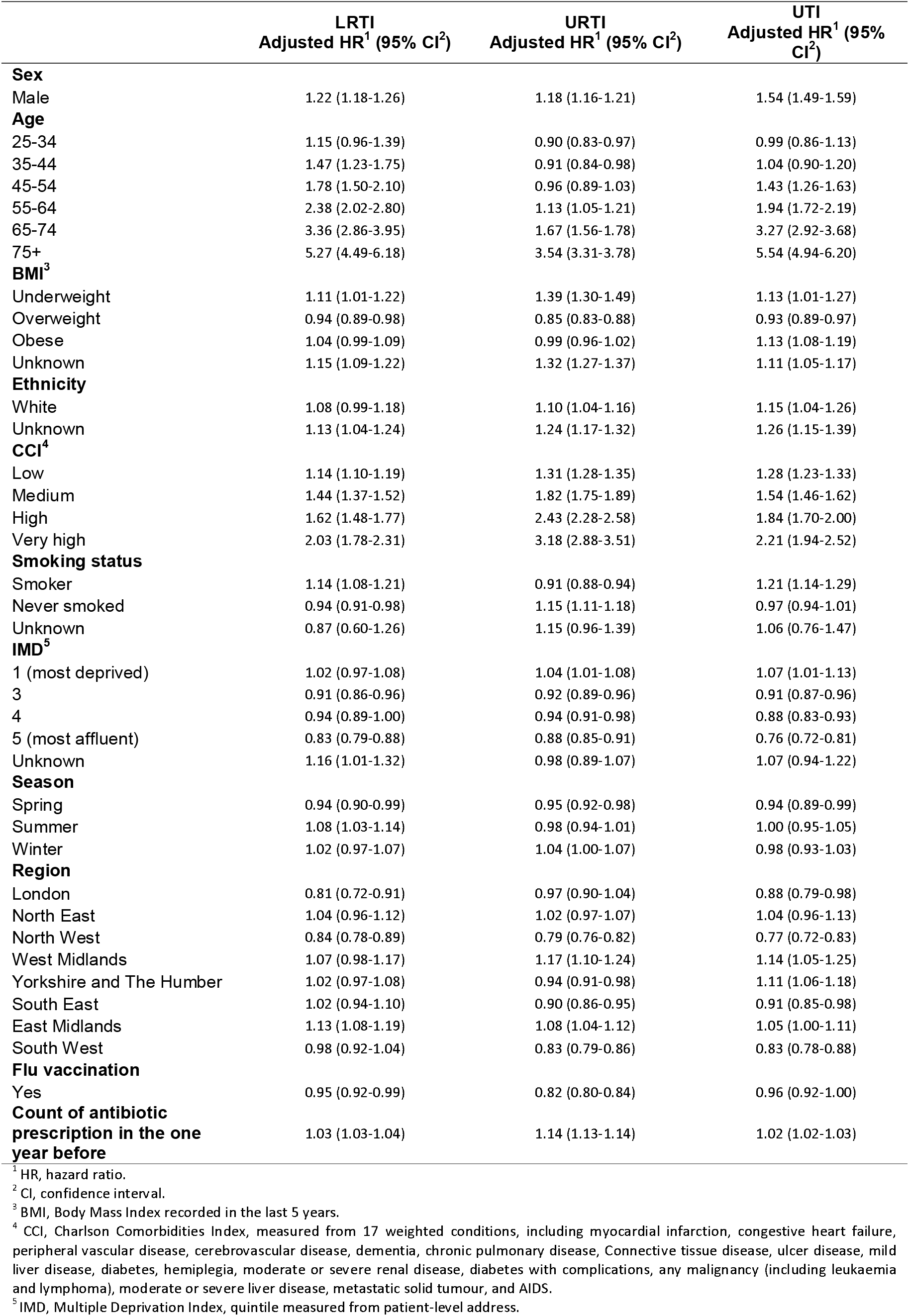
Adjusted hazard ratios of infection-related hospital admissions for several predictors for incident common infection stratified by infection (using data from January 2019 to August 2022)

|  | LRTI<br>Adjusted HR<sup>1</sup> (95% CI<sup>2</sup>) | URTI<br>Adjusted HR<sup>1</sup> (95% CI<sup>2</sup>) | UTI<br>Adjusted HR<sup>1</sup> (95% CI<sup>2</sup>) |
| --- | --- | --- | --- |
| **Sex** |  |  |  |
| Male | 1.22 (1.18-1.26) | 1.18 (1.16-1.21) | 1.54 (1.49-1.59) |
| **Age** |  |  |  |
| 25-34 | 1.15 (0.96-1.39) | 0.90 (0.83-0.97) | 0.99 (0.86-1.13) |
| 35-44 | 1.47 (1.23-1.75) | 0.91 (0.84-0.98) | 1.04 (0.90-1.20) |
| 45-54 | 1.78 (1.50-2.10) | 0.96 (0.89-1.03) | 1.43 (1.26-1.63) |
| 55-64 | 2.38 (2.02-2.80) | 1.13 (1.05-1.21) | 1.94 (1.72-2.19) |
| 65-74 | 3.36 (2.86-3.95) | 1.67 (1.56-1.78) | 3.27 (2.92-3.68) |
| 75+ | 5.27 (4.49-6.18) | 3.54 (3.31-3.78) | 5.54 (4.94-6.20) |
| **BMI<sup>3</sup>** |  |  |  |
| Underweight | 1.11 (1.01-1.22) | 1.39 (1.30-1.49) | 1.13 (1.01-1.27) |
| Overweight | 0.94 (0.89-0.98) | 0.85 (0.83-0.88) | 0.93 (0.89-0.97) |
| Obese | 1.04 (0.99-1.09) | 0.99 (0.96-1.02) | 1.13 (1.08-1.19) |
| Unknown | 1.15 (1.09-1.22) | 1.32 (1.27-1.37) | 1.11 (1.05-1.17) |
| **Ethnicity** |  |  |  |
| White | 1.08 (0.99-1.18) | 1.10 (1.04-1.16) | 1.15 (1.04-1.26) |
| Unknown | 1.13 (1.04-1.24) | 1.24 (1.17-1.32) | 1.26 (1.15-1.39) |
| **CCI<sup>4</sup>** |  |  |  |
| Low | 1.14 (1.10-1.19) | 1.31 (1.28-1.35) | 1.28 (1.23-1.33) |
| Medium | 1.44 (1.37-1.52) | 1.82 (1.75-1.89) | 1.54 (1.46-1.62) |
| High | 1.62 (1.48-1.77) | 2.43 (2.28-2.58) | 1.84 (1.70-2.00) |
| Very high | 2.03 (1.78-2.31) | 3.18 (2.88-3.51) | 2.21 (1.94-2.52) |
| **Smoking status** |  |  |  |
| Smoker | 1.14 (1.08-1.21) | 0.91 (0.88-0.94) | 1.21 (1.14-1.29) |
| Never smoked | 0.94 (0.91-0.98) | 1.15 (1.11-1.18) | 0.97 (0.94-1.01) |
| Unknown | 0.87 (0.60-1.26) | 1.15 (0.96-1.39) | 1.06 (0.76-1.47) |
| **IMD<sup>5</sup>** |  |  |  |
| 1 (most deprived) | 1.02 (0.97-1.08) | 1.04 (1.01-1.08) | 1.07 (1.01-1.13) |
| 3 | 0.91 (0.86-0.96) | 0.92 (0.89-0.96) | 0.91 (0.87-0.96) |
| 4 | 0.94 (0.89-1.00) | 0.94 (0.91-0.98) | 0.88 (0.83-0.93) |
| 5 (most affluent) | 0.83 (0.79-0.88) | 0.88 (0.85-0.91) | 0.76 (0.72-0.81) |
| Unknown | 1.16 (1.01-1.32) | 0.98 (0.89-1.07) | 1.07 (0.94-1.22) |
| **Season** |  |  |  |
| Spring | 0.94 (0.90-0.99) | 0.95 (0.92-0.98) | 0.94 (0.89-0.99) |
| Summer | 1.08 (1.03-1.14) | 0.98 (0.94-1.01) | 1.00 (0.95-1.05) |
| Winter | 1.02 (0.97-1.07) | 1.04 (1.00-1.07) | 0.98 (0.93-1.03) |
| **Region** |  |  |  |
| London | 0.81 (0.72-0.91) | 0.97 (0.90-1.04) | 0.88 (0.79-0.98) |
| North East | 1.04 (0.96-1.12) | 1.02 (0.97-1.07) | 1.04 (0.96-1.13) |
| North West | 0.84 (0.78-0.89) | 0.79 (0.76-0.82) | 0.77 (0.72-0.83) |
| West Midlands | 1.07 (0.98-1.17) | 1.17 (1.10-1.24) | 1.14 (1.05-1.25) |
| Yorkshire and The Humber | 1.02 (0.97-1.08) | 0.94 (0.91-0.98) | 1.11 (1.06-1.18) |
| South East | 1.02 (0.94-1.10) | 0.90 (0.86-0.95) | 0.91 (0.85-0.98) |
| East Midlands | 1.13 (1.08-1.19) | 1.08 (1.04-1.12) | 1.05 (1.00-1.11) |
| South West | 0.98 (0.92-1.04) | 0.83 (0.79-0.86) | 0.83 (0.78-0.88) |
| **Flu vaccination** |  |  |  |
| Yes | 0.95 (0.92-0.99) | 0.82 (0.80-0.84) | 0.96 (0.92-1.00) |
| **Count of antibiotic prescription in the one year before** | 1.03 (1.03-1.04) | 1.14 (1.13-1.14) | 1.02 (1.02-1.03) |

<sup>1</sup> HR, hazard ratio.
<sup>2</sup> CI, confidence interval.
<sup>3</sup> BMI, Body Mass Index recorded in the last 5 years.
<sup>4</sup> CCI, Charlson Comorbidities Index, measured from 17 weighted conditions, including myocardial infarction, congestive heart failure, peripheral vascular disease, cerebrovascular disease, dementia, chronic pulmonary disease, Connective tissue disease, ulcer disease, mild liver disease, diabetes, hemiplegia, moderate or severe renal disease, diabetes with complications, any malignancy (including leukaemia and lymphoma), moderate or severe liver disease, metastatic solid tumour, and AIDS.
<sup>5</sup> IMD, Multiple Deprivation Index, quintile measured from patient-level address.

HRs of antibiotic exposure (compared to non-exposure) for incident infections were 0.35 (95% CI 0.35 to 0.36) for LRTI, 1.04 (95% CI 1.03 to 1.06) for URTI, and 0.45 (95% CI 0.44 to 0.46) for UTI (Table 4). Patients prescribed the most prescribed antibiotic type had comparable risks to those prescribed the second most prescribed type, e.g., for incident LRTI 1.02 (95% CI 0.99-1.05). However, patients prescribed less frequent types had increased risks of infection-related hospital admission, e.g., HR of 1.72 (95% CI 1.67-1.78) for incident LRTI. No major effect modification in the HRs were observed when stratifying by sex, age, and period regarding COVID-19 status. Table 4 and Supplementary Tables 13 and 14 show HRs of antibiotic exposure and antibiotic types for incident and prevalent common infections and HRs of models with stratifications by sex categories, age categories, and period.

**Table 4.** Crude hazard ratios of antibiotic exposure stratified by incident and prevalent common infection (using data from January 2019 to August 2022)

|  | LRTI |  | URTI |  | UTI |  |
| --- | --- | --- | --- | --- | --- | --- |
|  | Crude HR<sup>1</sup> (95% CI<sup>2</sup>) |  | Crude HR<sup>1</sup> (95% CI<sup>2</sup>) |  | Crude HR<sup>1</sup> (95% CI<sup>2</sup>) |  |
|  | Incident | Prevalent | Incident | Prevalent | Incident | Prevalent |
| **Antibiotic exposure** |  |  |  |  |  |  |
| No exposure | reference | reference | reference | reference | reference | reference |
| Exposed | 0.35 (0.35-0.36) | 0.51 (0.48-0.53) | 1.04 (1.03-1.06) | 0.87 (0.84-0.91) | 0.45 (0.44-0.46) | 0.53 (0.51-0.55) |
| **Antibiotic type<sup>3</sup>** |  |  |  |  |  |  |
| Most prescribed | reference | reference | reference | reference | reference | reference |
| Second most prescribed | 1.02 (0.99-1.05) | 0.90 (0.83-0.97) | 1.00 (0.98-1.03) | 0.94 (0.88-1.00) | 1.15 (1.12-1.18) | 0.93 (0.88-1.00) |
| Others | 1.72 (1.67-1.78) | 1.42 (1.32-1.53) | 1.43 (1.40-1.47) | 1.36 (1.28-1.44) | 1.40 (1.36-1.43) | 1.14 (1.08-1.20) |
| None | 3.16 (3.09-3.24) | 2.17 (2.02-2.32) | 1.05 (1.03-1.07) | 1.26 (1.19-1.33) | 2.49 (2.43-2.55) | 1.96 (1.87-2.06) |
| **Stratified by sex** |  |  |  |  |  |  |
| Male | 0.34 (0.33-0.35) | 0.50 (0.47-0.54) | 1.07 (1.05-1.10) | 0.91 (0.86-0.96) | 0.54 (0.52-0.55) | 0.58 (0.54-0.61) |
| Female | 0.36 (0.35-0.37) | 0.51 (0.48-0.54) | 1.00 (0.98-1.03) | 0.86 (0.82-0.90) | 0.39 (0.38-0.40) | 0.51 (0.48-0.53) |
| **Stratified by age** |  |  |  |  |  |  |
| 18-24 | 0.32 (0.26-0.38) | 0.37 (0.24-0.57) | 0.88 (0.82-0.95) | 0.70 (0.59-0.83) | 0.37 (0.33-0.43) | 0.60 (0.45-0.80) |
| 25-34 | 0.32 (0.28-0.36) | 0.55 (0.42-0.73) | 0.86 (0.81-0.92) | 0.68 (0.58-0.79) | 0.43 (0.38-0.48) | 0.45 (0.36-0.56) |
| 35-44 | 0.26 (0.23-0.29) | 0.46 (0.36-0.58) | 0.79 (0.74-0.84) | 0.85 (0.71-1.01) | 0.39 (0.35-0.43) | 0.57 (0.46-0.72) |
| 45-54 | 0.26 (0.24-0.28) | 0.47 (0.39-0.56) | 0.93 (0.88-0.98) | 0.81 (0.71-0.92) | 0.37 (0.34-0.40) | 0.43 (0.36-0.51) |
| 55-64 | 0.27 (0.25-0.28) | 0.49 (0.43-0.56) | 1.02 (0.97-1.06) | 0.93 (0.83-1.03) | 0.44 (0.41-0.47) | 0.49 (0.43-0.56) |
| 65-74 | 0.31 (0.30-0.32) | 0.41 (0.37-0.46) | 1.08 (1.05-1.12) | 0.88 (0.81-0.95) | 0.41 (0.39-0.43) | 0.50 (0.46-0.54) |
| 75+ | 0.42 (0.41-0.43) | 0.55 (0.51-0.58) | 1.08 (1.05-1.10) | 0.89 (0.84-0.94) | 0.48 (0.47-0.50) | 0.57 (0.54-0.60) |
| **Stratified by time** |  |  |  |  |  |  |
| Pre-pandemic | 0.38 (0.36-0.39) | 0.55 (0.52-0.59) | 1.05 (1.03-1.08) | 0.88 (0.83-0.92) | 0.44 (0.42-0.45) | 0.54 (0.51-0.58) |
| Beginning and during pandemic | 0.40 (0.37-0.42) | 0.49 (0.43-0.55) | 1.16 (1.11-1.20) | 1.04 (0.95-1.15) | 0.40 (0.38-0.41) | 0.50 (0.46-0.54) |
| After 2<sup>nd</sup> lockdown | 0.37 (0.36-0.38) | 0.52 (0.49-0.55) | 1.10 (1.08-1.12) | 0.90 (0.86-0.94) | 0.41 (0.40-0.42) | 0.52 (0.49-0.54) |

<sup>1</sup> HR, hazard ratio.
<sup>2</sup> CI, confidence interval.
<sup>3</sup> The most prescribed and the second most prescribed type of antibiotic are, respectively, amoxicillin and doxycycline for LRTI and URTI, and nitrofurantoin and trimethoprim for UTI.

For further evaluation of impact of COVID-19 pandemic, we used the pre-pandemic data to develop and validate Cox models for hospital admission related to common infections (see supporting materials for more information).

## Discussion

The risk prediction models indicated that the main drivers of infection-related hospital admission are age, CCI, and history of prior antibiotics. Antibiotics were found to be more effective in preventing complications (compared to no treatment) related to infections such as LRTI and UTI, in contrast to URTI. The models demonstrated that common antibiotic types were associated with more reduction in the risk of infection-related hospital admission, whereas less common antibiotic types were associated with less reduction or increase in the risk of infection-related hospital admission.

Prescribed antibiotics were strongly associated with reduced risk of hospital admission related to incident LRTI and UTI, but not in URTI. We could not find much literature on the effectiveness of antibiotics in reducing the risk of hospital admission with common infections. There may be different possible explanations for our finding of greater antibiotic effectiveness in preventing hospital admission related to LRTI and UTI. The first one may be that viral infections may be more frequent with infections such as URTI. General practice in the UK does not routinely test whether common infections are bacterial or viral. The second explanation may be due to differential confounding, where antibiotics are prescribed to sicker patients in infections like URTI and healthier patients in infections like LRTI and UTI. However, we did not find substantive evidence for this and a study in a different database found that antibiotic prescribing was unrelated to patient’s risk of hospital admission for infection-related complications.^3^ The finding in this study of reduced antibiotic effects in preventing complications with increasing count of antibiotic prescription in the one year before is consistent with a previous study.^20^

We also found that antibiotics prescribed for infections like LRTI and UTI are more effective in preventing complications than infections like URTI. Our findings showed the most common antibiotic types for respiratory infections (e.g., LRTI or cough) were amoxicillin and doxycycline and for UTI were nitrofurantoin and trimethoprim, which correspond with the recommendations of the National Institute for Health and Care Excellence (NICE) guidelines for management of respiratory infections and UTI.^21–23^ Most common antibiotic types for each infection were associated with more reduction of infection-related complication risk, compared with less common antibiotic types. However, other types of antibiotics were less effective in general, although they were prescribed frequently, even more frequent than the second most common antibiotic type.

Infection-related hospital admissions reduced in the beginning and fluctuated during the pandemic, which can be interpreted as the consequence of less transmission of infections and lower diagnosis of common infections. Lower diagnosis can lead to lower prescribing of antibiotics for common infections. The latest report on antibiotics utilisation in England stated 10.9% decline in antibiotics consumption between 2019 to 2020, which is an evident impact of COVID-19 pandemic.^2^ An analysis of prescribing of first-line antibiotics in English primary care obtained a 13.5% reduction between March and September 2020 compared with March and September 2019.^24^ A qualitative interview study with GPs in England found that although GPs were more likely to prescribe empirical antibiotics for respiratory tract infections, they prescribed less antibiotics during the pandemic, except for UTI and skin infections,^25^ which is similar to our findings. International studies reported a similar decrease in antibiotics prescriptions, such as in Spain,^26^ Belgium,^27^ Brazil,^28^ and Netherlands,^29^ in the beginning of the pandemic. The reduction of antibiotics used could have unintended consequences such as severe infections and complications,^4^ which we tried to address in this study.

There are only a limited number of risk prediction models for the prognosis of common infections. Before the COVID-19 pandemic, risk prediction models were developed to predict complications related to incident LRTI, URTI, and UTI in UK primary care.^3^ This study’s HRs for infection-related hospital admissions are similar to our findings; greater age and CCI increase the risk of complications, but some variables such as white ethnicity or winter season have a different association in the current study. Another similar study investigated antibiotics prescribing for common infections (LRTI, URTI, UTI, sinusitis, otitis media, and otitis externa) in UK primary care, and found age, sex, region, and CCI as associating factors with prescribing antibiotics.^30^ Unlike these two studies and our study, others focused on risk prediction models for a specific infection, such as UTI,^31, 32^ sepsis,^33^ or pneumonia,^34^ and specific resistance strains, such as carbapenem-resistant Enterobacterales.^35, 36^

There are several strength and limitations of this study. The main strength of this study is that it only considered complications related to common infections in patients without COVID-19 that is conducted after the outbreak of COVID-19. Another strength of this study is that it employed a large national EHR dataset which made it possible to develop multiple prediction models for each common infection depending on the incidence or prevalence and whether antibiotics were prescribed or not. This is the first study to look at infection-related hospital admission following prevalent common infections in primary care. This is of importance since there are no guidelines in England for repeated infection, except for UTI. Models for hospital admissions related to prevalent common infections target patients who may be at higher risk of complication. Previous studies (such as Mistry et al.^3^) did not have this level of prediction models for different types of infection-related hospital admission (incident or prevalent). One weakness of our models that were developed with the overall data (January 2019 to August 2022) is that their accuracy can be challenged since they were developed with the fluctuating records of infections in primary care during the pandemic. Therefore, we developed and validated models with pre-pandemic data. This also highlights the need for updating risk prediction models for infection-related hospital admission in future, especially those that are or will be applied in clinical practice. A further limitation was the identification and exclusion of patients with COVID-19 and the uncertainty whether all COVID-19 tests performed by private companies were included in the SGSS. Records of infection diagnosis were based on GP consultations, either in person or virtual, which the latter became common during the COVID-19 pandemic and could potentially impact infection diagnosis remotely. There were also computational limits that constrained us in imputing missing values through imputation algorithms. The risk prediction models also did not include severity of the infections as signs and symptoms are generally not well coded by GPs. Confounding by severity of infection also was not controlled for in our analyses.

## Conclusion

The COVID-19 pandemic indirectly impacted the antibiotic treatment for common infections, particularly infections such as LRTI. Risk models found that age, CCI, and history of prior antibiotics were the main predictors of infection-related hospital admission. Prescribed antibiotics were strongly associated with lower risk of complications related to infections like LRTI and UTI, but not URTI. In order to improve risk-based antibiotic prescribing in primary care, there is a need for GPs and patients to be provided during consultation with personalised risks on the prognosis with a common infection.

## Data Availability

All data were linked, stored and analysed securely within the OpenSAFELY platform https://opensafely.org/. Data include pseudonymized data such as coded diagnoses, medications and physiological parameters. No free text data are included. All code is shared openly for review and re-use under MIT open license (https://github.com/opensafely/amr-uom-brit/). Detailed pseudonymised patient data is potentially re-identifiable and therefore not shared. We rapidly delivered the OpenSAFELY data analysis platform without prior funding to deliver timely analyses on urgent research questions in the context of the global COVID-19 health emergency: now that the platform is established, we are developing a formal process for external users to request access in collaboration with NHS England; details of this process will be published shortly on OpenSAFELY.org.

## Funding

This work was supported by Health Data Research UK (Better prescribing in frail elderly people with polypharmacy: learning from practice and nudging prescribers into better practice-BetterRx) and by National Institute for Health and Care Research (NIHR130581 - Cluster randomised trial to improve antibiotic prescribing in primary care: individualised knowledge support during consultation for general practitioners and patients – BRIT2). DMA is funded by the NIHR Greater Manchester Patient Safety Translational Research Centre (PSTRC-2016-003). The views expressed are those of the authors and not necessarily those of Health Data Research UK, the NHS, the NIHR, the Department of Health and Social Care or Public Health England.

## Acknowledgement

We are very grateful for all the support received from the TPP Technical Operations team throughout this work, and for generous assistance from the information governance and database teams at NHS England and the NHS England Transformation Directorate.

## Information governance and ethical approval

NHS England is the data controller for OpenSAFELY-TPP; TPP is the data processor; all study authors using OpenSAFELY have the approval of NHS England.^37^ This implementation of OpenSAFELY is hosted within the TPP environment which is accredited to the ISO 27001 information security standard and is NHS IG Toolkit compliant.^38^

Patient data has been pseudonymised for analysis and linkage using industry standard cryptographic hashing techniques; all pseudonymised datasets transmitted for linkage onto OpenSAFELY are encrypted; access to the platform is via a virtual private network (VPN) connection, restricted to a small group of researchers; the researchers hold contracts with NHS England and only access the platform to initiate database queries and statistical models; all database activity is logged; only aggregate statistical outputs leave the platform environment following best practice for anonymisation of results such as statistical disclosure control for low cell counts.^39^

The service adheres to the obligations of the UK General Data Protection Regulation (UK GDPR) and the Data Protection Act 2018. The service previously operated under notices initially issued in February 2020 by the Secretary of State under Regulation 3(4) of the Health Service (Control of Patient Information) Regulations 2002 (COPI Regulations), which required organisations to process confidential patient information for COVID-19 purposes; this set aside the requirement for patient consent.^40^ As of 1 July 2023, the Secretary of State has requested that NHS England continue to operate the Service under the COVID-19 Directions 2020.^41^ In some cases of data sharing, the common law duty of confidence is met using, for example, patient consent or support from the Health Research Authority Confidentiality Advisory Group.^42^

Taken together, these provide the legal bases to link patient datasets on the OpenSAFELY platform. GP practices, from which the primary care data are obtained, are required to share relevant health information to support the public health response to the pandemic, and have been informed of the OpenSAFELY analytics platform. This study was approved by the Health Research Authority and NHS Research Ethics Committee [REC reference 21/SC/0287].

## Data access and verification

Access to the underlying identifiable and potentially re-identifiable pseudonymised electronic health record data is tightly governed by various legislative and regulatory frameworks and restricted by best practice. The data in OpenSAFELY is drawn from General Practice data across England where TPP is the Data Processor. TPP developers initiate an automated process to create pseudonymised records in the core OpenSAFELY database, which are copies of key structured data tables in the identifiable records. These are linked onto key external data resources that have also been pseudonymised via SHA-512 one-way hashing of NHS numbers using a shared salt. DataLab developers and PIs holding contracts with NHS England have access to the OpenSAFELY pseudonymised data tables as needed to develop the OpenSAFELY tools. These tools in turn enable researchers with OpenSAFELY Data Access Agreements to write and execute code for data management and data analysis without direct access to the underlying raw pseudonymised patient data, and to review the outputs of this code. All code for the full data management pipeline—from raw data to completed results for this analysis—and for the OpenSAFELY platform as a whole is available for review at github.com/OpenSAFELY.

## Contribution

All authors contributed to and approved the final manuscript. Design of study was done by TvS, VP, and AF. Analysis was done by AF. Interpretation of results and first draft of manuscript: AF and TvS. The corresponding author attests that all listed authors meet authorship criteria and that no others meeting the criteria have been omitted.

## Conflict of Interest

All authors declare the following: BG and OpenSAFELY has received research funding from the Laura and John Arnold Foundation, the NHS National Institute for Health Research (NIHR), the NIHR School of Primary Care Research, NHS England, the NIHR Oxford Biomedical Research Centre, the Mohn-Westlake Foundation, NIHR Applied Research Collaboration Oxford and Thames Valley, the Wellcome Trust, the Good Thinking Foundation, Health Data Research UK, the Health Foundation, the World Health Organisation, UKRI MRC, Asthma UK, the British Lung Foundation, and the Longitudinal Health and Wellbeing strand of the National Core Studies programme; he is a Non-Executive Director at NHS Digital; he also receives personal income from speaking and writing for lay audiences on the misuse of science. AM has received consultancy fees (from https://inductionhealthcare.com) and is member of RCGP health informatics group and the NHS Digital GP data Professional Advisory Group that advises on access to GP Data for Pandemic Planning and Research (GDPPR). For the latter, he received payment for the GDPPR role.

## Notes

### Funding Statement

This study was funded by Health Data Research UK (Better prescribing in frail elderly people with polypharmacy: learning from practice and nudging prescribers into better practice BetterRx) and by National Institute for Health and Care Research.

### Author Declarations

NHS England is the data controller for OpenSAFELY-TPP; TPP is the data processor; all study authors using OpenSAFELY have the approval of NHS England. This implementation of OpenSAFELY is hosted within the TPP environment which is accredited to the ISO 27001 information security standard and is NHS IG Toolkit compliant. Patient data has been pseudonymised for analysis and linkage using industry standard cryptographic hashing techniques; all pseudonymised datasets transmitted for linkage onto OpenSAFELY are encrypted; access to the platform is via a virtual private network (VPN) connection, restricted to a small group of researchers; the researchers hold contracts with NHS England and only access the platform to initiate database queries and statistical models; all database activity is logged; only aggregate statistical outputs leave the platform environment following best practice for anonymisation of results such as statistical disclosure control for low cell counts. The service adheres to the obligations of the UK General Data Protection Regulation (UK GDPR) and the Data Protection Act 2018. The service previously operated under notices initially issued in February 2020 by the Secretary of State under Regulation 3(4) of the Health Service (Control of Patient Information) Regulations 2002 (COPI Regulations), which required organisations to process confidential patient information for COVID-19 purposes; this set aside the requirement for patient consent. As of 1 July 2023, the Secretary of State has requested that NHS England continue to operate the Service under the COVID-19 Directions 2020. In some cases of data sharing, the common law duty of confidence is met using, for example, patient consent or support from the Health Research Authority Confidentiality Advisory Group. Taken together, these provide the legal bases to link patient datasets on the OpenSAFELY platform. GP practices, from which the primary care data are obtained, are required to share relevant health information to support the public health response to the pandemic, and have been informed of the OpenSAFELY analytics platform. This study was approved by the Health Research Authority and NHS Research Ethics Committee [REC reference 21/SC/0287].

## References

1. Sugden R, Kelly R, Davies S. Combatting antimicrobial resistance globally. Nat Microbiol. Macmillan Publishers Limited; 2016;1(10).

2. Ashiru-Oredope D, Hopkins S, ESPAUR Oversight Group Writing Committee. English surveillance programme for antimicrobial utilisation and resistance (ESPAUR): Report 2020 to 2021. Public Heal Engl Publ [Internet]. 2021;1–209. Available from: https://assets.publishing.service.gov.uk/government/uploads/system/uploads/attachment_data/file/1069632/espaur-report-2020-to-2021-16-Nov-FINAL-v2.pdf

3. Mistry C, Palin V, Li Y, et al. Development and validation of a multivariable prediction model for infection-related complications in patients with common infections in UK primary care and the extent of risk-based prescribing of antibiotics. BMC Med. BMC Medicine; 2020;18(1):1–13.

4. Zhu N, Aylin P, Rawson T, Gilchrist M, Majeed A, Holmes A. Investigating the impact of COVID-19 on primary care antibiotic prescribing in North West London across two epidemic waves. Clin Microbiol Infect [Internet]. Elsevier Ltd; 2021;27(5):762–768. Available from: https://doi.org/10.1016/j.cmi.2021.02.007

5. Silva TM, Estrela M, Gomes ER, et al. The impact of the covid-19 pandemic on antibiotic prescribing trends in outpatient care: A nationwide, quasi-experimental approach. Antibiotics. 2021;10(9).

6. Bodegraven B van, Palin V, Mistry C, et al. Infection-related complications after common infection in association with new antibiotic prescribing in primary care: Retrospective cohort study using linked electronic health records. BMJ Open. 2021;11(1):1–11.

7. NHS. Opensafely: secure access to data to deepen our understanding of covid-19 [Internet]. [cited 2022 Aug 1]. Available from: https://transform.england.nhs.uk/key-tools-and-info/data-saves-lives/improving-care-through-research-and-innovation/opensafely-secure-access-to-data-to-deepen-our-understanding-of-covid-19/#:~:text=untilfurthernotice.-,OpenSAFELY%3Asecureaccesstodat

8. About OpenSAFELY [Internet]. [cited 2022 Jan 8]. Available from: https://www.opensafely.org/about/

9. OpenSAFELY. Hospital admissions [Internet]. [cited 2023 Jun 15]. Available from: https://docs.opensafely.org/data-sources/apc/

10. OpenSAFELY. Registered deaths [Internet]. [cited 2023 Jun 15]. Available from: https://docs.opensafely.org/data-sources/onsdeaths/

11. OpenSAFELY. Policies for Researchers [Internet]. 2023 [cited 2023 May 23]. Available from: https://www.opensafely.org/policies-for-researchers/

12. OpenSAFELY. Hospital admissions [Internet]. [cited 2022 Jan 11]. Available from: https://docs.opensafely.org/data-sources/apc/

13. Staa TP van, Fahmi A. Hospital admissions with infection-related complication [Internet]. 2021. Available from: https://www.opencodelists.org/codelist/user/alifahmi/hospital-admissions-with-infection-related-complication/29eaf6f8/

14. OpenSAFELY. Covid-19 test results [Internet]. [cited 2022 Jan 11]. Available from: https://docs.opensafely.org/data-sources/sgsscovid/

15. Fahmi A. hosp_pred amr-uom-brit [Internet]. 2022 [cited 2022 Aug 1]. Available from: https://github.com/opensafely/amr-uom-brit/tree/hosp_pred

16. Charlson ME, Pompei P, Ales KL, MacKenzie CRA. A New Method of Classifying Prognostic in Longitudinal Studies: Development and Validation. J Chronic Dis. 1987;40(5):373–383.

17. Zhang X, Staa V. Antibiotics codes [Internet]. Available from: https://www.opencodelists.org/codelist/user/BillyZhongUOM/brit_new_dmd/792101bd/

18. Foundation PS. Python 3.8.8 [Internet]. [cited 2022 Aug 1]. Available from: https://www.python.org/downloads/release/python-388/

19. Davidson-Pilon C. lifelines [Internet]. 2022 [cited 2022 Aug 1]. Available from: https://lifelines.readthedocs.io/en/latest/

20. Staa TP Van, Palin V, Li Y, et al. The effectiveness of frequent antibiotic use in reducing the risk of infection-related hospital admissions: Results from two large population-based cohorts. BMC Med. BMC Medicine; 2020;18(1):1–11.

21. NICE. Cough (acute): antimicrobial prescribing [Internet]. Natl. Inst. Heal. Care Excell. 2022. Available from: https://www.nice.org.uk/guidance/ng120/resources/cough-acute-antimicrobial-prescribing-pdf-66141652166341

22. NICE. Sore throat (acute): antimicrobial prescribing. NICE Guidel. 2018.

23. NICE. Urinary tract infection: antimicrobial prescribing. Nice 2018.

24. Hussain AZ, Paudyal V, Hadi MA. Impact of the COVID-19 pandemic on the prescribing patterns of first-line antibiotics in english primary care: A longitudinal analysis of national prescribing dataset. Antibiotics. MDPI AG; 2021 May 1;10(5).

25. Borek AJ, Maitland K, McLeod M, et al. Impact of the covid-19 pandemic on community antibiotic prescribing and stewardship: A qualitative interview study with general practitioners in england. Antibiotics. 2021;10(12).

26. Ruiz-Garbajosa P, Cantón R. Covid-19: Impact on prescribing and antimicrobial resistance. Rev Esp Quimioter. 2021;34:63–68.

27. Colliers A, Man J De, Adriaenssens N, et al. Antibiotic prescribing trends in belgian out-of-hours primary care during the covid-19 pandemic: Observational study using routinely collected health data. Antibiotics. 2021;10(12):1–10.

28. Caetano MC, Campos MR, Emmerick ICM, Luiza VL. Consumo de antimicrobianos nas farmácias e drogarias privadas brasileiras à luz do PAN-BR e da pandemia de COVID-19 / Antimicrobial consumption in Brazilian private pharmacies and drugstores considering the PAN-BR and COVID-19 pandemic. Brazilian J Dev. 2022;8(1):645–669.

29. Hek K, Ramerman L, Weesie YM, et al. Antibiotic Prescribing in Dutch Daytime and Out-of-Hours General Practice during the COVID-19 Pandemic: A Retrospective Database Study. Antibiotics. MDPI; 2022 Mar 1;11(3).

30. Palin V, Mölter A, Belmonte M, et al. Antibiotic prescribing for common infections in UK general practice: variability and drivers. J Antimicrob Chemother. 2019;74(8):2440–2450.

31. Weinstein EJ, Han JH, Lautenbach E, et al. A clinical prediction tool for extended-spectrum cephalosporin resistance in community-onset enterobacterales urinary tract infection. Open Forum Infect Dis. 2019;6(4):1–6.

32. Cole A, Telang J, Kim TK, et al. Infection-related hospitalization following ureteroscopic stone treatment: results from a surgical collaborative. BMC Urol. 2020;1–7.

33. Mooney C, Eogan M, Ní Áinle F, et al. Predicting bacteraemia in maternity patients using full blood count parameters: A supervised machine learning algorithm approach. Int J Lab Hematol. 2021;43(4):609–615.

34. Maat J van de, Nieboer D, Thompson M, Lakhanpaul M, Moll H, Oostenbrink R. Can clinical prediction models assess antibiotic need in childhood pneumonia? A validation study in paediatric emergency care. PLoS One. 2019;14(6):1–15.

35. Chen X, Zhou M, Yan Q, Jian Z, Liu W, Li H. Risk factors for carbapenem- - resistant Enterobacterales infection among hospitalized patients with previous colonization. 2022;(June):1–7.

36. Wang Y, Lin Q, Chen Z, et al. Construction of a risk prediction model for subsequent bloodstream infection in intestinal carriers of carbapenem-resistant enterobacteriaceae: A retrospective study in hematology department and intensive care unit. Infect Drug Resist. 2021;14:815–824.

37. NHS Digital (Now NHS England). The NHS England OpenSAFELY COVID-19 service - privacy notice [Internet]. [cited 2023 Jul 10]. Available from: https://digital.nhs.uk/coronavirus/coronavirus-covid-19-response-information-governance-hub/the-nhs-england-opensafely-covid-19-service-privacy-notice

38. NHS Digital (Now NHS England). Data Security and Protection Toolkit [Internet]. [cited 2023 Jul 10]. Available from: https://digital.nhs.uk/data-and-information/looking-after-information/data-security-and-information-governance/data-security-and-protection-toolkit

39. NHS Digital (Now NHS England). ISB1523: Anonymisation Standard for Publishing Health and Social Care Data [Internet]. [cited 2023 Jul 10]. Available from: https://digital.nhs.uk/data-and-information/information-standards/information-standards-and-data-collections-including-extractions/publications-and-notifications/standards-and-collections/isb1523-anonymisation-standard-for-publishing-health-and-social-car

40. Department of Health and Social Care. Coronavirus (COVID-19): notice under regulation 3(4) of the Health Service (Control of Patient Information) Regulations 2002 – general [Internet]. 2022 [cited 2023 Jul 10]. Available from: https://www.gov.uk/government/publications/coronavirus-covid-19-notification-of-data-controllers-to-share-information/coronavirus-covid-19-notice-under-regulation-34-of-the-health-service-control-of-patient-information-regulations-2002-general--2

41. Secretary of State for Health and Social Care - UK Government. COVID-19 Public Health Directions 2020: notification to NHS Digital [Internet]. 2020 [cited 2023 Jul 10]. Available from: https://digital.nhs.uk/about-nhs-digital/corporate-information-and-documents/directions-and-data-provision-notices/secretary-of-state-directions/covid-19-public-health-directions-2020

42. Health Research Authority. Confidentiality Advisory Group [Internet]. [cited 2023 Jul 10]. Available from: https://www.hra.nhs.uk/about-us/committees-and-services/confidentiality-advisory-group/

