## supplementary information for "Evaluation of the impact of COVID-19 pandemic on hospital admission related to common infections"

### Supplementary Tables

#### Baseline characteristics

Supplementary Table 1. Further baseline characteristics of cohort of incident common infections, including lower respiratory tract infection (LRTI), upper respiratory tract infection (URTI), and urinary tract infection (UTI), with no prescribed antibiotics.

|  | **Incident LRTI**  **with no ABs^1^** | **Incident URTI**  **with no ABs** | **Incident UTI**  **with no ABs** |
| --- | --- | --- | --- |
| **BMI^2^, N (%)** |  |  |  |
| Underweight | 7,295 (2.88) | 56,890 (2.22) | 6,600 (2.24) |
| Healthy weight | 62,175 (24.58) | 623,325 (24.28) | 78,325 (26.61) |
| Overweight | 65,930 (26.06) | 711,430 (27.71) | 75,895 (25.79) |
| Obese | 70,205 (27.75) | 727,810 (28.35) | 73,745 (25.06) |
| Unknown | 47,370 (18.72) | 447,785 (17.44) | 59,760 (20.30) |
| **Smoking status, N (%)** |  |  |  |
| Smoker | 42,535 (16.81) | 554,715 (21.61) | 35,500 (12.06) |
| Ex-smoker | 127,155 (50.26) | 1,216,215 (47.37) | 134,495 (45.70) |
| Never smoked | 82,260 (32.52) | 783,125 (30.50) | 122,840 (41.74) |
| Unknown | 1,025 (0.40) | 13,180 (0.51) | 1,495 (0.51) |
| **IMD^3^, N (%)** |  |  |  |
| 1 (most deprived) | 60,015 (23.72) | 633,025 (24.66) | 60,170 (20.44) |
| 2 | 49,970 (19.75) | 518,530 (20.20) | 57,195 (19.43) |
| 3 | 51,485 (20.35) | 522,010 (20.33) | 61,480 (20.89) |
| 4 | 46,170 (18.25) | 456,400 (17.78) | 57,275 (19.46) |
| 5 (most affluent) | 41,300 (16.33) | 395,710 (15.41) | 53,430 (18.15) |
| Unknown | 4,035 (1.59) | 41,560 (1.62) | 4,780 (1.62) |
| **Season, N (%)** |  |  |  |
| Spring | 65,035 (25.71) | 681,565 (26.55) | 76,355 (25.94) |
| Summer | 52,730 (20.84) | 572,105 (22.28) | 81,620 (27.73) |
| Autumn | 53,140 (21.01) | 525,865 (20.48) | 63,375 (21.53) |
| Winter | 82,070 (32.44) | 787,700 (30.68) | 72,975 (24.79) |
| **Region, N (%)** |  |  |  |
| London | 9,750 (3.85) | 103,185 (4.02) | 12,665 (4.30) |
| North East | 15,730 (6.22) | 168,565 (6.57) | 16,345 (5.55) |
| North West | 28,775 (11.38) | 310,845 (12.11) | 31,825 (10.81) |
| East | 58,080 (22.96) | 558,785 (21.77) | 68,935 (23.42) |
| West Midlands | 11,280 (4.46) | 99,320 (3.87) | 11,695 (3.97) |
| Yorkshire and The Humber | 40,920 (16.18) | 416,585 (16.23) | 43,225 (14.69) |
| South East | 15,945 (6.30) | 158,665 (6.18) | 20,580 (6.99) |
| East Midlands | 43,170 (17.07) | 450,550 (17.55) | 49,905 (16.96) |
| South West | 29,315 (11.59) | 300,740 (11.71) | 39,145 (13.30) |
| ^1^ ABs, antibiotics prescribed or not.  ^2^ BMI, Body Mass Index recorded in the last 5 years.  ^3^ IMD, Multiple Deprivation Index, quintile measured from patient-level address. | | | |

Supplementary Table 2. Baseline characteristics of cohort of incident common infections with prescribed antibiotics, prevalent common infections with no prescribed antibiotics, and prevalent common infections with prescribed antibiotics. Common infections include lower respiratory tract infection (LRTI), upper respiratory tract infection (URTI), and urinary tract infection (UTI).

|  | **LRTI** | | | **URTI** | | | **UTI** | | |
| --- | --- | --- | --- | --- | --- | --- | --- | --- | --- |
|  | **Incident** | **Prevalent** | | **Incident** | **Prevalent** | | **Incident** | **Prevalent** | |
|  | **With ABs^1^** | **No ABs** | **With ABs** | **With ABs** | **No ABs** | **With ABs** | **With ABs** | **No ABs** | **With ABs** |
| **Total, N cases** | 1,597,715 | 59,495 | 161,460 | 3,257,515 | 282,250 | 295,885 | 1,933,630 | 77,250 | 266,390 |
| **Age, N (%)** |  |  |  |  |  |  |  |  |  |
| 18-24 | 62,715 (3.93) | 1,370 (2.30) | 2,910 (1.80) | 248,575 (7.63) | 15,410 (5.46) | 15,355 (5.19) | 130,020 (6.72) | 4,455 (5.77) | 11,895 (4.47) |
| 25-34 | 145,930 (9.13) | 3,515 (5.90) | 8,995 (5.57) | 425,070 (13.05) | 26,585 (9.42) | 26,835 (9.07) | 218,530 (11.30) | 6,400 (8.29) | 19,500 (7.32) |
| 35-44 | 178,765 (11.19) | 4,915 (8.26) | 13,825 (8.56) | 416,590 (12.79) | 29,410 (10.42) | 29,990 (10.14) | 208,640 (10.79) | 5,725 (7.41) | 20,145 (7.56) |
| 45-54 | 252,655 (15.81) | 7,630 (12.83) | 22,600 (14.00) | 501,305 (15.39) | 43,235 (15.32) | 42,270 (14.29) | 260,745 (13.48) | 7,690 (9.96) | 29,720 (11.16) |
| 55-64 | 299,995 (18.78) | 9,665 (16.25) | 30,435 (18.85) | 562,065 (17.25) | 53,860 (19.08) | 53,750 (18.17) | 284,865 (14.73) | 10,180 (13.18) | 38,810 (14.57) |
| 65-74 | 309,880 (19.40) | 11,935 (20.06) | 35,960 (22.27) | 553,595 (16.99) | 56,955 (20.18) | 60,060 (20.30) | 362,020 (18.72) | 15,510 (20.08) | 59,940 (22.50) |
| 75+ | 347,770 (21.77) | 20,465 (34.40) | 46,735 (28.94) | 550,320 (16.89) | 56,795 (20.12) | 67,625 (22.86) | 468,810 (24.25) | 27,285 (35.32) | 86,380 (32.43) |
| **Sex, N (%)** |  |  |  |  |  |  |  |  |  |
| Male | 632,570 (39.59) | 23,690 (39.82) | 62,085 (38.45) | 1,233,165 (37.86) | 114,910 (40.71) | 111,185 (37.58) | 330,805 (17.11) | 22,090 (28.60) | 52,940 (19.87) |
| Female | 965,145 (60.41) | 35,805 (60.18) | 99,375 (61.55) | 2,024,350 (62.14) | 167,340 (59.29) | 184,700 (62.42) | 1,602,825 (82.89) | 55,160 (71.40) | 213,450 (80.13) |
| **BMI^2^, N (%)** |  |  |  |  |  |  |  |  |  |
| Underweight | 29,610 (1.85) | 1,640 (2.76) | 3,475 (2.15) | 59,035 (1.81) | 5,400 (1.91) | 5,965 (2.02) | 36,585 (1.89) | 1,615 (2.09) | 4,985 (1.87) |
| Healthy weight | 334,500 (20.94) | 14,670 (24.66) | 34,720 (21.50) | 693,470 (21.29) | 64,400 (22.82) | 63,810 (21.57) | 511,455 (26.45) | 20,725 (26.83) | 69,120 (25.95) |
| Overweight | 426,035 (26.67) | 15,860 (26.65) | 43,760 (27.10) | 838,830 (25.75) | 78,015 (27.64) | 79,455 (26.85) | 507,605 (26.25) | 21,065 (27.27) | 73,620 (27.64) |
| Obese | 504,440 (31.57) | 17,350 (29.16) | 54,335 (33.65) | 990,195 (30.40) | 84,540 (29.95) | 96,870 (32.74) | 499,085 (25.81) | 19,925 (25.79) | 71,895 (26.99) |
| Unknown | 303,125 (18.97) | 9,975 (16.77) | 25,170 (15.59) | 675,990 (20.75) | 49,900 (17.68) | 49,785 (16.83) | 378,895 (19.60) | 13,920 (18.02) | 46,765 (17.56) |
| **Ethnicity, N (%)** |  |  |  |  |  |  |  |  |  |
| White | 919,690 (57.56) | 36,730 (61.74) | 97,690 (60.51) | 1,824,280 (56.00) | 173,425 (61.44) | 176,335 (59.60) | 1,128,480 (58.36) | 47,195 (61.09) | 162,565 (61.02) |
| Non-White | 113,625 (7.11) | 2,960 (4.97) | 8,135 (5.04) | 277,660 (8.52) | 21,265 (7.53) | 19,455 (6.58) | 118,815 (6.14) | 3,645 (4.72) | 11,650 (4.37) |
| Unknown | 564,400 (35.33) | 19,805 (33.29) | 55,630 (34.46) | 1,155,580 (35.47) | 87,560 (31.02) | 100,095 (33.83) | 686,335 (35.49) | 26,415 (34.19) | 92,175 (34.60) |
| **CCI^3^, N (%)** |  |  |  |  |  |  |  |  |  |
| Very low | 855,855 (53.57) | 26,470 (44.49) | 72,130 (44.67) | 1,904,185 (58.46) | 158,745 (56.24) | 148,420 (50.16) | 1,222,700 (63.23) | 42,065 (54.45) | 150,215 (56.39) |
| Low | 584,155 (36.56) | 23,980 (40.30) | 67,430 (41.76) | 1,081,300 (33.19) | 94,915 (33.63) | 112,695 (38.09) | 543,415 (28.10) | 24,615 (31.87) | 84,495 (31.72) |
| Medium | 126,980 (7.95) | 6,965 (11.71) | 17,400 (10.78) | 219,755 (6.75) | 22,615 (8.01) | 27,620 (9.33) | 133,945 (6.93) | 7,975 (10.32) | 24,680 (9.26) |
| High | 23,670 (1.48) | 1,545 (2.59) | 3,450 (2.14) | 40,365 (1.24) | 4,555 (1.61) | 5,545 (1.87) | 26,290 (1.36) | 1,970 (2.55) | 5,355 (2.01) |
| Very high | 7,055 (0.44) | 540  (0.90) | 1,050 (0.65) | 11,910 (0.37) | 1,420 (0.50) | 1,605 (0.54) | 7,280 (0.38) | 620  (0.81) | 1,650 (0.62) |
| **Smoking status, N (%)** |  |  |  |  |  |  |  |  |  |
| Smoker | 318,060 (19.91) | 8,525 (14.33) | 28,870 (17.88) | 652,075 (20.02) | 47,345 (16.77) | 55,255 (18.68) | 225,800 (11.68) | 7,535 (9.76) | 26,145 (9.81) |
| Ex-smoker | 748,520 (46.85) | 31,450 (52.86) | 83,570 (51.76) | 1,439,045 (44.18) | 135,290 (47.93) | 144,090 (48.70) | 871,120 (45.05) | 37,545 (48.60) | 129,605 (48.65) |
| Never smoked | 526,150 (32.93) | 19,390 (32.59) | 48,790 (30.22) | 1,145,335 (35.16) | 98,570 (34.92) | 95,520 (32.28) | 830,300 (42.94) | 31,930 (41.33) | 110,080 (41.32) |
| Unknown | 4,985 (0.31) | 130  (0.22) | 225  (0.14) | 21,065 (0.65) | 1,040 (0.37) | 1,020 (0.34) | 6,410 (0.33) | 235  (0.30) | 565  (0.21) |
| **IMD^4^, N (%)** |  |  |  |  |  |  |  |  |  |
| 1 (most deprived) | 394,835 (24.71) | 12,850 (21.60) | 39,650 (24.56) | 802,480 (24.63) | 63,130 (22.37) | 71,715 (24.24) | 371,605 (19.22) | 13,625 (17.63) | 46,920 (17.61) |
| 2 | 324,645 (20.32) | 11,365 (19.10) | 32,360 (20.04) | 683,570 (20.98) | 54,955 (19.47) | 60,595 (20.48) | 363,030 (18.77) | 14,065 (18.21) | 48,055 (18.04) |
| 3 | 317,770 (19.89) | 12,310 (20.69) | 32,095 (19.88) | 650,670 (19.97) | 59,110 (20.94) | 60,445 (20.43) | 403,175 (20.85) | 16,360 (21.18) | 56,150 (21.08) |
| 4 | 288,195 (18.04) | 11,565 (19.44) | 29,700 (18.39) | 575,875 (17.68) | 52,600 (18.64) | 52,930 (17.89) | 391,825 (20.26) | 16,190 (20.96) | 55,930 (20.99) |
| 5 (most affluent) | 245,010 (15.34) | 10,470 (17.60) | 24,835 (15.38) | 492,020 (15.10) | 47,860 (16.96) | 45,130 (15.25) | 373,685 (19.33) | 15,780 (20.43) | 55,040 (20.66) |
| Unknown | 27,255 (1.71) | 930  (1.57) | 2,825 (1.75) | 52,905 (1.62) | 4,595 (1.63) | 5,070 (1.71) | 30,305 (1.57) | 1,230 (1.59) | 4,305 (1.62) |
| **Season, N (%)** |  |  |  |  |  |  |  |  |  |
| Spring | 382,590 (23.95) | 15,040 (25.28) | 38,775 (24.02) | 826,585 (25.37) | 80,630 (28.57) | 78,815 (26.64) | 501,500 (25.94) | 19,900 (25.76) | 69,425 (26.06) |
| Summer | 279,725 (17.51) | 10,670 (17.93) | 26,555 (16.45) | 619,035 (19.00) | 58,595 (20.76) | 52,175 (17.63) | 531,635 (27.49) | 20,735 (26.84) | 71,280 (26.76) |
| Autumn | 348,870 (21.84) | 11,630 (19.55) | 32,375 (20.05) | 695,680 (21.36) | 54,175 (19.19) | 58,120 (19.64) | 419,165 (21.68) | 17,185 (22.25) | 57,815 (21.70) |
| Winter | 586,530 (36.71) | 22,155 (37.24) | 63,755 (39.49) | 1,116,210 (34.27) | 88,850 (31.48) | 106,775 (36.09) | 481,330 (24.89) | 19,425 (25.15) | 67,870 (25.48) |
| **Region, N (%)** |  |  |  |  |  |  |  |  |  |
| London | 48,315 (3.02) | 1,570 (2.64) | 2,860 (1.77) | 146,275 (4.49) | 13,415 (4.75) | 10,335 (3.49) | 60,125 (3.11) | 2,705 (3.50) | 5,875 (2.21) |
| North East | 86,800 (5.43) | 3,650 (6.13) | 8,815 (5.46) | 171,105 (5.25) | 17,300 (6.13) | 16,320 (5.51) | 85,690 (4.43) | 3,775 (4.89) | 11,630 (4.37) |
| North West | 178,420 (11.17) | 7,590 (12.76) | 20,530 (12.71) | 351,360 (10.79) | 35,455 (12.56) | 36,910 (12.47) | 209,460 (10.83) | 8,895 (11.51) | 32,750 (12.29) |
| East | 368,845 (23.09) | 12,720 (21.38) | 35,580 (22.04) | 791,415 (24.30) | 59,410 (21.05) | 67,860 (22.93) | 461,430 (23.86) | 16,830 (21.79) | 61,440 (23.06) |
| West Midlands | 75,380 (4.72) | 2,480 (4.17) | 6,880 (4.26) | 163,510 (5.02) | 11,450 (4.06) | 13,000 (4.39) | 81,260 (4.20) | 2,810 (3.64) | 10,585 (3.97) |
| Yorkshire and The Humber | 307,885 (19.27) | 11,495 (19.32) | 32,925 (20.39) | 564,085 (17.32) | 49,285 (17.46) | 52,330 (17.69) | 312,910 (16.18) | 14,270 (18.47) | 45,570 (17.11) |
| South East | 85,310 (5.34) | 3,325 (5.59) | 8,055 (4.99) | 182,435 (5.60) | 17,265 (6.12) | 17,010 (5.75) | 118,940 (6.15) | 4,910 (6.35) | 15,710 (5.90) |
| East Midlands | 301,635 (18.88) | 10,795 (18.14) | 33,600 (20.81) | 589,625 (18.10) | 50,920 (18.04) | 56,900 (19.23) | 380,880 (19.70) | 13,945 (18.05) | 56,245 (21.11) |
| South West | 145,135 (9.08) | 5,865 (9.86) | 12,215 (7.57) | 297,705 (9.14) | 27,750 (9.83) | 25,220 (8.52) | 222,925 (11.53) | 9,105 (11.79) | 26,595 (9.98) |
| **Flu vaccination, N (%)** |  |  |  |  |  |  |  |  |  |
| Yes | 776,400 (48.59) | 34,015 (57.17) | 93,500 (57.91) | 1,393,175 (42.77) | 135,490 (48.00) | 153,540 (51.89) | 874,825 (45.24) | 41,370 (53.55) | 146,365 (54.94) |
| No | 821,315 (51.41) | 25,480 (42.83) | 67,960 (42.09) | 1,864,340 (57.23) | 146,760 (52.00) | 142,345 (48.11) | 1,058,800 (54.76) | 35,880 (46.45) | 120,030 (45.06) |
| **Period** |  |  |  |  |  |  |  |  |  |
| Pre-pandemic | 767,400 (48.03) | 29,730 (49.97) | 85,965 (53.24) | 1,413,980 (43.41) | 115,310 (40.85) | 136,365 (46.09) | 599,040 (30.98) | 25,625 (33.17) | 86,185 (32.35) |
| During pandemic | 342,045 (21.41) | 15,225 (25.59) | 38,430 (23.80) | 760,400 (23.34) | 83,620 (29.63) | 78,010 (26.36) | 663,955 (34.34) | 25,525 (33.04) | 91,880 (34.49) |
| After 2^nd^ lockdown | 488,265 (30.56) | 14,545 (24.44) | 37,060 (22.95) | 1,083,135 (33.25) | 83,320 (29.52) | 81,515 (27.55) | 670,630 (34.68) | 26,095 (33.78) | 88,325 (33.16) |
| **Count of antibiotic prescription, mean (SD^5^)** | 2.75  (2.41) | 3.39 (2.95) | 4.56 (2.78) | 2.53  (2.08) | 2.34 (2.31) | 3.88 (2.34) | 3.21  (2.89) | 4.07 (3.51) | 5.51 (3.46) |
| ^1^ ABs, antibiotics prescribed or not.  ^2^ BMI, Body Mass Index recorded in the last 5 years.  ^3^ CCI, Charlson Comorbidities Index, measured from 17 weighted conditions, including myocardial infarction, congestive heart failure, peripheral vascular disease, cerebrovascular disease, dementia, chronic pulmonary disease, Connective tissue disease, ulcer disease, mild liver disease, diabetes, hemiplegia, moderate or severe renal disease, diabetes with complications, any malignancy (including leukaemia and lymphoma), moderate or severe liver disease, metastatic solid tumour, and AIDS.  ^4^ IMD, Multiple Deprivation Index, quintile measured from patient-level address.  ^5^ SD, standard deviation. | | | | | | | | | |

Supplementary Table 3. Baseline characteristics of cohort of other common infections, including sinusitis, otitis media, and otitis externa. The cohort consists of incident infections with no prescribed antibiotics, incident infections with prescribed antibiotics, prevalent infections with no prescribed antibiotics, and prevalent infections with prescribed antibiotics.

|  | **Sinusitis** | | | | **Otitis media** | | | | **Otitis externa** | | | |
| --- | --- | --- | --- | --- | --- | --- | --- | --- | --- | --- | --- | --- |
|  | **Incident** | | **Prevalent** | | **Incident** | | **Prevalent** | | **Incident** | | **Prevalent** | |
|  | **No ABs^1^** | **With ABs** | **No ABs** | **With ABs** | **No ABs** | **With ABs** | **No ABs** | **With ABs** | **No ABs** | **With ABs** | **No ABs** | **With ABs** |
| **Total, N cases** | 129,780 | 363,670 | 14,225 | 26,280 | 80,955 | 228,710 | 9,905 | 15,685 | 590,740 | 157,775 | 57,675 | 23,640 |
| **Age, N (%)** |  |  |  |  |  |  |  |  |  |  |  |  |
| 18-24 | 9,295 (7.16) | 19,165 (5.27) | 775 (5.43) | 1,025 (3.90) | 8,385 (10.36) | 23,955 (10.47) | 865 (8.75) | 1,310 (8.36) | 48,800 (8.26) | 13,275 (8.41) | 4,385 (7.60) | 1,690 (7.14) |
| 25-34 | 23,955 (18.46) | 60,220 (16.56) | 2,305 (16.20) | 3,865 (14.71) | 15,040 (18.58) | 47,160 (20.62) | 1,810 (18.29) | 3,205 (20.44) | 92,520 (15.66) | 27,500 (17.43) | 8,365 (14.51) | 3,825 (16.18) |
| 35-44 | 25,250 (19.46) | 72,575 (19.96) | 2,835 (19.92) | 5,395 (20.52) | 13,145 (16.24) | 43,030 (18.81) | 1,690 (17.04) | 3,005 (19.15) | 91,785 (15.54) | 27,250 (17.27) | 8,270 (14.34) | 3,850 (16.29) |
| 45-54 | 25,735 (19.83) | 78,275 (21.52) | 3,125 (21.97) | 6,020 (22.90) | 13,900 (17.17) | 41,755 (18.26) | 1,835 (18.54) | 2,965 (18.90) | 107,095 (18.13) | 30,680 (19.45) | 10,100 (17.51) | 4,300 (18.20) |
| 55-64 | 22,040 (16.98) | 67,575 (18.58) | 2,580 (18.12) | 5,060 (19.26) | 12,545 (15.50) | 33,675 (14.72) | 1,570 (15.84) | 2,395 (15.26) | 99,665 (16.87) | 26,245 (16.64) | 9,585 (16.62) | 3,970 (16.79) |
| 65-74 | 16,255 (12.53) | 47,310 (13.01) | 1,895 (13.33) | 3,660 (13.93) | 10,410 (12.86) | 24,605 (10.76) | 1,230 (12.43) | 1,775 (11.32) | 88,855 (15.04) | 20,015 (12.69) | 9,215 (15.98) | 3,360 (14.22) |
| 75+ | 7,240 (5.58) | 18,550 (5.10) | 715 (5.03) | 1,255 (4.78) | 7,530 (9.30) | 14,530 (6.35) | 900 (9.11) | 1,030 (6.57) | 62,020 (10.50) | 12,805 (8.12) | 7,750 (13.44) | 2,645 (11.19) |
| **Sex, N (%)** |  |  |  |  |  |  |  |  |  |  |  |  |
| Male | 45,125 (34.77) | 96,105 (26.43) | 4,450 (31.30) | 6,780 (25.80) | 33,005 (40.77) | 80,280 (35.10) | 3,890 (39.29) | 5,705 (36.37) | 244,595 (41.40) | 58,715 (37.21) | 24,275 (42.09) | 9,340 (39.51) |
| Female | 84,655 (65.23) | 267,565 (73.57) | 9,775 (68.70) | 19,500 (74.20) | 47,950 (59.23) | 148,430 (64.90) | 6,015 (60.71) | 9,980 (63.63) | 346,145 (58.60) | 99,060 (62.79) | 33,400 (57.91) | 14,300 (60.49) |
| **BMI^2^, N (%)** |  |  |  |  |  |  |  |  |  |  |  |  |
| Underweight | 1,750 (1.35) | 4,750 (1.31) | 190 (1.33) | 395 (1.51) | 1,060 (1.31) | 2,865 (1.25) | 110 (1.09) | 190 (1.21) | 7,210 (1.22) | 1,815 (1.15) | 670 (1.16) | 260 (1.10) |
| Healthy weight | 32,245 (24.85) | 86,240 (23.71) | 3,535 (24.85) | 6,170 (23.48) | 17,265 (21.32) | 44,755 (19.57) | 2,015 (20.35) | 2,840 (18.10) | 122,745 (20.78) | 29,700 (18.83) | 11,720 (20.32) | 4,465 (18.89) |
| Overweight | 33,900 (26.12) | 92,995 (25.57) | 3,785 (26.62) | 6,870 (26.13) | 19,860 (24.53) | 54,955 (24.03) | 2,430 (24.54) | 3,855 (24.59) | 152,375 (25.79) | 38,285 (24.27) | 15,430 (26.76) | 6,035 (25.53) |
| Obese | 30,790 (23.73) | 95,635 (26.30) | 3,515 (24.70) | 7,270 (27.65) | 21,510 (26.57) | 66,160 (28.93) | 2,820 (28.46) | 4,825 (30.77) | 161,350 (27.31) | 48,845 (30.96) | 16,710 (28.97) | 7,470 (31.59) |
| Unknown | 31,090 (23.96) | 84,050 (23.11) | 3,200 (22.51) | 5,580 (21.23) | 21,260 (26.26) | 59,980 (26.23) | 2,530 (25.57) | 3,975 (25.34) | 147,055 (24.89) | 39,125 (24.80) | 13,140 (22.78) | 5,410 (22.89) |
| **Ethnicity, N (%)** |  |  |  |  |  |  |  |  |  |  |  |  |
| White | 76,955 (59.30) | 215,675 (59.31) | 9,065 (63.71) | 16,400 (62.39) | 45,700 (56.45) | 126,560 (55.34) | 5,710 (57.64) | 8,905 (56.75) | 339,635 (57.49) | 90,790 (57.54) | 34,595 (59.99) | 14,025 (59.32) |
| Non-White | 10,210 (7.87) | 23,785 (6.54) | 895 (6.31) | 1,305 (4.97) | 7,020 (8.67) | 19,975 (8.73) | 840 (8.49) | 1,240 (7.89) | 45,925 (7.77) | 11,010 (6.98) | 3,845 (6.67) | 1,385 (5.86) |
| Unknown | 42,615 (32.84) | 124,210 (34.15) | 4,265 (29.98) | 8,580 (32.64) | 28,235 (34.88) | 82,180 (35.93) | 3,355 (33.87) | 5,545 (35.36) | 205,175 (34.73) | 55,975 (35.48) | 19,235 (33.35) | 8,235 (34.83) |
| **CCI^3^, N (%)** |  |  |  |  |  |  |  |  |  |  |  |  |
| Very low | 94,135 (72.54) | 259,605 (71.38) | 10,065 (70.76) | 17,915 (68.16) | 58,835 (72.68) | 167,040 (73.03) | 7,100 (71.69) | 11,195 (71.35) | 426,885 (72.26) | 111,905 (70.93) | 39,885 (69.16) | 16,175 (68.41) |
| Low | 31,135 (23.99) | 90,195 (24.80) | 3,590 (25.24) | 7,280 (27.70) | 18,355 (22.67) | 52,235 (22.84) | 2,305 (23.29) | 3,805 (24.24) | 134,470 (22.76) | 37,820 (23.97) | 14,035 (24.33) | 5,900 (24.96) |
| Medium | 3,735 (2.88) | 11,680 (3.21) | 475 (3.35) | 905 (3.44) | 3,070 (3.79) | 7,790 (3.41) | 400 (4.05) | 555 (3.52) | 24,185 (4.09) | 6,610 (4.19) | 2,980 (5.17) | 1,275 (5.39) |
| High | 615 (0.47) | 1,685 (0.46) | 75 (0.53) | 145 (0.54) | 540 (0.66) | 1,300 (0.57) | 70 (0.73) | 100 (0.64) | 4,110 (0.70) | 1,145 (0.73) | 630 (1.09) | 250 (1.05) |
| Very high | 160 (0.12) | 505 (0.14) | 15 (0.12) | 40  (0.16) | 155 (0.19) | 345 (0.15) | 25 (0.24) | 40  (0.25) | 1,085 (0.18) | 295 (0.19) | 145 (0.25) | 45  (0.19) |
| **Smoking status, N (%)** |  |  |  |  |  |  |  |  |  |  |  |  |
| Smoker | 19,005 (14.64) | 58,120 (15.98) | 2,095 (14.73) | 4,200 (15.98) | 14,585 (18.02) | 45,395 (19.85) | 1,745 (17.62) | 2,945 (18.77) | 96,410 (16.32) | 32,190 (20.40) | 9,135 (15.84) | 4,225 (17.86) |
| Ex-smoker | 54,270 (41.82) | 153,350 (42.17) | 6,140 (43.15) | 11,485 (43.71) | 32,120 (39.68) | 88,220 (38.57) | 4,005 (40.43) | 6,395 (40.76) | 247,825 (41.95) | 65,010 (41.20) | 24,970 (43.30) | 10,275 (43.45) |
| Never smoked | 55,710 (42.93) | 150,540 (41.39) | 5,935 (41.72) | 10,530 (40.06) | 33,395 (41.25) | 92,860 (40.60) | 4,090 (41.29) | 6,230 (39.72) | 241,360 (40.86) | 59,355 (37.62) | 23,200 (40.23) | 9,015 (38.14) |
| Unknown | 795 (0.61) | 1,660 (0.46) | 55 (0.40) | 65  (0.25) | 855 (1.05) | 2,235 (0.98) | 65 (0.67) | 115 (0.74) | 5,145 (0.87) | 1,215 (0.77) | 370 (0.64) | 130 (0.55) |
| **IMD^4^, N (%)** |  |  |  |  |  |  |  |  |  |  |  |  |
| 1 (most deprived) | 25,195 (19.41) | 65,785 (18.09) | 2,610 (18.35) | 4,705 (17.91) | 19,045 (23.53) | 57,735 (25.24) | 2,260 (22.83) | 4,000 (25.48) | 123,205 (20.86) | 36,280 (22.99) | 11,350 (19.68) | 4,895 (20.70) |
| 2 | 24,695 (19.03) | 69,350 (19.07) | 2,655 (18.66) | 5,005 (19.04) | 16,460 (20.33) | 47,930 (20.96) | 1,885 (19.02) | 3,270 (20.84) | 113,150 (19.15) | 32,170 (20.39) | 10,480 (18.17) | 4,480 (18.94) |
| 3 | 26,990 (20.80) | 77,180 (21.22) | 3,035 (21.34) | 5,475 (20.83) | 16,050 (19.83) | 44,490 (19.45) | 1,965 (19.82) | 2,915 (18.59) | 121,115 (20.50) | 32,085 (20.33) | 11,865 (20.57) | 4,900 (20.72) |
| 4 | 26,285 (20.25) | 75,540 (20.77) | 2,875 (20.21) | 5,520 (20.99) | 14,730 (18.19) | 40,380 (17.65) | 1,865 (18.85) | 2,845 (18.14) | 115,040 (19.47) | 29,235 (18.53) | 11,680 (20.25) | 4,760 (20.13) |
| 5 (most affluent) | 24,445 (18.84) | 69,850 (19.21) | 2,825 (19.85) | 5,135 (19.54) | 13,315 (16.45) | 34,380 (15.03) | 1,770 (17.87) | 2,400 (15.31) | 108,955 (18.44) | 25,465 (16.14) | 11,380 (19.74) | 4,225 (17.88) |
| Unknown | 2,165 (1.67) | 5,965 (1.64) | 230 (1.60) | 445 (1.69) | 1,355 (1.67) | 3,800 (1.66) | 160 (1.61) | 255 (1.63) | 9,270 (1.57) | 2,540 (1.61) | 915 (1.59) | 385 (1.63) |
| **Season, N (%)** |  |  |  |  |  |  |  |  |  |  |  |  |
| Spring | 35,340 (27.23) | 98,140 (26.99) | 4,150 (29.19) | 7,400 (28.16) | 20,030 (24.74) | 56,450 (24.68) | 2,415 (24.40) | 3,865 (24.63) | 140,505 (23.78) | 37,460 (23.74) | 13,130 (22.76) | 5,250 (22.21) |
| Summer | 26,140 (20.14) | 66,950 (18.41) | 2,760 (19.42) | 4,705 (17.90) | 20,855 (25.76) | 56,320 (24.62) | 2,340 (23.63) | 3,835 (24.45) | 163,315 (27.65) | 42,910 (27.20) | 15,725 (27.26) | 6,455 (27.31) |
| Autumn | 27,350 (21.07) | 74,705 (20.54) | 2,850 (20.04) | 5,210 (19.83) | 17,370 (21.46) | 48,805 (21.34) | 2,150 (21.73) | 3,310 (21.10) | 132,540 (22.44) | 35,770 (22.67) | 13,910 (24.12) | 5,765 (24.39) |
| Winter | 40,945 (31.55) | 123,875 (34.06) | 4,460 (31.35) | 8,965 (34.11) | 22,700 (28.04) | 67,140 (29.35) | 2,995 (30.24) | 4,680 (29.83) | 154,375 (26.13) | 41,635 (26.39) | 14,910 (25.86) | 6,170 (26.10) |
| **Region, N (%)** |  |  |  |  |  |  |  |  |  |  |  |  |
| London | 5,975 (4.60) | 13,020 (3.58) | 545 (3.83) | 675 (2.56) | 3,705 (4.58) | 8,610 (3.77) | 400 (4.03) | 490 (3.13) | 24,035 (4.07) | 6,100 (3.87) | 2,120 (3.68) | 690 (2.92) |
| North East | 6,970 (5.37) | 15,830 (4.35) | 745 (5.24) | 1,065 (4.04) | 4,745 (5.86) | 11,415 (4.99) | 525 (5.29) | 810 (5.17) | 31,355 (5.31) | 8,115 (5.14) | 3,135 (5.44) | 1,300 (5.51) |
| North West | 16,170 (12.46) | 38,655 (10.63) | 1,955 (13.74) | 3,285 (12.50) | 9,245 (11.42) | 24,820 (10.85) | 1,230 (12.41) | 1,855 (11.82) | 66,295 (11.22) | 16,230 (10.29) | 6,945 (12.04) | 2,605 (11.02) |
| East | 29,660 (22.85) | 98,970 (27.21) | 3,315 (23.30) | 7,025 (26.72) | 21,200 (26.18) | 60,195 (26.32) | 2,540 (25.64) | 4,155 (26.50) | 132,125 (22.37) | 40,050 (25.38) | 12,500 (21.67) | 5,745 (24.29) |
| West Midlands | 4,610 (3.55) | 12,660 (3.48) | 475 (3.34) | 795 (3.02) | 3,545 (4.38) | 10,245 (4.48) | 440 (4.43) | 700 (4.46) | 21,360 (3.62) | 5,855 (3.71) | 1,685 (2.93) | 720 (3.04) |
| Yorkshire and The Humber | 22,195 (17.10) | 58,030 (15.96) | 2,535 (17.81) | 4,440 (16.89) | 13,720 (16.95) | 40,330 (17.63) | 1,815 (18.32) | 2,730 (17.41) | 98,195 (16.62) | 25,335 (16.06) | 9,995 (17.33) | 3,840 (16.24) |
| South East | 8,495 (6.55) | 22,745 (6.25) | 810 (5.68) | 1,640 (6.24) | 4,365 (5.39) | 11,455 (5.01) | 525 (5.30) | 735 (4.70) | 39,155 (6.63) | 9,865 (6.25) | 4,070 (7.06) | 1,585 (6.70) |
| East Midlands | 21,140 (16.29) | 67,170 (18.47) | 2,590 (18.20) | 5,125 (19.50) | 13,405 (16.56) | 43,235 (18.90) | 1,760 (17.77) | 3,225 (20.56) | 109,585 (18.55) | 30,575 (19.38) | 10,845 (18.81) | 4,855 (20.53) |
| South West | 14,570 (11.23) | 36,585 (10.06) | 1,260 (8.86) | 2,240 (8.52) | 7,020 (8.67) | 18,410 (8.05) | 675 (6.82) | 980 (6.26) | 68,640 (11.62) | 15,650 (9.92) | 6,380 (11.06) | 2,305 (9.76) |
| **Flu vaccination, N (%)** |  |  |  |  |  |  |  |  |  |  |  |  |
| Yes | 37,070 (28.57) | 109,615 (30.14) | 4,335 (30.48) | 8,660 (32.95) | 24,400 (30.14) | 62,765 (27.44) | 3,090 (31.18) | 4,680 (29.82) | 195,500 (33.09) | 48,930 (31.01) | 21,435 (37.16) | 8,400 (35.52) |
| No | 92,705 (71.43) | 254,055 (69.86) | 9,890 (69.52) | 17,625 (67.05) | 56,555 (69.86) | 165,945 (72.56) | 6,815 (68.82) | 11,010 (70.18) | 395,240 (66.91) | 108,845 (68.99) | 36,240 (62.84) | 15,245 (64.48) |
| **Period** |  |  |  |  |  |  |  |  |  |  |  |  |
| Pre-pandemic | 50,625 (39.01) | 141,755 (38.98) | 5,225 (36.72) | 9,960 (37.90) | 29,565 (36.52) | 82,880 (36.24) | 3,840 (38.75) | 6,090 (38.81) | 223,040 (37.76) | 54,840 (34.76) | 22,320 (38.70) | 9,195 (38.89) |
| During pandemic | 36,965 (28.49) | 100,710 (27.69) | 4,535 (31.89) | 8,310 (31.63) | 24,165 (29.85) | 72,505 (31.70) | 3,180 (32.11) | 5,140 (32.75) | 176,710 (29.91) | 51,230 (32.47) | 17,685 (30.66) | 7,660 (32.39) |
| After 2^nd^ lockdown | 42,185 (32.51) | 121,205 (33.33) | 4,465 (31.39) | 8,010 (30.47) | 27,225 (33.63) | 73,325 (32.06) | 2,885 (29.14) | 4,460 (28.44) | 190,990 (32.33) | 51,705 (32.77) | 17,670 (30.64) | 6,790 (28.71) |
| **Count of antibiotic prescription, mean (SD^5^)** | 1.15 (1.69) | 2.31 (1.73) | 2.39 (1.97) | 3.52 (1.93) | 1.07 (1.55) | 2.07 (1.49) | 2.07 (1.65) | 3.07 (1.54) | 0.91 (1.38) | 2.05 (1.47) | 1.51 (1.58) | 2.49 (1.50) |
| ^1^ ABs, antibiotics prescribed or not.  ^2^ BMI, Body Mass Index recorded in the last 5 years.  ^3^ CCI, Charlson Comorbidities Index, measured from 17 weighted conditions, including myocardial infarction, congestive heart failure, peripheral vascular disease, cerebrovascular disease, dementia, chronic pulmonary disease, Connective tissue disease, ulcer disease, mild liver disease, diabetes, hemiplegia, moderate or severe renal disease, diabetes with complications, any malignancy (including leukaemia and lymphoma), moderate or severe liver disease, metastatic solid tumour, and AIDS.  ^4^ IMD, Multiple Deprivation Index, quintile measured from patient-level address.  ^5^ SD, standard deviation. | | | | | | | | | | | | |

Supplementary Table 4. Baseline characteristics of cohort of URTI infections, including specific upper respiratory tract infection (URTI), cough, cold with cough, and sore throat. The cohort consists of incident infections with no prescribed antibiotics, incident infections with prescribed antibiotics, prevalent infections with no prescribed antibiotics, and prevalent infections with prescribed antibiotics.

|  | **Specific URTI** | | | | **Cough** | | | | **Cold with cough** | | | | **Sore throat** | | | |
| --- | --- | --- | --- | --- | --- | --- | --- | --- | --- | --- | --- | --- | --- | --- | --- | --- |
|  | **Incident** | | **Prevalent** | | **Incident** | | **Prevalent** | | **Incident** | | **Prevalent** | | **Incident** | | **Prevalent** | |
|  | **No ABs^1^** | **With ABs** | **No ABs** | **With ABs** | **No ABs** | **With ABs** | **No ABs** | **With ABs** | **No ABs** | **With ABs** | **No ABs** | **With ABs** | **No ABs** | **With ABs** | **No ABs** | **With ABs** |
| **Total, N cases** | 213,345 | 405,825 | 9,725 | 16,790 | 944,865 | 637,095 | 83,305 | 49,385 | 1,138,815 | 1,683,095 | 159,260 | 199,440 | 270,215 | 531,510 | 29,965 | 30,270 |
| **Age, N (%)** |  |  |  |  |  |  |  |  |  |  |  |  |  |  |  |  |
| 18-24 | 22,940 (10.75) | 24,225 (5.97) | 640 (6.58) | 625 (3.72) | 35,675 (3.78) | 26,370 (4.14) | 2,550 (3.06) | 1,275 (2.59) | 48,170 (4.23) | 66,595 (3.96) | 4,655 (2.92) | 4,300 (2.16) | 51,510 (19.06) | 131,385 (24.72) | 7,565 (25.25) | 9,155 (30.25) |
| 25-34 | 39,040 (18.30) | 51,680 (12.73) | 1,320 (13.57) | 1,650 (9.83) | 67,755 (7.17) | 59,665 (9.37) | 5,865 (7.04) | 3,460 (7.00) | 91,055 (8.00) | 153,255 (9.11) | 11,310 (7.10) | 12,335 (6.18) | 65,345 (24.18) | 160,465 (30.19) | 8,090 (27.00) | 9,395 (31.03) |
| 35-44 | 35,625 (16.70) | 58,105 (14.32) | 1,470 (15.13) | 2,115 (12.61) | 80,440 (8.51) | 71,945 (11.29) | 7,975 (9.57) | 4,745 (9.61) | 103,355 (9.08) | 186,650 (11.09) | 15,090 (9.48) | 17,865 (8.96) | 48,420 (17.92) | 99,890 (18.79) | 4,870 (16.26) | 5,265 (17.39) |
| 45-54 | 36,070 (16.91) | 69,580 (17.14) | 1,760 (18.11) | 2,740 (16.31) | 135,385 (14.33) | 102,865 (16.15) | 13,510 (16.22) | 7,655 (15.50) | 161,580 (14.19) | 265,190 (15.76) | 24,215 (15.21) | 28,855 (14.47) | 39,535 (14.63) | 63,665 (11.98) | 3,745 (12.50) | 3,015 (9.96) |
| 55-64 | 32,135 (15.06) | 73,960 (18.23) | 1,705 (17.56) | 3,225 (19.21) | 195,775 (20.72) | 126,995 (19.93) | 18,115 (21.75) | 10,210 (20.67) | 222,365 (19.53) | 320,675 (19.05) | 31,420 (19.73) | 38,435 (19.27) | 30,575 (11.32) | 40,430 (7.61) | 2,615 (8.73) | 1,880 (6.20) |
| 65-74 | 24,890 (11.67) | 66,385 (16.36) | 1,335 (13.75) | 3,165 (18.86) | 236,800 (25.06) | 130,900 (20.55) | 19,325 (23.20) | 11,365 (23.01) | 265,040 (23.27) | 333,285 (19.80) | 34,345 (21.57) | 44,500 (22.31) | 21,485 (7.95) | 23,025 (4.33) | 1,945 (6.50) | 1,030 (3.40) |
| 75+ | 22,650 (10.62) | 61,885 (15.25) | 1,485 (15.30) | 3,265 (19.46) | 193,035 (20.43) | 118,355 (18.58) | 15,960 (19.16) | 10,670 (21.61) | 247,250 (21.71) | 357,435 (21.24) | 38,225 (24.00) | 53,155 (26.65) | 13,340 (4.94) | 12,640 (2.38) | 1,125 (3.76) | 535 (1.77) |
| **Sex, N (%)** |  |  |  |  |  |  |  |  |  |  |  |  |  |  |  |  |
| Male | 74,440 (34.89) | 149,380 (36.81) | 3,120 (32.08) | 5,865 (34.93) | 432,820 (45.81) | 253,790 (39.84) | 35,185 (42.23) | 19,195 (38.87) | 516,745 (45.38) | 669,540 (39.78) | 66,325 (41.65) | 77,620 (38.92) | 92,730 (34.32) | 160,460 (30.19) | 10,280 (34.31) | 8,505 (28.10) |
| Female | 138,905 (65.11) | 256,445 (63.19) | 6,605 (67.92) | 10,925 (65.07) | 512,045 (54.19) | 383,305 (60.16) | 48,120 (57.77) | 30,190 (61.13) | 622,070 (54.62) | 1,013,555 (60.22) | 92,935 (58.35) | 121,820 (61.08) | 177,485 (65.68) | 371,050 (69.81) | 19,685 (65.69) | 21,765 (71.90) |
| **BMI^2^, N (%)** |  |  |  |  |  |  |  |  |  |  |  |  |  |  |  |  |
| Underweight | 3,550 (1.66) | 6,775 (1.67) | 190 (1.93) | 270 (1.62) | 21,870 (2.31) | 12,065 (1.89) | 1,440 (1.73) | 970 (1.97) | 26,780 (2.35) | 31,830 (1.89) | 3,240 (2.04) | 4,175 (2.09) | 4,690 (1.74) | 8,365 (1.57) | 530 (1.78) | 545 (1.80) |
| Healthy weight | 48,700 (22.83) | 84,815 (20.90) | 2,065 (21.22) | 3,555 (21.17) | 231,510 (24.50) | 135,555 (21.28) | 18,675 (22.42) | 10,490 (21.24) | 278,465 (24.45) | 355,615 (21.13) | 36,480 (22.90) | 42,960 (21.54) | 64,650 (23.93) | 117,490 (22.11) | 7,185 (23.97) | 6,810 (22.49) |
| Overweight | 52,505 (24.61) | 106,210 (26.17) | 2,550 (26.23) | 4,485 (26.72) | 274,235 (29.02) | 171,445 (26.91) | 24,030 (28.84) | 13,785 (27.92) | 323,185 (28.38) | 450,385 (26.76) | 44,880 (28.18) | 54,985 (27.57) | 61,505 (22.76) | 110,785 (20.84) | 6,555 (21.88) | 6,195 (20.47) |
| Obese | 55,855 (26.18) | 124,585 (30.70) | 2,935 (30.19) | 5,720 (34.06) | 279,690 (29.60) | 203,495 (31.94) | 25,990 (31.20) | 16,835 (34.09) | 329,695 (28.95) | 531,540 (31.58) | 48,850 (30.67) | 66,695 (33.44) | 62,570 (23.16) | 130,575 (24.57) | 6,760 (22.57) | 7,625 (25.19) |
| Unknown | 52,735 (24.72) | 83,440 (20.56) | 1,985 (20.43) | 2,760 (16.43) | 137,560 (14.56) | 114,530 (17.98) | 13,170 (15.81) | 7,305 (14.80) | 180,690 (15.87) | 313,730 (18.64) | 25,810 (16.20) | 30,620 (15.35) | 76,800 (28.42) | 164,290 (30.91) | 8,930 (29.81) | 9,095 (30.05) |
| **Ethnicity, N (%)** |  |  |  |  |  |  |  |  |  |  |  |  |  |  |  |  |
| White | 117,535 (55.09) | 212,030 (52.25) | 5,815 (59.80) | 9,170 (54.61) | 575,975 (60.96) | 365,930 (57.44) | 52,790 (63.37) | 30,165 (61.08) | 683,380 (60.01) | 973,120 (57.82) | 98,255 (61.69) | 120,840 (60.59) | 142,505 (52.74) | 273,195 (51.40) | 16,565 (55.28) | 16,160 (53.38) |
| Non-White | 22,970 (10.77) | 51,605 (12.72) | 1,010 (10.41) | 1,920 (11.42) | 57,610 (6.10) | 51,940 (8.15) | 5,905 (7.09) | 3,335 (6.75) | 76,125 (6.68) | 120,085 (7.13) | 11,425 (7.17) | 11,505 (5.77) | 33,360 (12.35) | 54,030 (10.17) | 2,925 (9.75) | 2,700 (8.92) |
| Unknown | 72,840 (34.14) | 142,185 (35.04) | 2,895 (29.79) | 5,705 (33.97) | 311,280 (32.94) | 219,225 (34.41) | 24,610 (29.54) | 15,890 (32.17) | 379,310 (33.31) | 589,885 (35.05) | 49,580 (31.13) | 67,095 (33.64) | 94,350 (34.92) | 204,280 (38.43) | 10,480 (34.97) | 11,410 (37.69) |
| **CCI^3^, N (%)** |  |  |  |  |  |  |  |  |  |  |  |  |  |  |  |  |
| Very low | 149,165 (69.92) | 243,980 (60.12) | 6,080 (62.54) | 9,025 (53.76) | 516,885 (54.70) | 343,300 (53.89) | 46,310 (55.59) | 23,680 (47.95) | 626,710 (55.03) | 898,575 (53.39) | 83,250 (52.27) | 92,205 (46.23) | 206,620 (76.47) | 418,330 (78.71) | 23,105 (77.10) | 23,510 (77.67) |
| Low | 52,985 (24.84) | 130,575 (32.18) | 2,895 (29.78) | 6,040 (35.97) | 338,830 (35.86) | 232,850 (36.55) | 28,710 (34.47) | 19,585 (39.66) | 400,250 (35.15) | 617,750 (36.70) | 57,345 (36.01) | 81,035 (40.63) | 54,385 (20.13) | 100,130 (18.84) | 5,965 (19.91) | 6,035 (19.93) |
| Medium | 9,005 (4.22) | 25,325 (6.24) | 590 (6.09) | 1,405 (8.36) | 72,855 (7.71) | 49,440 (7.76) | 6,655 (7.99) | 4,900 (9.92) | 90,190 (7.92) | 134,455 (7.99) | 14,670 (9.21) | 20,755 (10.41) | 7,390 (2.73) | 10,535 (1.98) | 695 (2.33) | 565 (1.86) |
| High | 1,695 (0.79) | 4,585 (1.13) | 115 (1.20) | 245 (1.45) | 12,930 (1.37) | 8,950 (1.41) | 1,250 (1.50) | 965 (1.95) | 16,815 (1.48) | 24,900 (1.48) | 3,025 (1.90) | 4,225 (2.12) | 1,365 (0.51) | 1,925 (0.36) | 160 (0.53) | 115 (0.38) |
| Very high | 495 (0.23) | 1,360 (0.34) | 40 (0.39) | 75  (0.45) | 3,360 (0.36) | 2,555 (0.40) | 370 (0.45) | 260 (0.52) | 4,845 (0.43) | 7,410 (0.44) | 970 (0.61) | 1,225 (0.61) | 455 (0.17) | 585 (0.11) | 40  (0.13) | 45  (0.16) |
| **Smoking status, N (%)** |  |  |  |  |  |  |  |  |  |  |  |  |  |  |  |  |
| Smoker | 33,735 (15.81) | 73,295 (18.06) | 1,340 (13.80) | 2,780 (16.55) | 221,800 (23.47) | 135,415 (21.26) | 14,020 (16.83) | 9,665 (19.57) | 254,580 (22.35) | 341,970 (20.32) | 26,645 (16.73) | 37,260 (18.68) | 44,595 (16.50) | 101,395 (19.08) | 5,340 (17.81) | 5,550 (18.34) |
| Ex-smoker | 83,810 (39.28) | 171,750 (42.32) | 4,275 (43.95) | 7,695 (45.82) | 473,645 (50.13) | 296,980 (46.61) | 40,975 (49.19) | 24,520 (49.65) | 563,125 (49.45) | 791,895 (47.05) | 79,645 (50.01) | 101,795 (51.04) | 95,635 (35.39) | 178,415 (33.57) | 10,400 (34.71) | 10,080 (33.30) |
| Never smoked | 93,855 (43.99) | 158,810 (39.13) | 4,065 (41.80) | 6,285 (37.43) | 246,825 (26.12) | 202,870 (31.84) | 28,175 (33.82) | 15,140 (30.66) | 317,175 (27.85) | 544,045 (32.32) | 52,655 (33.06) | 60,095 (30.13) | 125,275 (46.36) | 239,610 (45.08) | 13,680 (45.65) | 14,000 (46.25) |
| Unknown | 1,940 (0.91) | 1,970 (0.49) | 45 (0.44) | 35  (0.20) | 2,590 (0.27) | 1,830 (0.29) | 130 (0.16) | 60  (0.13) | 3,935 (0.35) | 5,180 (0.31) | 315 (0.20) | 285 (0.14) | 4,710 (1.74) | 12,085 (2.27) | 550 (1.84) | 640 (2.11) |
| **IMD^4^, N (%)** |  |  |  |  |  |  |  |  |  |  |  |  |  |  |  |  |
| 1 (most deprived) | 46,160 (21.64) | 102,400 (25.23) | 2,060 (21.21) | 4,445 (26.47) | 238,030 (25.19) | 161,155 (25.30) | 18,460 (22.16) | 12,140 (24.58) | 283,945 (24.93) | 412,920 (24.53) | 36,155 (22.70) | 48,405 (24.27) | 64,890 (24.02) | 126,005 (23.71) | 6,450 (21.52) | 6,730 (22.23) |
| 2 | 42,575 (19.96) | 88,600 (21.83) | 1,960 (20.16) | 3,685 (21.94) | 190,670 (20.18) | 136,675 (21.45) | 15,880 (19.07) | 10,200 (20.65) | 230,210 (20.21) | 347,010 (20.62) | 31,110 (19.53) | 40,465 (20.29) | 55,075 (20.38) | 111,285 (20.94) | 6,005 (20.04) | 6,250 (20.65) |
| 3 | 44,160 (20.70) | 80,040 (19.72) | 1,965 (20.22) | 3,345 (19.93) | 192,290 (20.35) | 127,720 (20.05) | 17,685 (21.23) | 10,430 (21.12) | 232,040 (20.38) | 336,915 (20.02) | 33,380 (20.96) | 40,455 (20.29) | 53,520 (19.81) | 105,995 (19.94) | 6,075 (20.28) | 6,210 (20.52) |
| 4 | 40,555 (19.01) | 68,890 (16.98) | 1,830 (18.80) | 2,715 (16.18) | 166,450 (17.62) | 109,100 (17.12) | 15,540 (18.65) | 8,400 (17.01) | 201,340 (17.68) | 301,925 (17.94) | 29,645 (18.61) | 36,210 (18.16) | 48,055 (17.78) | 95,955 (18.05) | 5,590 (18.65) | 5,605 (18.51) |
| 5 (most affluent) | 36,755 (17.23) | 59,110 (14.57) | 1,755 (18.05) | 2,305 (13.73) | 142,045 (15.03) | 92,385 (14.50) | 14,405 (17.29) | 7,390 (14.96) | 172,735 (15.17) | 256,660 (15.25) | 26,400 (16.58) | 30,445 (15.27) | 44,170 (16.35) | 83,865 (15.78) | 5,300 (17.68) | 4,985 (16.48) |
| Unknown | 3,135 (1.47) | 6,780 (1.67) | 150 (1.55) | 295 (1.75) | 15,375 (1.63) | 10,060 (1.58) | 1,330 (1.60) | 830 (1.68) | 18,550 (1.63) | 27,665 (1.64) | 2,570 (1.61) | 3,455 (1.73) | 4,505 (1.67) | 8,400 (1.58) | 550 (1.83) | 490 (1.62) |
| **Season, N (%)** |  |  |  |  |  |  |  |  |  |  |  |  |  |  |  |  |
| Spring | 50,435 (23.64) | 98,975 (24.39) | 2,480 (25.49) | 4,255 (25.34) | 253,450 (26.82) | 166,190 (26.09) | 24,145 (28.99) | 13,730 (27.80) | 303,520 (26.65) | 414,675 (24.64) | 45,220 (28.39) | 51,890 (26.02) | 74,155 (27.44) | 146,745 (27.61) | 8,785 (29.32) | 8,945 (29.55) |
| Summer | 30,420 (14.26) | 69,480 (17.12) | 1,300 (13.39) | 2,525 (15.03) | 222,620 (23.56) | 118,975 (18.67) | 18,525 (22.24) | 8,755 (17.73) | 257,910 (22.65) | 304,515 (18.09) | 31,540 (19.80) | 33,885 (16.99) | 61,155 (22.63) | 126,065 (23.72) | 7,225 (24.11) | 7,015 (23.18) |
| Autumn | 45,565 (21.36) | 87,570 (21.58) | 1,895 (19.49) | 3,185 (18.98) | 194,015 (20.53) | 138,835 (21.79) | 16,105 (19.34) | 9,820 (19.89) | 233,060 (20.47) | 367,655 (21.84) | 30,430 (19.11) | 39,780 (19.95) | 53,225 (19.70) | 101,615 (19.12) | 5,745 (19.17) | 5,330 (17.61) |
| Winter | 86,925 (40.74) | 149,795 (36.91) | 4,045 (41.63) | 6,825 (40.66) | 274,775 (29.08) | 213,095 (33.45) | 24,525 (29.44) | 17,085 (34.59) | 344,325 (30.24) | 596,240 (35.43) | 52,070 (32.70) | 73,885 (37.05) | 81,675 (30.23) | 157,080 (29.55) | 8,210 (27.40) | 8,980 (29.66) |
| **Region, N (%)** |  |  |  |  |  |  |  |  |  |  |  |  |  |  |  |  |
| London | 13,135 (6.16) | 29,825 (7.35) | 580 (5.95) | 890 (5.31) | 30,180 (3.19) | 30,210 (4.74) | 3,250 (3.90) | 1,655 (3.35) | 44,680 (3.92) | 59,385 (3.53) | 8,090 (5.08) | 6,510 (3.27) | 15,185 (5.62) | 26,855 (5.05) | 1,495 (4.99) | 1,280 (4.23) |
| North East | 9,875 (4.63) | 16,465 (4.06) | 375 (3.84) | 645 (3.85) | 66,390 (7.03) | 35,980 (5.65) | 5,215 (6.26) | 2,905 (5.88) | 77,490 (6.80) | 92,320 (5.49) | 10,185 (6.40) | 11,385 (5.71) | 14,810 (5.48) | 26,340 (4.96) | 1,525 (5.10) | 1,380 (4.56) |
| North West | 24,375 (11.43) | 37,155 (9.16) | 1,000 (10.31) | 1,610 (9.58) | 119,025 (12.60) | 70,615 (11.08) | 10,885 (13.07) | 6,345 (12.85) | 138,155 (12.13) | 190,575 (11.32) | 20,355 (12.78) | 25,710 (12.89) | 29,290 (10.84) | 53,015 (9.97) | 3,210 (10.72) | 3,250 (10.73) |
| East | 46,510 (21.80) | 107,130 (26.40) | 1,950 (20.08) | 3,940 (23.48) | 202,995 (21.48) | 156,500 (24.56) | 17,460 (20.96) | 11,625 (23.54) | 246,000 (21.60) | 391,775 (23.28) | 33,170 (20.83) | 44,175 (22.15) | 63,280 (23.42) | 136,010 (25.59) | 6,825 (22.78) | 8,120 (26.83) |
| West Midlands | 9,815 (4.60) | 24,390 (6.01) | 525 (5.39) | 1,070 (6.37) | 33,690 (3.57) | 31,760 (4.98) | 3,345 (4.01) | 2,125 (4.30) | 42,220 (3.71) | 80,720 (4.80) | 6,270 (3.94) | 8,345 (4.18) | 13,595 (5.03) | 26,645 (5.01) | 1,310 (4.38) | 1,460 (4.82) |
| Yorkshire and The Humber | 32,650 (15.30) | 78,405 (19.32) | 2,285 (23.51) | 3,495 (20.83) | 155,250 (16.43) | 99,705 (15.65) | 14,280 (17.14) | 7,935 (16.06) | 186,320 (16.36) | 298,520 (17.74) | 27,630 (17.35) | 36,210 (18.16) | 42,365 (15.68) | 87,455 (16.45) | 5,090 (16.99) | 4,685 (15.49) |
| South East | 14,190 (6.65) | 19,390 (4.78) | 575 (5.89) | 945 (5.62) | 57,960 (6.13) | 39,020 (6.12) | 5,275 (6.33) | 3,270 (6.62) | 70,310 (6.17) | 95,555 (5.68) | 9,435 (5.92) | 11,070 (5.55) | 16,205 (6.00) | 28,470 (5.36) | 1,980 (6.60) | 1,730 (5.71) |
| East Midlands | 39,850 (18.68) | 66,730 (16.44) | 1,700 (17.51) | 3,360 (20.00) | 165,210 (17.49) | 112,025 (17.58) | 15,065 (18.09) | 8,830 (17.88) | 197,075 (17.31) | 313,310 (18.62) | 28,760 (18.06) | 39,040 (19.57) | 48,415 (17.92) | 97,565 (18.36) | 5,395 (18.00) | 5,675 (18.76) |
| South West | 22,945 (10.75) | 26,335 (6.49) | 730 (7.53) | 835 (4.96) | 114,160 (12.08) | 61,280 (9.62) | 8,525 (10.23) | 4,705 (9.52) | 136,565 (11.99) | 160,940 (9.56) | 15,365 (9.65) | 16,995 (8.52) | 27,070 (10.02) | 49,150 (9.25) | 3,130 (10.45) | 2,685 (8.87) |
| **Flu vaccination, N (%)** |  |  |  |  |  |  |  |  |  |  |  |  |  |  |  |  |
| Yes | 66,855 (31.34) | 166,040 (40.91) | 3,865 (39.74) | 8,125 (48.39) | 512,820 (54.27) | 308,975 (48.50) | 42,275 (50.75) | 27,000 (54.67) | 597,265 (52.45) | 824,215 (48.97) | 83,245 (52.27) | 113,020 (56.67) | 60,975 (22.56) | 93,945 (17.68) | 6,105 (20.37) | 5,395 (17.83) |
| No | 146,490 (68.66) | 239,785 (59.09) | 5,860 (60.26) | 8,665 (51.61) | 432,045 (45.73) | 328,115 (51.50) | 41,025 (49.25) | 22,385 (45.33) | 541,550 (47.55) | 858,880 (51.03) | 76,015 (47.73) | 86,420 (43.33) | 209,240 (77.44) | 437,560 (82.32) | 23,865 (79.63) | 24,870 (82.17) |
| **Period** |  |  |  |  |  |  |  |  |  |  |  |  |  |  |  |  |
| Pre-pandemic | 122,030 (57.20) | 191,855 (47.27) | 5,240 (53.91) | 8,440 (50.28) | 333,000 (35.24) | 248,725 (39.04) | 31,170 (37.42) | 20,070 (40.64) | 416,280 (36.55) | 752,035 (44.68) | 66,655 (41.85) | 94,995 (47.63) | 119,605 (44.26) | 221,370 (41.65) | 12,245 (40.86) | 12,855 (42.47) |
| During pandemic | 45,695 (21.42) | 85,355 (21.03) | 2,355 (24.20) | 3,975 (23.68) | 275,730 (29.18) | 156,890 (24.63) | 25,430 (30.53) | 13,855 (28.05) | 329,000 (28.89) | 380,930 (22.63) | 46,735 (29.34) | 51,485 (25.81) | 72,575 (26.86) | 137,230 (25.82) | 9,105 (30.38) | 8,695 (28.72) |
| After 2^nd^ lockdown | 45,620 (21.38) | 128,615 (31.69) | 2,130 (21.89) | 4,370 (26.04) | 336,130 (35.57) | 231,485 (36.33) | 26,705 (32.06) | 15,460 (31.31) | 393,535 (34.56) | 550,130 (32.69) | 45,870 (28.80) | 52,960 (26.55) | 78,035 (28.88) | 172,910 (32.53) | 8,620 (28.76) | 8,720 (28.81) |
| **Count of antibiotic prescription, mean (SD^5^)** | 1.00 (1.62) | 2.35 (1.81) | 2.05 (2.01) | 3.60 (2.02) | 1.57 (2.29) | 2.80 (2.39) | 2.26 (2.43) | 3.97 (2.66) | 1.55  (2.30) | 2.73 (2.32) | 2.59 (2.54) | 4.17 (2.60) | 0.87 (1.34) | 1.89 (1.26) | 1.69 (1.51) | 2.76 (1.38) |
| ^1^ ABs, antibiotics prescribed or not.  ^2^ BMI, Body Mass Index recorded in the last 5 years.  ^3^ CCI, Charlson Comorbidities Index, measured from 17 weighted conditions, including myocardial infarction, congestive heart failure, peripheral vascular disease, cerebrovascular disease, dementia, chronic pulmonary disease, Connective tissue disease, ulcer disease, mild liver disease, diabetes, hemiplegia, moderate or severe renal disease, diabetes with complications, any malignancy (including leukaemia and lymphoma), moderate or severe liver disease, metastatic solid tumour, and AIDS.  ^4^ IMD, Multiple Deprivation Index, quintile measured from patient-level address.  ^5^ SD, standard deviation. | | | | | | | | | | | | | | | | |

#### Counts and rates of hospital admission cases

Supplementary Table 5. Further counts and rates of hospital admission of patients with incident common infections, including lower respiratory tract infection (LRTI), upper respiratory tract infection (URTI), and urinary tract infection (UTI), with no prescribed antibiotics.

|  | **Incident LRTI**  **with no ABs^1^** | **Incident URTI**  **with no ABs** | **Incident UTI**  **with no ABs** |
| --- | --- | --- | --- |
| **BMI^2^, N (rate)** |  |  |  |
| Underweight | 625 (85.7) | 1,335 (23.5) | 425 (64.4) |
| Healthy weight | 4,605 (74.1) | 9,930 (15.9) | 4,475 (57.1) |
| Overweight | 4,655 (70.6) | 10,360 (14.6) | 4,850 (63.9) |
| Obese | 5,010 (71.4) | 11,730 (16.1) | 5,170 (70.1) |
| Unknown | 3,020 (63.8) | 6,680 (14.9) | 3,215 (53.8) |
| **Smoking status, N (rate)** |  |  |  |
| Smoker | 2,515 (59.1) | 6,665 (12.0) | 1,825 (51.4) |
| Ex-smoker | 10,285 (80.9) | 22,145 (18.2) | 9,960 (74.1) |
| Never smoked | 5,075 (61.7) | 11,075 (14.1) | 6,305 (51.3) |
| Unknown | 40 (39.0) | 150 (11.4) | 45 (30.1) |
| **IMD^3^, N (rate)** |  |  |  |
| 1 (most deprived) | 4,210 (70.1) | 9,935 (15.7) | 3,830 (63.7) |
| 2 | 3,580 (71.6) | 8,205 (15.8) | 3,715 (65.0) |
| 3 | 3,620 (70.3) | 8,130 (15.6) | 3,825 (62.2) |
| 4 | 3,400 (73.6) | 7,275 (15.9) | 3,545 (61.9) |
| 5 (most affluent) | 2,780 (67.3) | 5,860 (14.8) | 2,890 (54.1) |
| Unknown | 325 (80.5) | 625 (15.0) | 330 (69.0) |
| **Season, N (rate)** |  |  |  |
| Spring | 4,335 (66.7) | 10,065 (14.8) | 4,600 (60.2) |
| Summer | 4,125 (78.2) | 9,180 (16.0) | 5,110 (62.6) |
| Autumn | 3,710 (69.8) | 8,335 (15.9) | 3,920 (61.9) |
| Winter | 5,745 (70.0) | 12,460 (15.8) | 4,510 (61.8) |
| **Region, N (rate)** |  |  |  |
| London | 470 (48.2) | 1,200 (11.6) | 585 (46.2) |
| North East | 1,190 (75.7) | 2,780 (16.5) | 1,060 (64.9) |
| North West | 1,670 (58.0) | 4,035 (13.0) | 1,565 (49.2) |
| East | 4,075 (70.2) | 9,050 (16.2) | 4,395 (63.8) |
| West Midlands | 830 (73.6) | 1,840 (18.5) | 760 (65.0) |
| Yorkshire and The Humber | 2,960 (72.3) | 6,440 (15.5) | 3,005 (69.5) |
| South East | 1,165 (73.1) | 2,470 (15.6) | 1,245 (60.5) |
| East Midlands | 3,475 (80.5) | 7,915 (17.6) | 3,330 (66.7) |
| South West | 2,085 (71.1) | 4,310 (14.3) | 2,190 (55.9) |
| ^1^ ABs, antibiotics prescribed or not.  ^2^ BMI, Body Mass Index recorded in the last 5 years.  ^3^ IMD, Multiple Deprivation Index, quintile measured from patient-level address. | | | |

Supplementary Table 6. Counts and rates of hospital admission of patients with incident common infections with prescribed antibiotics, prevalent common infections with no prescribed antibiotics, and prevalent common infections with prescribed antibiotics. Common infections include lower respiratory tract infection (LRTI), upper respiratory tract infection (URTI), and urinary tract infection (UTI).

|  | **LRTI** | | | **URTI** | | | **UTI** | | |
| --- | --- | --- | --- | --- | --- | --- | --- | --- | --- |
|  | **Incident** | **Prevalent** | | **Incident** | **Prevalent** | | **Incident** | **Prevalent** | |
|  | **With ABs^1^** | **No ABs** | **With ABs** | **With ABs** | **No ABs** | **With ABs** | **With ABs** | **No ABs** | **With ABs** |
| **Total, N cases** | 35,890 | 4,270 | 4,270 | 58,320 | 7,620 | 8,885 | 47,570 | 5,380 | 9,790 |
| **Age, N (rate^2^)** |  |  |  |  |  |  |  |  |  |
| 18-24 | 530 (8.5) | 60 (43.8) | 60 (20.6) | 2,640 (10.6) | 435 (28.2) | 360 (23.4) | 1,010 (7.8) | 100 (22.4) | 185 (15.6) |
| 25-34 | 1,320 (9.0) | 115 (32.7) | 115 (12.8) | 3,800 (8.9) | 500 (18.8) | 445 (16.6) | 1,790 (8.2) | 180 (28.1) | 290 (14.9) |
| 35-44 | 1,555 (8.7) | 155 (31.5) | 155 (11.2) | 3,385 (8.1) | 360 (12.2) | 390 (13.0) | 1,755 (8.4) | 160 (27.9) | 350 (17.4) |
| 45-54 | 2,735 (10.8) | 280 (36.7) | 280 (12.4) | 4,930 (9.8) | 650 (15.0) | 715 (16.9) | 2,850 (10.9) | 280 (36.4) | 555 (18.7) |
| 55-64 | 4,320 (14.4) | 450 (46.6) | 450 (14.8) | 7,125 (12.7) | 890 (16.5) | 1,100 (20.5) | 4,560 (16.0) | 475 (46.7) | 940 (24.2) |
| 65-74 | 7,245 (23.4) | 940 (78.8) | 940 (26.1) | 11,145 (20.1) | 1,540 (27.0) | 1,915 (31.9) | 9,340 (25.8) | 1,065 (68.7) | 2,025 (33.8) |
| 75+ | 18,185 (52.3) | 2,270 (110.9) | 2,270 (48.6) | 25,300 (46.0) | 3,245 (57.1) | 3,955 (58.5) | 26,265 (56.0) | 3,120 (114.3) | 5,455 (63.2) |
| **Sex, N (rate)** |  |  |  |  |  |  |  |  |  |
| Male | 15,765 (24.9) | 1,955 (82.5) | 1,955 (31.5) | 25,320 (20.5) | 3,435 (29.9) | 3,900 (35.1) | 18,065 (54.6) | 2,320 (105.0) | 3,465 (65.5) |
| Female | 20,125 (20.9) | 2,315 (64.7) | 2,315 (23.3) | 33,000 (16.3) | 4,185 (25.0) | 4,985 (27.0) | 29,505 (18.4) | 3,060 (55.5) | 6,325 (29.6) |
| **BMI^3^, N (rate)** |  |  |  |  |  |  |  |  |  |
| Underweight | 1,195 (40.4) | 170 (103.7) | 170 (48.9) | 1,795 (30.4) | 240 (44.4) | 280 (46.9) | 1,160 (31.7) | 120 (74.3) | 200 (40.1) |
| Healthy weight | 8,750 (26.2) | 1,140 (77.7) | 1,140 (32.8) | 13,855 (20.0) | 1,830 (28.4) | 2,175 (34.1) | 11,690 (22.9) | 1,275 (61.5) | 2,380 (34.4) |
| Overweight | 9,545 (22.4) | 1,175 (74.1) | 1,175 (26.9) | 15,050 (17.9) | 1,975 (25.3) | 2,305 (29.0) | 12,860 (25.3) | 1,470 (69.8) | 2,795 (38.0) |
| Obese | 10,570 (21.0) | 1,125 (64.8) | 1,125 (20.7) | 17,280 (17.5) | 2,225 (26.3) | 2,740 (28.3) | 13,560 (27.2) | 1,635 (82.1) | 2,810 (39.1) |
| Unknown | 5,830 (19.2) | 660 (66.2) | 660 (26.2) | 10,340 (15.3) | 1,350 (27.1) | 1,385 (27.8) | 8,295 (21.9) | 880 (63.2) | 1,610 (34.4) |
| **Ethnicity, N (rate)** |  |  |  |  |  |  |  |  |  |
| White | 20,175 (21.9) | 2,505 (68.2) | 2,505 (25.6) | 32,445 (17.8) | 4,365 (25.2) | 5,140 (29.1) | 27,055 (24.0) | 3,100 (65.7) | 5,785 (35.6) |
| Non-White | 1,770 (15.6) | 175 (59.1) | 175 (21.5) | 3,330 (12.0) | 400 (18.8) | 365 (18.8) | 1,910 (16.1) | 210 (57.6) | 355 (30.5) |
| Unknown | 13,945 (24.7) | 1,590 (80.3) | 1,590 (28.6) | 22,550 (19.5) | 2,855 (32.6) | 3,375 (33.7) | 18,605 (27.1) | 2,070 (78.4) | 3,650 (39.6) |
| **CCI^4^, N (rate)** |  |  |  |  |  |  |  |  |  |
| Very low | 13,125 (15.3) | 1,335 (50.4) | 1,335 (18.5) | 23,960 (12.6) | 2,930 (18.5) | 3,145 (21.2) | 19,785 (16.2) | 1,990 (47.3) | 3,830 (25.5) |
| Low | 15,105 (25.9) | 1,875 (78.2) | 1,875 (27.8) | 23,070 (21.3) | 3,115 (32.8) | 3,700 (32.8) | 18,125 (33.4) | 2,075 (84.3) | 3,825 (45.3) |
| Medium | 5,670 (44.7) | 785 (112.7) | 785 (45.1) | 8,415 (38.3) | 1,185 (52.4) | 1,495 (54.1) | 7,185 (53.6) | 940 (117.9) | 1,540 (62.4) |
| High | 1,510 (63.8) | 195 (126.2) | 195 (56.5) | 2,135 (52.9) | 290 (63.7) | 410 (73.9) | 1,895 (72.1) | 285 (144.7) | 455 (85.0) |
| Very high | 480 (68.0) | 75 (138.9) | 75 (71.4) | 740 (62.1) | 100 (70.4) | 140 (87.2) | 580 (79.7) | 90 (145.2) | 145 (87.9) |
| **Smoking status, N (rate)** |  |  |  |  |  |  |  |  |  |
| Smoker | 4,640 (14.6) | 460 (54.0) | 460 (15.9) | 8,535 (13.1) | 1,030 (21.8) | 1,160 (21.0) | 4,760 (21.1) | 480 (63.7) | 905 (34.6) |
| Ex-smoker | 20,445 (27.3) | 2,605 (82.8) | 2,605 (31.2) | 31,725 (22.0) | 4,365 (32.3) | 5,180 (35.9) | 25,935 (29.8) | 3,040 (81.0) | 5,410 (41.7) |
| Never smoked | 10,730 (20.4) | 1,195 (61.6) | 1,195 (24.5) | 17,795 (15.5) | 2,190 (22.2) | 2,510 (26.3) | 16,765 (20.2) | 1,855 (58.1) | 3,475 (31.6) |
| Unknown | 75 (15.0) | 10 (76.9) | 10 (44.4) | 270 (12.8) | 40 (38.5) | 35 (34.3) | 110 (17.2) | SN^5^ | 10 (17.7) |
| **IMD^6^, N (rate)** |  |  |  |  |  |  |  |  |  |
| 1 (most deprived) | 8,305 (21.0) | 940 (73.2) | 940 (23.7) | 13,835 (17.2) | 1,815 (28.8) | 2,060 (28.7) | 9,485 (25.5) | 1,105 (81.1) | 1,865 (39.7) |
| 2 | 7,110 (21.9) | 820 (72.2) | 820 (25.3) | 11,875 (17.4) | 1,520 (27.7) | 1,785 (29.5) | 9,190 (25.3) | 1,055 (75.0) | 1,925 (40.1) |
| 3 | 7,375 (23.2) | 880 (71.5) | 880 (27.4) | 11,840 (18.2) | 1,530 (25.9) | 1,875 (31.0) | 10,150 (25.2) | 1,175 (71.8) | 2,095 (37.3) |
| 4 | 6,865 (23.8) | 825 (71.3) | 825 (27.8) | 10,990 (19.1) | 1,475 (28.0) | 1,650 (31.2) | 9,535 (24.3) | 1,030 (63.6) | 2,010 (35.9) |
| 5 (most affluent) | 5,640 (23.0) | 725 (69.2) | 725 (29.2) | 8,840 (18.0) | 1,140 (23.8) | 1,360 (30.1) | 8,450 (22.6) | 925 (58.6) | 1,745 (31.7) |
| Unknown | 595 (21.8) | 80 (86.0) | 80 (28.3) | 940 (17.8) | 140 (30.5) | 150 (29.6) | 760 (25.1) | 95 (77.2) | 150 (34.8) |
| **Season, N (rate)** |  |  |  |  |  |  |  |  |  |
| Spring | 8,385 (21.9) | 1,005 (66.8) | 1,005 (25.9) | 14,190 (17.2) | 1,940 (24.1) | 2,135 (27.1) | 12,470 (24.9) | 1,390 (69.8) | 2,530 (36.4) |
| Summer | 6,855 (24.5) | 865 (81.1) | 865 (32.6) | 11,395 (18.4) | 1,600 (27.3) | 1,630 (31.2) | 13,210 (24.8) | 1,505 (72.6) | 2,680 (37.6) |
| Autumn | 7,825 (22.4) | 825 (70.9) | 825 (25.5) | 12,600 (18.1) | 1,500 (27.7) | 1,810 (31.1) | 10,205 (24.3) | 1,150 (66.9) | 2,170 (37.5) |
| Winter | 12,825 (21.9) | 1,575 (71.1) | 1,575 (24.7) | 20,140 (18.0) | 2,580 (29.0) | 3,310 (31.0) | 11,685 (24.3) | 1,340 (69.0) | 2,420 (35.7) |
| **Region, N (rate)** |  |  |  |  |  |  |  |  |  |
| London | 845 (17.5) | 100 (63.7) | 100 (35.0) | 1,880 (12.9) | 255 (19.0) | 190 (18.4) | 1,320 (22.0) | 190 (70.2) | 185 (31.5) |
| North East | 1,925 (22.2) | 280 (76.7) | 280 (31.8) | 3,055 (17.9) | 500 (28.9) | 470 (28.8) | 2,200 (25.7) | 280 (74.2) | 470 (40.4) |
| North West | 3,990 (22.4) | 480 (63.2) | 480 (23.4) | 6,455 (18.4) | 810 (22.8) | 1,060 (28.7) | 5,100 (24.3) | 535 (60.1) | 1,155 (35.3) |
| East | 8,025 (21.8) | 925 (72.7) | 925 (26.0) | 13,335 (16.8) | 1,690 (28.4) | 1,950 (28.7) | 11,550 (25.0) | 1,260 (74.9) | 2,235 (36.4) |
| West Midlands | 1,700 (22.6) | 185 (74.6) | 185 (26.9) | 2,930 (17.9) | 355 (31.0) | 405 (31.2) | 2,030 (25.0) | 240 (85.4) | 415 (39.2) |
| Yorkshire and The Humber | 6,480 (21.0) | 805 (70.0) | 805 (24.4) | 9,960 (17.7) | 1,350 (27.4) | 1,555 (29.7) | 7,945 (25.4) | 1,000 (70.1) | 1,840 (40.4) |
| South East | 2,040 (23.9) | 245 (73.7) | 245 (30.4) | 3,460 (19.0) | 480 (27.8) | 560 (32.9) | 2,915 (24.5) | 290 (59.1) | 550 (35.0) |
| East Midlands | 7,170 (23.8) | 830 (76.9) | 830 (24.7) | 11,270 (19.1) | 1,485 (29.2) | 1,855 (32.6) | 9,255 (24.3) | 995 (71.4) | 2,040 (36.3) |
| South West | 3,720 (25.6) | 420 (71.6) | 420 (34.4) | 5,985 (20.1) | 695 (25.0) | 845 (33.5) | 5,250 (23.6) | 590 (64.8) | 900 (33.8) |
| **Flu vaccination, N (rate)** |  |  |  |  |  |  |  |  |  |
| Yes | 23,865 (30.7) | 2,925 (86.0) | 2,925 (31.3) | 35,715 (25.6) | 4,645 (34.3) | 5,815 (37.9) | 31,140 (35.6) | 3,600 (87.0) | 6,670 (45.6) |
| No | 12,025 (14.6) | 1,345 (52.8) | 1,345 (19.8) | 22,610 (12.1) | 2,975 (20.3) | 3,070 (21.6) | 16,425 (15.5) | 1,780 (49.6) | 3,125 (26.0) |
| **Period** |  |  |  |  |  |  |  |  |  |
| Pre-pandemic | 18,865 (24.6) | 2,140 (72.0) | 2,140 (24.9) | 28,740 (20.3) | 3,595 (31.2) | 4,660 (34.2) | 16,395 (27.4) | 1,995 (77.9) | 3,615 (41.9) |
| During pandemic | 7,905 (23.1) | 1,110 (72.9) | 1,110 (28.9) | 13,710 (18.0) | 1,970 (23.6) | 2,205 (28.3) | 14,615 (22.0) | 1,580 (61.9) | 2,970 (32.3) |
| After 2^nd^ lockdown | 9,125 (18.7) | 1,020 (70.1) | 1,020 (27.5) | 15,870 (14.7) | 2,055 (24.7) | 2,020 (24.8) | 16,555 (24.7) | 1,805 (69.2) | 3,210 (36.3) |
| ^1^ ABs, antibiotics prescribed or not.  ^2^ Rate is the number of cases per 1000 patients with common infection, calculated by dividing the count of infection-related hospital admission cases (numerator) by the count of infection diagnosis (denominator) and then multiplied by 1000.  ^3^ BMI, Body Mass Index recorded in the last 5 years.  ^4^ CCI, Charlson Comorbidities Index, measured from 17 weighted conditions, including myocardial infarction, congestive heart failure, peripheral vascular disease, cerebrovascular disease, dementia, chronic pulmonary disease, Connective tissue disease, ulcer disease, mild liver disease, diabetes, hemiplegia, moderate or severe renal disease, diabetes with complications, any malignancy (including leukaemia and lymphoma), moderate or severe liver disease, metastatic solid tumour, and AIDS.  ^5^ SN, small numbers, equal or less than 5.  ^6^ IMD, Multiple Deprivation Index, quintile measured from patient-level address. | | | | | | | | | |

Supplementary Table 7. Counts and rate of hospital admission of patients with other common infections, including sinusitis, otitis media, and otitis externa. The other common infections include incident infections with no prescribed antibiotics, incident infections with prescribed antibiotics, prevalent infections with no prescribed antibiotics, and prevalent infections with prescribed antibiotics.

|  | **Sinusitis** | | | | **Otitis media** | | | | **Otitis externa** | | | |
| --- | --- | --- | --- | --- | --- | --- | --- | --- | --- | --- | --- | --- |
|  | **Incident** | | **Prevalent** | | **Incident** | | **Prevalent** | | **Incident** | | **Prevalent** | |
|  | **No** **ABs^1^** | **With ABs** | **No ABs** | **With ABs** | **No ABs** | **With ABs** | **No ABs** | **With ABs** | **No ABs** | **With ABs** | **No ABs** | **With ABs** |
| **Total, N cases** | 535 | 1,205 | 75 | 130 | 755 | 1,545 | 155 | 170 | 3,015 | 1,660 | 1,010 | 465 |
| **Age, N (rate^2^)** |  |  |  |  |  |  |  |  |  |  |  |  |
| 18-24 | 40 (4.3) | 75 (3.9) | SN^3^ | SN | 50 (6.0) | 130 (5.4) | 10 (11.6) | 15 (11.5) | 205 (4.2) | 115 (8.7) | 45 (10.3) | 25 (14.8) |
| 25-34 | 75 (3.1) | 175 (2.9) | 15 (6.5) | 15 (3.9) | 115 (7.6) | 290 (6.1) | 20 (11.0) | 35 (10.9) | 375 (4.1) | 255 (9.3) | 75 (9.0) | 45 (11.8) |
| 35-44 | 80 (3.2) | 175 (2.4) | 10 (3.5) | 25 (4.6) | 95 (7.2) | 225 (5.2) | 20 (11.8) | 25  (8.3) | 315 (3.4) | 210 (7.7) | 105 (12.7) | 45 (11.7) |
| 45-54 | 80 (3.1) | 185 (2.4) | 15 (4.8) | 25 (4.2) | 95 (6.8) | 225 (5.4) | 25 (13.6) | 20  (6.7) | 395 (3.7) | 245 (8.0) | 105 (10.4) | 40 (9.3) |
| 55-64 | 85 (3.9) | 220 (3.3) | 15 (5.8) | 15 (3.0) | 110 (8.8) | 190 (5.6) | 10 (6.4) | 20  (8.4) | 365 (3.7) | 220 (8.4) | 105 (11.0) | 60 (15.1) |
| 65-74 | 70 (4.3) | 195 (4.1) | 10 (5.3) | 20 (5.5) | 120 (11.5) | 205 (8.3) | 20 (16.3) | 20 (11.3) | 515 (5.8) | 245 (12.2) | 155 (16.8) | 80 (23.8) |
| 75+ | 105 (14.5) | 185 (10.0) | 15 (21.0) | 20 (15.9) | 175 (23.2) | 285 (19.6) | 45 (50.0) | 35 (34.0) | 845 (13.6) | 370 (28.9) | 415 (53.5) | 170 (64.3) |
| **Sex, N (rate)** |  |  |  |  |  |  |  |  |  |  |  |  |
| Male | 205 (4.5) | 350 (3.6) | 25 (5.6) | 35 (5.2) | 350 (10.6) | 535 (6.7) | 60 (15.4) | 60 (10.5) | 1,305 (5.3) | 640 (10.9) | 540 (22.2) | 225 (24.1) |
| Female | 330 (3.9) | 855 (3.2) | 50 (5.1) | 95 (4.9) | 405 (8.4) | 1,010 (6.8) | 95 (15.8) | 110 (11.0) | 1,710 (4.9) | 1,020 (10.3) | 470 (14.1) | 240 (16.8) |
| **BMI^4^, N (rate)** |  |  |  |  |  |  |  |  |  |  |  |  |
| Underweight | 15 (8.6) | 15 (3.2) | - | SN | 15 (14.2) | 20  (7.0) | SN | SN | 45 (6.2) | 15 (8.3) | SN | SN |
| Healthy weight | 125 (3.9) | 285 (3.3) | 20 (5.7) | 30 (4.9) | 155 (9.0) | 300 (6.7) | 25 (12.4) | 30 (10.6) | 585 (4.8) | 320 (10.8) | 195 (16.6) | 80 (17.9) |
| Overweight | 130 (3.8) | 295 (3.2) | 20 (5.3) | 30 (4.4) | 210 (10.6) | 365 (6.6) | 35 (14.4) | 45 (11.7) | 785 (5.2) | 440 (11.5) | 325 (21.1) | 165 (27.3) |
| Obese | 150 (4.9) | 385 (4.0) | 25 (7.1) | 45 (6.2) | 215 (10.0) | 550 (8.3) | 50 (17.7) | 65 (13.5) | 1,035 (6.4) | 595 (12.2) | 345 (20.6) | 150 (20.1) |
| Unknown | 115 (3.7) | 225 (2.7) | 15 (78.9) | 25 (4.5) | 165 (7.8) | 310 (5.2) | 35 (13.8) | 30  (7.5) | 565 (3.8) | 290 (7.4) | 135 (10.3) | 60 (11.1) |
| **Ethnicity, N (rate)** |  |  |  |  |  |  |  |  |  |  |  |  |
| White | 290 (3.8) | 705 (3.3) | 55 (12.9) | 90 (5.5) | 455 (10.0) | 865 (6.8) | 80 (14.0) | 105 (11.8) | 1,750 (5.2) | 920 (10.1) | 585 (16.9) | 270 (19.3) |
| Non-White | 50 (4.9) | 85 (3.6) | SN | SN | 55 (7.8) | 135 (6.8) | 20 (23.8) | 10  (8.1) | 225 (4.9) | 105 (9.5) | 60 (15.6) | 25 (18.1) |
| Unknown | 200 (4.7) | 420 (3.4) | 20 (22.3) | 35 (4.1) | 250 (8.9) | 550 (6.7) | 50 (14.9) | 55 (9.9) | 1,040 (5.1) | 630 (11.3) | 370 (19.2) | 165 (20.0) |
| **CCI^5^, N (rate)** |  |  |  |  |  |  |  |  |  |  |  |  |
| Very low | 305 (3.2) | 655 (2.5) | 45 (4.5) | 65 (3.6) | 415 (7.1) | 920 (5.5) | 85 (12.0) | 90  (8.0) | 1,540 (3.6) | 925 (8.3) | 440 (11.0) | 195 (12.1) |
| Low | 175 (5.6) | 415 (4.6) | 25 (7.0) | 55 (7.6) | 245 (13.3) | 445 (8.5) | 40 (17.4) | 50 (13.1) | 985 (7.3) | 490 (13.0) | 335 (23.9) | 165 (28.0) |
| Medium | 40 (10.7) | 105 (9.0) | SN | SN | 75 (24.4) | 140 (18.0) | 20 (50.0) | 20 (36.0) | 350 (14.5) | 175 (26.5) | 170 (57.0) | 75 (58.8) |
| High | 10 (16.3) | 20 (11.9) | SN | SN | 20 (37.0) | 30 (23.1) | SN | SN | 95 (23.1) | 55 (48.0) | 60 (95.2) | 15 (60.0) |
| Very high | SN | 10 (19.8) | SN | SN | SN | 10 (29.0) | SN | SN | 40 (36.9) | 15 (50.8) | 15 (103.4) | 10 (222.2) |
| **Smoking status, N (rate)** |  |  |  |  |  |  |  |  |  |  |  |  |
| Smoker | 90 (4.7) | 195 (3.4) | 10 (1.7) | 20 (4.8) | 155 (10.6) | 295 (6.5) | 25 (14.3) | 30 (10.2) | 495 (5.1) | 295 (9.2) | 120 (13.1) | 70 (16.6) |
| Ex-smoker | 245 (4.5) | 550 (3.6) | 40 (4.4) | 55 (4.8) | 325 (10.1) | 660 (7.5) | 65 (16.2) | 80 (12.5) | 1,460 (5.9) | 765 (11.8) | 540 (21.6) | 245 (23.8) |
| Never smoked | 195 (3.5) | 460 (3.1) | 30 (4.9) | 55 (5.2) | 275 (8.2) | 585 (6.3) | 60 (14.7) | 60  (9.6) | 1,045 (4.3) | 600 (10.1) | 345 (14.9) | 150 (16.6) |
| Unknown | SN | SN | - | - | SN | SN | SN | SN | 10 (1.9) | SN | SN | - |
| **IMD^6^, N (rate)** |  |  |  |  |  |  |  |  |  |  |  |  |
| 1 (most deprived) | 120 (4.8) | 235 (3.6) | 10 (43.5) | 20 (4.3) | 180 (9.5) | 415 (7.2) | 30 (13.3) | 55 (13.8) | 690 (5.6) | 400 (11.0) | 225 (19.8) | 100 (20.4) |
| 2 | 100 (4.0) | 230 (3.3) | 15 (5.9) | 25 (5.0) | 150 (9.1) | 330 (6.9) | 35 (18.6) | 40 (12.2) | 580 (5.1) | 350 (10.9) | 175 (16.7) | 95 (21.2) |
| 3 | 105 (3.9) | 280 (3.6) | 20 (7.5) | 35 (6.4) | 135 (8.4) | 275 (6.2) | 40 (20.4) | 35 (12.0) | 635 (5.2) | 320 (10.0) | 190 (16.0) | 85 (17.3) |
| 4 | 100 (3.8) | 225 (3.0) | 15 (4.9) | 25 (4.5) | 150 (10.2) | 250 (6.2) | 20 (10.7) | 20  (7.0) | 550 (4.8) | 300 (10.3) | 230 (19.7) | 85 (17.9) |
| 5 (most affluent) | 100 (4.1) | 215 (3.1) | 20 (7.7) | 30 (5.8) | 130 (9.8) | 240 (7.0) | 25 (14.1) | 20  (8.3) | 505 (4.6) | 270 (10.6) | 175 (15.4) | 85 (20.1) |
| Unknown | 10 (4.6) | 20 (3.4) | SN | SN | 15 (11.1) | 35 (9.2) | SN | SN | 50 (5.4) | 20 (7.9) | 20 (21.9) | 10 (26.0) |
| **Season, N (rate)** |  |  |  |  |  |  |  |  |  |  |  |  |
| Spring | 150 (4.2) | 305 (3.1) | 20 (363.6) | 35 (6.7) | 180 (9.0) | 350 (6.2) | 30 (12.4) | 35  (9.1) | 720 (5.1) | 355 (9.5) | 235 (17.9) | 85 (14.7) |
| Summer | 130 (5.0) | 240 (3.6) | 15 (5.3) | 30 (4.1) | 195 (9.4) | 410 (7.3) | 40 (17.1) | 55 (14.3) | 850 (5.2) | 470 (11.0) | 265 (16.9) | 115 (21.9) |
| Autumn | 105 (3.8) | 255 (3.4) | 20 (9.5) | 30 (461.5) | 175 (10.1) | 375 (7.7) | 35 (16.3) | 40 (12.1) | 725 (5.5) | 415 (11.6) | 290 (20.8) | 120 (923.1) |
| Winter | 150 (3.7) | 410 (3.3) | 25 (6.0) | 35 (7.4) | 210 (9.3) | 410 (6.1) | 45 (15.0) | 35  (7.5) | 720 (4.7) | 420 (10.1) | 225 (15.1) | 140 (21.7) |
| **Region, N (rate)** |  |  |  |  |  |  |  |  |  |  |  |  |
| London | 20 (3.3) | 40 (3.1) | SN | SN | 30 (8.1) | 40  (4.6) | SN | SN | 125 (5.2) | 45 (7.4) | 35 (16.5) | 10 (14.5) |
| North East | 35 (5.0) | 45 (2.8) | 10 (18.3) | SN | 50 (10.5) | 80 (7.0) | 15 (28.6) | 10 (12.3) | 160 (5.1) | 100 (12.3) | 60 (19.1) | 25 (19.2) |
| North West | 60 (3.7) | 135 (3.5) | 10 (13.4) | 15 (4.6) | 80 (8.7) | 170 (6.8) | 10 (8.1) | 15  (8.1) | 335 (5.1) | 155 (9.6) | 120 (17.3) | 50 (19.2) |
| East | 115 (3.9) | 285 (2.9) | 20 (6.2) | 40 (5.7) | 160 (7.5) | 320 (5.3) | 30 (11.8) | 25  (6.0) | 520 (3.9) | 320 (8.0) | 145 (11.6) | 65 (11.3) |
| West Midlands | 25 (5.4) | 55 (4.3) | SN | SN | 35 (9.9) | 70 (6.8) | SN | SN | 110 (5.1) | 65 (11.1) | 25 (14.8) | 10 (13.9) |
| Yorkshire and The Humber | 90 (4.1) | 215 (3.7) | 15 (31.6) | 15 (3.4) | 130 (9.5) | 280 (6.9) | 30 (16.5) | 35 (12.8) | 525 (5.3) | 300 (11.8) | 180 (18.0) | 85 (22.1) |
| South East | 30 (3.5) | 75 (3.3) | SN | SN | 45 (10.3) | 80 (7.0) | 10 (19.0) | 10 (13.6) | 190 (4.9) | 95 (9.6) | 60 (14.7) | 30 (18.9) |
| East Midlands | 95 (4.5) | 245 (3.6) | 10 (3.0) | 25 (4.9) | 165 (12.3) | 355 (8.2) | 30 (17.0) | 45 (14.0) | 655 (6.0) | 385 (12.6) | 250 (23.1) | 125 (25.7) |
| South West | 65 (4.5) | 120 (3.3) | SN | 10 (4.5) | 70 (10.0) | 150 (8.1) | 15 (22.2) | 15 (15.3) | 390 (5.7) | 195 (12.5) | 135 (21.2) | 65 (28.2) |
| **Flu vaccination, N (rate)** |  |  |  |  |  |  |  |  |  |  |  |  |
| Yes | 235 (6.3) | 555 (5.1) | 40 (9.2) | 65 (7.5) | 360 (14.8) | 630 (10.0) | 80 (25.9) | 85 (18.2) | 1,530 (7.8) | 765 (15.6) | 590 (27.5) | 275 (32.7) |
| No | 300 (3.2) | 655 (2.6) | 40 (4.0) | 60 (3.4) | 400 (7.1) | 920 (5.5) | 75 (11.0) | 85  (7.7) | 1,485 (3.8) | 895 (8.2) | 425 (11.7) | 185 (12.1) |
| **Period** |  |  |  |  |  |  |  |  |  |  |  |  |
| Pre-pandemic | 230 (4.5) | 545 (3.8) | 25 (5.5) | 60 (7.5) | 320 (10.8) | 665 (8.0) | 70 (18.2) | 80 (13.1) | 1,270 (5.7) | 690 (12.6) | 465 (20.8) | 200 (29.5) |
| During pandemic | 155 (4.2) | 285 (2.8) | 25 (9.1) | 25 (2.8) | 205 (8.5) | 415 (5.7) | 35 (11.0) | 50  (9.7) | 785 (4.4) | 435 (8.5) | 235 (13.3) | 140 (22.7) |
| After 2^nd^ lockdown | 150 (3.6) | 375 (3.1) | 30 (6.7) | 40 (4.8) | 230 (8.4) | 465 (6.3) | 45 (15.6) | 40  (9.0) | 955 (5.0) | 535 (10.3) | 310 (17.5) | 125 (16.3) |
| ^1^ ABs, antibiotics prescribed or not.  ^2^ Rate is the number of cases per 1000 patients with common infection, calculated by dividing the count of infection-related hospital admission cases (numerator) by the count of infection diagnosis (denominator) and then multiplied by 1000.  ^3^ SN, small numbers, equal or less than 5.  ^4^ BMI, Body Mass Index recorded in the last 5 years.  ^5^ CCI, Charlson Comorbidities Index, measured from 17 weighted conditions, including myocardial infarction, congestive heart failure, peripheral vascular disease, cerebrovascular disease, dementia, chronic pulmonary disease, Connective tissue disease, ulcer disease, mild liver disease, diabetes, hemiplegia, moderate or severe renal disease, diabetes with complications, any malignancy (including leukaemia and lymphoma), moderate or severe liver disease, metastatic solid tumour, and AIDS.  ^6^ IMD, Multiple Deprivation Index, quintile measured from patient-level address. | | | | | | | | | | | | |

Supplementary Table 8. Counts and rates of hospital admission of patients with URTI infections, including upper respiratory tract infection (URTI), cough, cold with cough, and sore throat. These infections include incident infections with no prescribed antibiotics, incident infections with prescribed antibiotics, prevalent infections with no prescribed antibiotics, and prevalent infections with prescribed antibiotics.

|  | **URTI** | | | | **Cough** | | | | **Cold with cough** | | | | **Sore throat** | | | |
| --- | --- | --- | --- | --- | --- | --- | --- | --- | --- | --- | --- | --- | --- | --- | --- | --- |
|  | **Incident** | | **Prevalent** | | **Incident** | | **Prevalent** | | **Incident** | | **Prevalent** | | **Incident** | | **Prevalent** | |
|  | **No** **ABs^1^** | **With ABs** | **No ABs** | **With ABs** | **No** **ABs** | **With ABs** | **No ABs** | **With ABs** | **No** **ABs** | **With ABs** | **No ABs** | **With ABs** | **No** **ABs** | **With ABs** | **No ABs** | **With ABs** |
| **Total, N cases** | 2,600 | 5,880 | 205 | 345 | 10,145 | 10,605 | 1,230 | 1,165 | 24,055 | 36,010 | 5,305 | 6,725 | 3,240 | 5,840 | 885 | 650 |
| **Age, N (rate^2^)** |  |  |  |  |  |  |  |  |  |  |  |  |  |  |  |  |
| 18-24 | 140 (6.1) | 180 (7.4) | 10 (15.6) | SN^3^ | 195 (5.5) | 175 (6.6) | 15  (5.9) | 10  (7.8) | 385 (8.0) | 525 (7.9) | 65 (14.0) | 55 (12.8) | 845 (16.4) | 1,760 (13.4) | 345 (45.6) | 290 (31.7) |
| 25-34 | 230 (5.9) | 325 (6.3) | 10  (7.6) | 15 (9.1) | 380 (5.6) | 480 (8.0) | 50  (8.5) | 50 (14.5) | 795 (8.7) | 1,380 (9.0) | 175 (15.5) | 210 (17.0) | 750 (11.5) | 1,615 (10.1) | 265 (32.8) | 175 (18.6) |
| 35-44 | 245 (6.9) | 370 (6.4) | SN | 25 (11.8) | 470 (5.8) | 555 (7.7) | 55  (6.9) | 50 (10.5) | 985 (9.5) | 1,640 (8.8) | 190 (12.6) | 250 (14.0) | 455 (9.4) | 820 (8.2) | 110 (22.6) | 65 (12.3) |
| 45-54 | 255 (7.1) | 560 (8.0) | 20 (11.4) | 30 (10.9) | 850 (6.3) | 995 (9.7) | 140 (10.4) | 125 (16.3) | 1,740 (10.8) | 2,815 (10.6) | 420 (17.3) | 520 (18.0) | 335 (8.5) | 555 (8.7) | 70 (18.7) | 40 (13.3) |
| 55-64 | 305 (9.5) | 795 (10.7) | 20 (11.7) | 40 (12.4) | 1,440 (7.4) | 1,470 (11.6) | 185 (10.2) | 160 (15.7) | 2,930 (13.2) | 4,480 (14.0) | 640 (20.4) | 875 (22.8) | 290 (9.5) | 385 (9.5) | 45 (17.2) | 25 (13.3) |
| 65-74 | 470 (18.9) | 1,095 (16.5) | 45 (33.7) | 75 (23.7) | 2,455 (10.4) | 2,355 (18.0) | 295 (15.3) | 295 (26.0) | 5,320 (20.1) | 7,385 (22.2) | 1,180 (34.4) | 1,510 (33.9) | 245 (11.4) | 310 (13.5) | 25 (12.9) | 30 (29.1) |
| 75+ | 955 (42.2) | 2,555 (41.3) | 95 (64.0) | 160 (49.0) | 4,360 (22.6) | 4,575 (38.7) | 495 (31.0) | 475 (44.5) | 11,905 (48.1) | 17,780 (49.7) | 2,630 (68.8) | 3,305 (62.2) | 325 (24.4) | 390 (30.9) | 30 (26.7) | 20 (37.4) |
| **Sex, N (rate)** |  |  |  |  |  |  |  |  |  |  |  |  |  |  |  |  |
| Male | 1,070 (14.4) | 2,485 (16.6) | 70 (22.4) | 150 (25.6) | 5,005 (11.6) | 4,720 (18.6) | 560 (15.9) | 525 (27.4) | 11,740 (22.7) | 15,935 (23.8) | 2,460 (37.1) | 3,010 (38.8) | 1,425 (15.4) | 2,185 (13.6) | 350 (34.0) | 215 (25.3) |
| Female | 1,530 (11.0) | 3,395 (13.2) | 135 (20.4) | 195 (17.8) | 5,140 (10.0) | 5,885 (15.4) | 670 (13.9) | 640 (21.2) | 12,315 (19.8) | 20,075 (19.8) | 2,845 (30.6) | 3,715 (30.5) | 1,815 (10.2) | 3,655 (9.9) | 535 (27.2) | 435 (20.0) |
| **BMI^4^, N (rate)** |  |  |  |  |  |  |  |  |  |  |  |  |  |  |  |  |
| Underweight | 75 (21.1) | 195 (28.8) | 10 (3.9) | 10 (37.0) | 375 (17.1) | 325 (26.9) | 30 (20.8) | 35 (36.1) | 825 (30.8) | 1,165 (36.6) | 175 (54.0) | 225 (53.9) | 65 (13.9) | 110 (13.2) | 25 (47.2) | 10 (18.3) |
| Healthy weight | 625 (12.8) | 1,355 (16.0) | 45 (11.6) | 90 (25.3) | 2,570 (11.1) | 2,515 (18.6) | 275 (14.7) | 270 (25.7) | 6,060 (21.8) | 8,715 (24.5) | 1,320 (36.2) | 1,690 (39.3) | 675 (10.4) | 1,270 (10.8) | 190 (26.4) | 125 (18.4) |
| Overweight | 675 (12.9) | 1,575 (14.8) | 55 (18.7) | 90 (20.1) | 2,700 (9.8) | 2,760 (16.1) | 320 (13.3) | 300 (21.8) | 6,345 (19.6) | 9,570 (21.2) | 1,445 (32.2) | 1,800 (32.7) | 640 (10.4) | 1,145 (10.3) | 155 (23.6) | 115 (18.6) |
| Obese | 720 (12.9) | 1,790 (14.4) | 45 (21.8) | 100 (17.5) | 3,155 (11.3) | 3,370 (16.6) | 420 (16.2) | 410 (24.4) | 7,070 (21.4) | 10,710 (20.1) | 1,570 (32.1) | 2,075 (31.1) | 780 (12.5) | 1,415 (10.8) | 185 (27.4) | 160 (21.0) |
| Unknown | 505 (9.6) | 965 (11.6) | 45 (236.8) | 55 (19.9) | 1,345 (9.8) | 1,630 (14.2) | 180 (13.7) | 150 (20.5) | 3,750 (20.8) | 5,845 (18.6) | 795 (30.8) | 940 (30.7) | 1,080 (14.1) | 1,900 (11.6) | 330 (37.0) | 235 (25.8) |
| **Ethnicity, N (rate)** |  |  |  |  |  |  |  |  |  |  |  |  |  |  |  |  |
| White | 1,380 (11.7) | 3,085 (14.5) | 105 (36.3) | 175 (19.1) | 5,890 (10.2) | 5,990 (16.4) | 705 (13.4) | 690 (22.9) | 13,805 (20.2) | 20,390 (21.0) | 3,090 (31.4) | 3,950 (32.7) | 1,730 (12.1) | 2,980 (10.9) | 465 (28.1) | 320 (19.8) |
| Non-White | 185 (8.1) | 495 (9.6) | 15 (8.5) | 20 (10.4) | 505 (8.8) | 585 (11.3) | 65 (11.0) | 50 (15.0) | 1,110 (14.6) | 1,760 (14.7) | 245 (21.4) | 245 (21.3) | 325 (9.7) | 490 (9.1) | 80 (27.4) | 50 (18.5) |
| Unknown | 1,030 (14.1) | 2,295 (16.1) | 85 (84.2) | 150 (26.3) | 3,750 (12.0) | 4,030 (18.4) | 460 (18.7) | 425 (26.7) | 9,140 (24.1) | 13,855 (23.5) | 1,965 (39.6) | 2,525 (37.6) | 1,180 (12.5) | 2,370 (11.6) | 345 (32.9) | 275 (24.1) |
| **CCI^5^, N (rate)** |  |  |  |  |  |  |  |  |  |  |  |  |  |  |  |  |
| Very low | 1,160 (7.8) | 2,385 (9.8) | 85 (13.98) | 135 (15.0) | 3,915 (7.6) | 4,050 (11.8) | 435 (9.4) | 365 (15.4) | 9,140 (14.6) | 13,290 (14.8) | 1,740 (20.9) | 2,170 (23.5) | 2,280 (11.0) | 4,240 (10.1) | 665 (28.8) | 470 (20.0) |
| Low | 970 (18.3) | 2,395 (18.3) | 90 (31.1) | 140 (23.2) | 4,140 (12.2) | 4,375 (18.8) | 535 (18.6) | 515 (26.3) | 9,730 (24.3) | 15,005 (24.3) | 2,290 (39.9) | 2,895 (35.7) | 740 (13.6) | 1,295 (12.9) | 200 (33.5) | 145 (24.0) |
| Medium | 320 (35.5) | 800 (31.6) | 20 (33.9) | 55 (39.1) | 1,550 (21.3) | 1,635 (33.1) | 200 (30.1) | 205 (41.8) | 3,810 (42.2) | 5,750 (42.8) | 955 (65.1) | 1,210 (58.3) | 160 (21.7) | 230 (21.8) | 10 (14.4) | 20 (35.4) |
| High | 115 (67.8) | 220 (48.0) | 10 (87.0) | 10 (40.8) | 415 (32.1) | 395 (44.1) | 45 (36.0) | 60 (62.2) | 1,010 (60.1) | 1,470 (59.0) | 230 (76.0) | 330 (78.1) | 40 (29.3) | 50 (26.0) | 10 (62.5) | SN |
| Very high | 30 (60.6) | 75 (55.1) | - | SN | 125 (37.2) | 145 (56.8) | 15 (40.5) | 20 (76.9) | 360 (74.3) | 495 (66.8) | 85 (87.6) | 115 (93.9) | 20 (44.0) | 25 (42.7) | SN | SN |
| **Smoking status, N (rate)** |  |  |  |  |  |  |  |  |  |  |  |  |  |  |  |  |
| Smoker | 390 (11.6) | 750 (10.2) | 20  (4.9) | 45 (16.2) | 1,820 (8.2) | 1,650 (12.2) | 150 (10.7) | 145 (15.0) | 3,725 (14.6) | 4,840 (14.2) | 660 (24.8) | 845 (22.7) | 735 (16.5) | 1,295 (12.8) | 200 (37.5) | 130 (23.4) |
| Ex-smoker | 1,290 (15.4) | 3,180 (18.5) | 120 (20.6) | 200 (26.0) | 5,875 (12.4) | 5,975 (20.1) | 740 (18.1) | 720 (29.4) | 13,840 (24.6) | 20,505 (25.9) | 3,195 (40.1) | 4,050 (39.8) | 1,135 (11.9) | 2,060 (11.5) | 305 (29.3) | 210 (20.8) |
| Never smoked | 900 (9.6) | 1,925 (12.1) | 65 (15.2) | 100 (15.9) | 2,440 (9.9) | 2,955 (14.6) | 335 (11.9) | 300 (19.8) | 6,450 (20.3) | 10,585 (19.5) | 1,435 (27.3) | 1,820 (30.3) | 1,280 (10.2) | 2,325 (9.7) | 355 (26.0) | 290 (20.7) |
| Unknown | 10 (5.2) | 20 (10.2) | - | - | 10  (3.9) | 20 (10.9) | SN | SN | 40 (10.2) | 75 (14.5) | 10 (31.7) | 10 (35.1) | 90 (19.1) | 155 (12.8) | 25 (45.5) | 20 (31.2) |
| **IMD^6^, N (rate)** |  |  |  |  |  |  |  |  |  |  |  |  |  |  |  |  |
| 1 (most deprived) | 595 (12.9) | 1,510 (14.7) | 40 (19.4) | 80 (18.0) | 2,655 (11.2) | 2,660 (16.5) | 290 (15.7) | 275 (22.7) | 5,915 (20.8) | 8,320 (20.1) | 1,265 (35.0) | 1,570 (32.4) | 770 (11.9) | 1,345 (10.7) | 215 (33.3) | 135 (20.1) |
| 2 | 515 (12.1) | 1,220 (13.8) | 45 (23.0) | 85 (23.1) | 2,115 (11.1) | 2,230 (16.3) | 255 (16.1) | 245 (24.0) | 4,865 (21.1) | 7,255 (20.9) | 1,050 (33.8) | 1,330 (32.9) | 710 (12.9) | 1,165 (10.5) | 165 (27.5) | 130 (20.8) |
| 3 | 500 (11.3) | 1,155 (14.4) | 40 (20.4) | 65 (19.4) | 2,090 (10.9) | 2,105 (16.5) | 245 (13.9) | 255 (24.4) | 4,890 (21.1) | 7,370 (21.9) | 1,070 (32.1) | 1,420 (35.1) | 655 (12.2) | 1,210 (11.4) | 180 (29.6) | 135 (21.7) |
| 4 | 525 (12.9) | 1,035 (15.0) | 40 (21.9) | 70 (25.8) | 1,780 (10.7) | 1,940 (17.8) | 255 (16.4) | 215 (25.6) | 4,415 (21.9) | 6,930 (23.0) | 1,035 (34.9) | 1,255 (34.7) | 555 (11.5) | 1,090 (11.4) | 145 (25.9) | 110 (19.6) |
| 5 (most affluent) | 425 (11.6) | 860 (14.5) | 40 (22.8) | 40 (17.4) | 1,370 (9.6) | 1,505 (16.3) | 165 (11.5) | 160 (21.7) | 3,575 (20.7) | 5,545 (21.6) | 780 (29.5) | 1,035 (34.0) | 490 (11.1) | 930 (11.1) | 155 (29.2) | 125 (25.1) |
| Unknown | 35 (11.2) | 95 (14.0) | SN | - | 135 (8.8) | 165 (16.4) | 15 (11.3) | 20 (24.1) | 395 (21.3) | 585 (21.1) | 100 (38.9) | 120 (34.7) | 60 (13.3) | 100 (11.9) | 20 (36.4) | 15 (30.6) |
| **Season, N (rate)** |  |  |  |  |  |  |  |  |  |  |  |  |  |  |  |  |
| Spring | 615 (12.2) | 1,375 (13.9) | 45 (18.1) | 70 (16.5) | 2,610 (10.3) | 2,665 (16.0) | 325 (13.5) | 315 (22.9) | 5,945 (19.6) | 8,620 (20.8) | 1,320 (29.2) | 1,600 (30.8) | 890 (12.0) | 1,525 (10.4) | 250 (28.5) | 155 (17.3) |
| Summer | 430 (14.1) | 1,050 (15.1) | 45 (34.6) | 60 (23.8) | 2,355 (10.6) | 2,020 (17.0) | 260 (14.0) | 210 (24.0) | 5,545 (21.5) | 6,900 (22.7) | 1,065 (33.8) | 1,200 (35.4) | 850 (13.9) | 1,420 (11.3) | 235 (32.5) | 160 (22.8) |
| Autumn | 560 (12.3) | 1,265 (14.4) | 45 (23.7) | 75 (23.5) | 2,160 (11.1) | 2,285 (16.5) | 250 (15.5) | 225 (22.9) | 5,035 (21.6) | 7,865 (21.4) | 1,040 (34.2) | 1,395 (35.1) | 580 (10.9) | 1,190 (11.7) | 165 (28.7) | 120 (22.5) |
| Winter | 990 (11.4) | 2,185 (14.6) | 70 (17.3) | 140 (20.5) | 3,025 (11.0) | 3,635 (17.1) | 395 (16.1) | 415 (24.3) | 7,525 (21.9) | 12,620 (21.2) | 1,880 (36.1) | 2,535 (34.3) | 915 (11.2) | 1,705 (10.9) | 240 (29.2) | 215 (23.9) |
| **Region, N (rate)** |  |  |  |  |  |  |  |  |  |  |  |  |  |  |  |  |
| London | 105 (8.0) | 270 (9.1) | 10 (17.2) | 15 (16.9) | 280 (9.3) | 380 (12.6) | 35 (10.8) | 20 (12.1) | 655 (14.7) | 965 (16.2) | 160 (19.8) | 130 (20.0) | 160 (10.5) | 260 (9.7) | 50 (33.4) | 105 (8.0) |
| North East | 130 (13.2) | 245 (14.9) | SN | 10 (15.5) | 770 (11.6) | 590 (16.4) | 85 (16.3) | 70 (24.1) | 1,695 (21.9) | 1,950 (21.1) | 365 (35.8) | 360 (31.6) | 185 (12.5) | 275 (10.4) | 45 (29.5) | 130 (13.2) |
| North West | 280 (11.5) | 575 (15.5) | 15 (15.0) | 40 (24.8) | 1,095 (9.2) | 1,215 (17.2) | 145 (13.3) | 135 (21.3) | 2,315 (16.8) | 4,100 (21.5) | 570 (28.0) | 810 (31.5) | 345 (11.8) | 565 (10.7) | 80 (24.9) | 280 (11.5) |
| East | 560 (12.0) | 1,455 (13.6) | 40 (20.5) | 80 (20.3) | 2,355 (11.6) | 2,480 (15.8) | 285 (16.3) | 255 (21.9) | 5,460 (22.2) | 8,035 (20.5) | 1,160 (35.0) | 1,455 (32.9) | 670 (10.6) | 1,370 (10.1) | 200 (29.3) | 560 (12.0) |
| West Midlands | 140 (14.3) | 350 (14.4) | 25 (47.6) | 20 (18.7) | 440 (13.1) | 535 (16.8) | 50 (14.9) | 60 (28.2) | 1,075 (25.5) | 1,735 (21.5) | 235 (37.5) | 285 (34.2) | 185 (13.6) | 310 (11.6) | 45 (34.4) | 140 (14.3) |
| Yorkshire and The Humber | 410 (12.6) | 1,145 (14.6) | 55 (24.1) | 60 (17.2) | 1,610 (10.4) | 1,685 (16.9) | 195 (13.7) | 180 (22.7) | 3,900 (20.9) | 6,230 (20.9) | 955 (34.6) | 1,215 (33.6) | 520 (12.3) | 900 (10.3) | 140 (27.5) | 410 (12.6) |
| South East | 195 (13.7) | 320 (16.5) | 10 (17.4) | 20 (21.2) | 565 (9.7) | 665 (17.0) | 90 (17.1) | 85 (26.0) | 1,500 (21.3) | 2,100 (22.0) | 320 (33.9) | 405 (36.6) | 210 (13.0) | 375 (13.2) | 60 (30.3) | 195 (13.7) |
| East Midlands | 500 (12.5) | 1,075 (16.1) | 35 (20.6) | 70 (20.8) | 2,020 (12.2) | 2,015 (18.0) | 230 (15.3) | 240 (27.2) | 4,810 (24.4) | 7,090 (22.6) | 1,065 (37.0) | 1,430 (36.6) | 585 (12.1) | 1,090 (11.2) | 155 (28.7) | 500 (12.5) |
| South West | 275 (12.0) | 440 (16.7) | 10 (13.7) | 20 (24.0) | 1,015 (8.9) | 1,045 (17.1) | 110 (12.9) | 120 (25.5) | 2,640 (19.3) | 3,805 (23.6) | 470 (30.6) | 635 (37.4) | 380 (14.0) | 695 (14.1) | 105 (33.5) | 275 (12.0) |
| **Flu vaccination, N (rate)** |  |  |  |  |  |  |  |  |  |  |  |  |  |  |  |  |
| Yes | 1,415 (21.2) | 3,610 (21.7) | 120 (31.0) | 215 (26.5) | 6,625 (12.9) | 6,955 (22.5) | 820 (19.4) | 800 (29.6) | 15,690 (26.3) | 23,860 (28.9) | 3,575 (42.9) | 4,675 (41.4) | 805 (13.2) | 1,290 (13.7) | 130 (21.3) | 125 (23.2) |
| No | 1,180 (8.1) | 2,270 (9.5) | 85 (14.5) | 130 (15.0) | 3,520 (8.1) | 3,650 (11.1) | 405 (9.9) | 365 (16.3) | 8,365 (15.4) | 12,150 (14.1) | 1,725 (22.7) | 2,050 (23.7) | 2,430 (11.6) | 4,545 (10.4) | 755 (31.6) | 520 (20.9) |
| **Period** |  |  |  |  |  |  |  |  |  |  |  |  |  |  |  |  |
| Pre-pandemic | 1,455 (11.9) | 3,105 (16.2) | 110 (21.0) | 205 (24.3) | 3,770 (11.3) | 4,755 (19.1) | 555 (17.8) | 520 (25.9) | 9,515 (22.9) | 18,085 (24.0) | 2,545 (38.2) | 3,600 (37.9) | 1,420 (11.9) | 2,795 (12.6) | 385 (31.4) | 335 (26.1) |
| During pandemic | 540 (11.8) | 1,260 (14.8) | 50 (21.3) | 75 (18.9) | 2,770 (10.0) | 2,715 (17.3) | 325 (12.8) | 330 (23.8) | 6,345 (19.3) | 8,295 (21.8) | 1,375 (29.4) | 1,635 (31.8) | 785 (10.8) | 1,440 (10.5) | 215 (23.6) | 165 (19.0) |
| After 2^nd^ lockdown | 605 (13.3) | 1,510 (11.7) | 45 (21.1) | 65 (14.9) | 3,605 (10.7) | 3,135 (13.5) | 350 (13.1) | 315 (20.4) | 8,190 (20.8) | 9,625 (17.5) | 1,380 (30.1) | 1,490 (28.1) | 1,035 (13.3) | 1,600 (9.3) | 285 (33.1) | 150 (17.2) |
| ^1^ ABs, antibiotics prescribed or not.  ^2^ Rate is the number of cases per 1000 patients with common infection, calculated by dividing the count of infection-related hospital admission cases (numerator) by the count of infection diagnosis (denominator) and then multiplied by 1000.  ^3^ SN, small numbers, equal or less than 5.  ^4^ BMI, Body Mass Index recorded in the last 5 years.  ^5^ CCI, Charlson Comorbidities Index, measured from 17 weighted conditions, including myocardial infarction, congestive heart failure, peripheral vascular disease, cerebrovascular disease, dementia, chronic pulmonary disease, Connective tissue disease, ulcer disease, mild liver disease, diabetes, hemiplegia, moderate or severe renal disease, diabetes with complications, any malignancy (including leukaemia and lymphoma), moderate or severe liver disease, metastatic solid tumour, and AIDS.  ^6^ IMD, Multiple Deprivation Index, quintile measured from patient-level address. | | | | | | | | | | | | | | | | |

### Cox models with overall data

#### Performance

All Cox models converged, except the models for prevalent sinusitis with no antibiotics, prevalent sinusitis with antibiotics, and prevalent otitis externa with antibiotics. Extreme values were calculated in hazard ratios (HRs) of some models, i.e., models for incident and prevalent otitis media with antibiotics and models for prevalent URTI with and without antibiotics. C-statistics of the converged models without extreme HRs using development and validation datasets were close; therefore, they were validated.

Supplementary Table 9. C-statistics of Cox models for hospital admissions related to common infections, including lower respiratory tract infection, upper respiratory tract infection, urinary tract infection (UTI), sinusitis, otitis media, and otitis externa, using overall data (from January 2019 to August 2022).

|  | | | **C-statistics** | |
| --- | --- | --- | --- | --- |
|  |  |  | **Development dataset** | **Validation dataset** |
| LRTI | Incident | No ABs^1^ | 0.67 | 0.67 |
|  |  | With ABs | 0.72 | 0.72 |
|  | Prevalent | No ABs | 0.67 | 0.64 |
|  |  | With ABs | 0.68 | 0.69 |
| URTI | Incident | No ABs | 0.71 | 0.71 |
|  |  | With ABs | 0.71 | 0.71 |
|  | Prevalent | No ABs | 0.71 | 0.70 |
|  |  | With ABs | 0.69 | 0.69 |
| UTI | Incident | No ABs | 0.73 | 0.73 |
|  |  | With ABs | 0.75 | 0.75 |
|  | Prevalent | No ABs | 0.69 | 0.70 |
|  |  | With ABs | 0.69 | 0.69 |
| Sinusitis | Incident | No ABs | 0.71 | 0.70 |
|  |  | With ABs | 0.69 | 0.68 |
|  | Prevalent | No ABs | - | - |
|  |  | With ABs | - | - |
| Otitis media | Incident | No ABs | 0.69 | 0.69 |
|  |  | With ABs | - | - |
|  | Prevalent | No ABs | 0.73 | 0.66 |
|  |  | With ABs | - | - |
| Otitis externa | Incident | No ABs | 0.71 | 0.70 |
|  |  | With ABs | 0.67 | 0.63 |
|  | Prevalent | No ABs | 0.75 | 0.75 |
|  |  | With ABs | - | - |
| URTI components | |  |  |  |
| Specific URTI | Incident | No ABs | 0.76 | 0.74 |
|  |  | With ABs | 0.73 | 0.73 |
|  | Prevalent | No ABs | - | - |
|  |  | With ABs | - | - |
| Cough | Incident | No ABs | 0.71 | 0.71 |
|  |  | With ABs | 0.70 | 0.71 |
|  | Prevalent | No ABs | 0.73 | 0.70 |
|  |  | With ABs | 0.70 | 0.67 |
| Cold with cough | Incident | No ABs | 0.72 | 0.72 |
|  |  | With ABs | 0.72 | 0.72 |
|  | Prevalent | No ABs | 0.72 | 0.72 |
|  |  | With ABs | 0.69 | 0.69 |
| Sore throat | Incident | No ABs | 0.65 | 0.63 |
|  |  | With ABs | 0.62 | 0.61 |
|  | Prevalent | No ABs | 0.65 | 0.60 |
|  |  | With ABs | 0.65 | 0.62 |
| ^1^ABs, antibiotics prescribed or not. | | | | |

#### Hazard ratios

Supplementary Table 10. Adjusted hazard ratios of Cox models for hospital admissions related to common infections, including lower respiratory tract infection (LRTI), upper respiratory tract infections (URTI), and urinary tract infection (UTI), using overall data (from January 2019 to August 2022).

|  | **LRTI, adjusted HR^1^ (95% CI^2^)** | | | **URTI, adjusted HR^1^ (95% CI)** | | | **UTI, adjusted HR^1^ (95% CI)** | | |
| --- | --- | --- | --- | --- | --- | --- | --- | --- | --- |
|  | **Incident** | **Prevalent** | | **Incident** | **Prevalent** | | **Incident** | **Prevalent** | |
|  | **With ABs^1^** | **No ABs** | **With ABs** | **No ABs** | **With ABs** | **No ABs** | **With ABs** | **No ABs** | **With ABs** |
| **Sex** |  |  |  |  |  |  |  |  |  |
| Male | 1.20  (1.17-1.23) | 1.23  (1.15-1.33) | 1.18  (1.11-1.26) | 1.20  (1.18-1.23) | 1.21  (1.15-1.28) | 1.25  (1.19-1.31) | 2.20  (2.15-2.25) | 1.59  (1.49-1.7) | 1.85  (1.76-1.95) |
| **Age** |  |  |  |  |  |  |  |  |  |
| 25-34 | 1.12  (0.99-1.26) | 0.74  (0.51-1.07) | 1.13  (0.79-1.61) | 0.86  (0.81-0.91) | 0.68  (0.59-0.79) | 0.76  (0.64-0.89) | 1.04  (0.95-1.14) | 1.18  (0.9-1.56) | 0.96  (0.78-1.2) |
| 35-44 | 1.06  (0.94-1.19) | 0.73  (0.51-1.04) | 1.01  (0.72-1.42) | 0.76  (0.72-0.81) | 0.4  (0.34-0.48) | 0.58  (0.49-0.69) | 1.06  (0.97-1.16) | 1.09  (0.82-1.45) | 1.01  (0.81-1.24) |
| 45-54 | 1.32  (1.18-1.47) | 0.81  (0.58-1.14) | 1.03  (0.74-1.43) | 0.89  (0.84-0.94) | 0.51  (0.44-0.59) | 0.69  (0.6-0.81) | 1.30  (1.19-1.41) | 1.41  (1.09-1.83) | 1.05  (0.86-1.28) |
| 55-64 | 1.64  (1.47-1.82) | 1.00  (0.72-1.38) | 1.4  (1.01-1.94) | 1.06  (1.01-1.12) | 0.51  (0.44-0.58) | 0.77  (0.67-0.89) | 1.73  (1.6-1.88) | 1.75  (1.37-2.24) | 1.23  (1.02-1.48) |
| 65-74 | 2.49  (2.24-2.76) | 1.60  (1.16-2.20) | 1.71  (1.24-2.37) | 1.56  (1.48-1.64) | 0.76  (0.67-0.87) | 1.08  (0.93-1.24) | 2.57  (2.37-2.77) | 2.45  (1.93-3.11) | 1.67  (1.39-2.0) |
| 75+ | 5.09  (4.59-5.65) | 2.20  (1.61-3.02) | 2.91  (2.12-4.01) | 3.22  (3.06-3.39) | 1.39  (1.22-1.58) | 1.78  (1.55-2.05) | 5.23  (4.84-5.65) | 3.99  (3.16-5.04) | 2.94  (2.45-3.51) |
| **BMI^2^** |  |  |  |  |  |  |  |  |  |
| Underweight | 1.45  (1.36-1.56) | 1.34  (1.11-1.61) | 1.37  (1.16-1.62) | 1.42  (1.34-1.5) | 1.37  (1.17-1.6) | 1.39  (1.21-1.6) | 1.43  (1.34-1.54) | 1.26  (1.02-1.56) | 1.16  (0.98-1.38) |
| Overweight | 0.83  (0.8-0.86) | 0.94  (0.85-1.03) | 0.82  (0.75-0.89) | 0.86  (0.83-0.88) | 0.88  (0.82-0.95) | 0.81  (0.76-0.87) | 0.92  (0.9-0.95) | 0.99  (0.91-1.08) | 0.98  (0.92-1.04) |
| Obese | 0.88  (0.85-0.91) | 0.89  (0.81-0.98) | 0.88  (0.81-0.95) | 0.92  (0.89-0.94) | 0.94  (0.87-1.01) | 0.85  (0.80-0.91) | 1.08  (1.05-1.11) | 1.25  (1.15-1.37) | 1.07  (1.00-1.14) |
| Unknown | 1.03  (0.99-1.07) | 1.05  (0.94-1.18) | 1.01  (0.92-1.12) | 1.09  (1.06-1.12) | 1.24  (1.14-1.35) | 1.02  (0.94-1.11) | 1.11  (1.07-1.15) | 1.23  (1.11-1.35) | 1.14  (1.06-1.23) |
| **Ethnicity** |  |  |  |  |  |  |  |  |  |
| White | 1.09  (1.03-1.16) | 1.00  (0.83-1.21) | 1.23  (1.05-1.45) | 1.15  (1.1-1.2) | 1.05  (0.92-1.19) | 1.30  (1.14-1.48) | 1.08  (1.02-1.15) | 0.95  (0.8-1.13) | 0.95  (0.83-1.09) |
| Unknown | 1.20  (1.13-1.27) | 1.14  (0.94-1.38) | 1.32  (1.12-1.56) | 1.24  (1.19-1.30) | 1.30  (1.14-1.48) | 1.45  (1.27-1.66) | 1.20  (1.13-1.27) | 1.12  (0.95-1.33) | 1.06  (0.93-1.22) |
| **CCI^3^** |  |  |  |  |  |  |  |  |  |
| Low | 1.22  (1.19-1.26) | 1.21  (1.11-1.32) | 1.18  (1.1-1.27) | 1.22  (1.19-1.25) | 1.43  (1.34-1.52) | 1.19  (1.12-1.26) | 1.37  (1.34-1.41) | 1.35  (1.26-1.46) | 1.38  (1.3-1.45) |
| Medium | 1.62  (1.56-1.68) | 1.50  (1.35-1.68) | 1.58  (1.44-1.73) | 1.66  (1.61-1.71) | 1.91  (1.76-2.08) | 1.63  (1.51-1.76) | 1.67  (1.62-1.73) | 1.68  (1.53-1.85) | 1.65  (1.54-1.78) |
| High | 2.05  (1.92-2.18) | 1.69  (1.42-2.02) | 1.84  (1.58-2.14) | 2.05  (1.94-2.16) | 2.0  (1.73-2.31) | 2.03  (1.80-2.30) | 2.00  (1.89-2.11) | 1.82  (1.57-2.11) | 2.07  (1.85-2.33) |
| Very high | 2.46  (2.21-2.73) | 2.08  (1.59-2.71) | 2.88  (2.33-3.56) | 2.58  (2.37-2.81) | 2.34  (1.85-2.95) | 2.80  (2.31-3.39) | 2.32  (2.11-2.56) | 1.93  (1.51-2.48) | 2.12  (1.74-2.59) |
| **Smoking** |  |  |  |  |  |  |  |  |  |
| Smoker | 0.88  (0.84-0.91) | 0.97  (0.86-1.10) | 0.86  (0.78-0.94) | 0.89  (0.87-0.92) | 0.88  (0.81-0.95) | 0.80  (0.74-0.86) | 1.21  (1.17-1.26) | 1.24  (1.11-1.39) | 1.19  (1.09-1.29) |
| Never smoked | 0.98  (0.96-1.01) | 0.87  (0.80-0.94) | 0.95  (0.89-1.02) | 0.96  (0.94-0.99) | 0.90  (0.85-0.96) | 0.96  (0.91-1.02) | 0.94  (0.92-0.96) | 0.95  (0.89-1.02) | 0.95  (0.90-1.00) |
| Unknown | 1.10  (0.85-1.43) | 0.95  (0.45-2.02) | 1.46  (0.76-2.83) | 0.99  (0.85-1.14) | 1.23  (0.86-1.76) | 1.27  (0.86-1.88) | 1.15  (0.92-1.43) | 0.57  (0.24-1.38) | 0.46  (0.21-1.03) |
| **IMD^4^** |  |  |  |  |  |  |  |  |  |
| 1 (most deprived) | 1.05  (1.02-1.10) | 1.05  (0.94-1.17) | 1.05  (0.96-1.15) | 1.01  (0.98-1.04) | 1.03  (0.95-1.12) | 1.01  (0.94-1.09) | 1.08  (1.05-1.12) | 1.06  (0.96-1.17) | 1.01  (0.94-1.09) |
| 3 | 0.98  (0.94-1.01) | 0.98  (0.88-1.10) | 0.99  (0.9-1.08) | 0.97  (0.94-1.00) | 0.89  (0.82-0.96) | 0.97  (0.90-1.04) | 0.94  (0.91-0.97) | 0.88  (0.8-0.96) | 0.87  (0.81-0.93) |
| 4 | 0.99  (0.95-1.03) | 0.95  (0.85-1.06) | 0.96  (0.88-1.06) | 1.00  (0.97-1.03) | 0.98  (0.90-1.06) | 0.95  (0.88-1.03) | 0.88  (0.85-0.91) | 0.79  (0.71-0.87) | 0.84  (0.78-0.9) |
| 5 (most affluent) | 0.93  (0.90-0.97) | 0.91  (0.81-1.03) | 0.92  (0.83-1.01) | 0.93  (0.9-0.96) | 0.83  (0.76-0.91) | 0.94  (0.87-1.02) | 0.82  (0.79-0.85) | 0.73  (0.66-0.80) | 0.75  (0.69-0.80) |
| Unknown | 0.98  (0.89-1.08) | 1.19  (0.91-1.57) | 1.07  (0.85-1.33) | 0.97  (0.9-1.05) | 1.14  (0.93-1.39) | 1.00  (0.83-1.21) | 0.98  (0.89-1.06) | 1.02  (0.79-1.31) | 0.92  (0.77-1.11) |
| **Season** |  |  |  |  |  |  |  |  |  |
| Spring | 0.94  (0.90-0.97) | 0.88  (0.79-0.98) | 0.88  (0.81-0.96) | 0.94  (0.91-0.96) | 0.94  (0.87-1.02) | 0.86  (0.80-0.93) | 0.99  (0.96-1.02) | 0.98  (0.9-1.07) | 0.92  (0.86-0.98) |
| Summer | 1.03  (0.99-1.07) | 0.99  (0.89-1.11) | 0.89  (0.81-0.98) | 1.0  (0.97-1.03) | 1.03  (0.95-1.11) | 0.95  (0.88-1.03) | 1.01  (0.98-1.04) | 1.02  (0.93-1.11) | 0.96  (0.9-1.03) |
| Winter | 0.96  (0.93-0.99) | 0.97  (0.88-1.07) | 0.91  (0.84-0.98) | 0.98  (0.95-1.0) | 1.09  (1.01-1.17) | 0.95  (0.89-1.02) | 0.98  (0.95-1.01) | 0.96  (0.88-1.05) | 0.91  (0.85-0.97) |
| **Region** |  |  |  |  |  |  |  |  |  |
| London | 0.95  (0.87-1.03) | 1.05  (0.82-1.33) | 0.99  (0.77-1.27) | 0.93  (0.88-0.98) | 0.86  (0.73-1.01) | 0.81  (0.67-0.97) | 1.04  (0.97-1.11) | 1.11  (0.93-1.33) | 0.96  (0.8-1.15) |
| North East | 0.96  (0.91-1.02) | 1.03  (0.88-1.21) | 0.86  (0.74-0.99) | 1.02  (0.98-1.07) | 1.00 (0.88-1.12) | 0.98  (0.87-1.11) | 1.08  (1.02-1.13) | 1.01  (0.87-1.17) | 1.05  (0.94-1.19) |
| North West | 0.96  (0.92-1.00) | 0.88  (0.77-1.00) | 0.93  (0.84-1.03) | 1.03  (0.99-1.06) | 0.79  (0.72-0.87) | 0.98  (0.90-1.07) | 0.97  (0.93-1.01) | 0.78  (0.70-0.88) | 0.97  (0.89-1.05) |
| West Midlands | 1.05  (0.99-1.12) | 1.05  (0.87-1.27) | 0.92  (0.79-1.08) | 1.08  (1.03-1.13) | 1.10  (0.96-1.26) | 1.10  (0.96-1.25) | 1.01  (0.95-1.07) | 1.21  (1.03-1.42) | 1.12  (0.99-1.26) |
| Yorkshire and The Humber | 0.96  (0.93-1.00) | 0.94  (0.84-1.05) | 0.96  (0.88-1.05) | 1.03  (1.00-1.06) | 0.96  (0.88-1.04) | 1.0  (0.93-1.09) | 1.05  (1.02-1.09) | 0.95  (0.86-1.05) | 1.08  (1.01-1.16) |
| South East | 0.98  (0.93-1.04) | 0.91  (0.77-1.08) | 0.93  (0.81-1.08) | 1.01  (0.96-1.05) | 0.96  (0.86-1.08) | 1.03  (0.92-1.15) | 0.93  (0.88-0.97) | 0.80  (0.69-0.93) | 0.95  (0.85-1.06) |
| East Midlands | 1.07  (1.03-1.11) | 1.06  (0.95-1.19) | 1.03  (0.94-1.12) | 1.10  (1.07-1.14) | 1.07  (0.99-1.16) | 1.12  (1.04-1.20) | 0.99  (0.96-1.02) | 1.03  (0.94-1.13) | 1.01  (0.95-1.09) |
| South West | 1.05  (1.01-1.1) | 1.01  (0.89-1.16) | 1.08  (0.96-1.21) | 1.07  (1.03-1.11) | 0.88  (0.80-0.98) | 1.12  (1.02-1.23) | 0.91  (0.87-0.94) | 0.88  (0.79-0.99) | 0.90  (0.82-0.98) |
| **Flu vaccination** |  |  |  |  |  |  |  |  |  |
| Yes | 0.97  (0.94-1.00) | 0.98  (0.90-1.07) | 0.95  (0.88-1.01) | 0.98  (0.96-1.00) | 0.91  (0.85-0.97) | 0.98  (0.92-1.04) | 0.96  (0.93-0.98) | 0.95  (0.88-1.02) | 1.00  (0.95-1.06) |
| **Count of antibiotic prescription in the one year before** | 1.10  (1.09-1.10) | 1.05  (1.04-1.06) | 1.09  (1.07-1.10) | 1.12  (1.12-1.13) | 1.14  (1.13-1.15) | 1.11  (1.10-1.12) | 1.05  (1.04-1.05) | 1.03  (1.02-1.04) | 1.04  (1.03-1.04) |
| ^1^ HR, hazard ratio.  ^2^ CI, confidence interval.  ^3^ ABs, antibiotics prescribed or not.  ^4^ BMI, Body Mass Index recorded in the last 5 years.  ^5^ CCI, Charlson Comorbidities Index, measured from 17 weighted conditions, including myocardial infarction, congestive heart failure, peripheral vascular disease, cerebrovascular disease, dementia, chronic pulmonary disease, Connective tissue disease, ulcer disease, mild liver disease, diabetes, hemiplegia, moderate or severe renal disease, diabetes with complications, any malignancy (including leukaemia and lymphoma), moderate or severe liver disease, metastatic solid tumour, and AIDS.  ^6^ IMD, Multiple Deprivation Index, quintile measured from patient-level address.  Reference group for variable sex is female, for age is 18-25, for BMI is healthy weight, for ethnicity is non-white, for CCI is very low, for smoking status is ex-smoker, for IMD is 2, for season is autumn, for region is east, for flu vaccination is no. | | | | | | | | | |

Supplementary Table 11. Adjusted hazard ratios of Cox models for hospital admissions related to other common infections, including sinusitis, otitis media, and otitis externa, using overall data (from January 2019 to August 2022).

|  | **Sinusitis, adjusted HR^1^**  **(95% CI^2^)** | | **Otitis media, adjusted HR (95% CI)** | | **Otitis externa, adjusted HR**  **(95% CI)** | | |
| --- | --- | --- | --- | --- | --- | --- | --- |
|  | **Incident** | | **Incident** | **Prevalent** | **Incident** | | **Prevalent** |
|  | **No ABs^3^** | **With ABs** | **No ABs** | **No ABs** | **No ABs** | **With ABs** | **No ABs** |
| **Sex** |  |  |  |  |  |  |  |
| Male | 1.40 (1.14-1.72) | 1.28  (1.10-1.48) | 1.29 (1.08-1.54) | 0.96 (0.64-1.43) | 1.18 (1.08-1.28) | 1.13 (1.0-1.27) | 1.40 (1.20-1.63) |
| **Age** |  |  |  |  |  |  |  |
| 25-34 | 0.94  (0.60-1.49) | 0.79  (0.57-1.08) | 1.24 (0.83-1.86) | 1.01 (0.40-2.51) | 0.92 (0.76-1.13) | 1.03 (0.79-1.34) | 0.87 (0.57-1.32) |
| 35-44 | 0.79  (0.50-1.27) | 0.59  (0.43-0.82) | 1.20 (0.79-1.82) | 1.48 (0.61-3.56) | 0.79 (0.64-0.97) | 0.82 (0.63-1.08) | 1.13 (0.75-1.7) |
| 45-54 | 0.80  (0.50-1.27) | 0.53  (0.38-0.73) | 1.14 (0.76-1.73) | 0.93 (0.36-2.41) | 0.79 (0.65-0.97) | 0.82 (0.63-1.07) | 0.81 (0.53-1.22) |
| 55-64 | 0.88  (0.55-1.40) | 0.68  (0.49-0.94) | 1.33 (0.89-2.01) | 0.54 (0.18-1.60) | 0.71 (0.58-0.88) | 0.76 (0.58-1.00) | 0.84 (0.55-1.27) |
| 65-74 | 0.89  (0.54-1.48) | 0.80 (0.57-1.12) | 1.52 (1-2.32) | 1.42 (0.53-3.82) | 1.00 (0.81-1.23) | 0.98 (0.74-1.31) | 0.97 (0.64-1.48) |
| 75+ | 2.55  (1.55-4.18) | 1.72  (1.22-2.43) | 2.71 (1.79-4.1) | 4.00 (1.57-10.18) | 2.13 (1.74-2.61) | 2.25 (1.71-2.96) | 3.07 (2.07-4.56) |
| **BMI**^4^ |  |  |  |  |  |  |  |
| Underweight | 1.47  (0.71-3.04) | 1.12  (0.65-1.93) | 1.66 (0.90-3.10) | 3.03 (0.88-10.38) | 1.30 (0.92-1.84) | 0.77 (0.43-1.38) | 0.64 (0.26-1.56) |
| Overweight | 0.80  (0.59-1.07) | 0.92  (0.76-1.11) | 1.18 (0.92-1.52) | 0.76 (0.41-1.39) | 1.01 (0.89-1.14) | 0.93 (0.79-1.10) | 1.21 (0.98-1.49) |
| Obese | 1.14  (0.87-1.51) | 1.14  (0.95-1.37) | 1.10 (0.85-1.4) | 1.26 (0.73-2.20) | 1.22 (1.08-1.38) | 1.07 (0.91-1.25) | 1.23 (0.99-1.52) |
| Unknown | 1.21  (0.89-1.62) | 1.06  (0.86-1.30) | 1.27 (0.97-1.65) | 1.43 (0.79-2.59) | 1.13 (0.98-1.29) | 0.81 (0.67-0.97) | 0.91 (0.69-1.18) |
| **Ethnicity** |  |  |  |  |  |  |  |
| White | 0.71  (0.49-1.03) | 0.84  (0.64-1.11) | 1.18 (0.83-1.68) | 0.40 (0.22-0.73) | 1.01 (0.85-1.21) | 1.02 (0.80-1.30) | 0.97 (0.70-1.34) |
| Unknown | 0.84  (0.57-1.24) | 0.87  (0.66-1.16) | 1.12 (0.78-1.62) | 0.47 (0.25-0.89) | 0.99 (0.82-1.19) | 1.10 (0.86-1.41) | 1.06 (0.76-1.48) |
| **CCI**^5^ |  |  |  |  |  |  |  |
| Low | 1.26  (1.00-1.60) | 1.38  (1.19-1.62) | 1.41 (1.15-1.72) | 1.26 (0.79-2.01) | 1.48 (1.34-1.64) | 1.27 (1.11-1.45) | 1.43 (1.19-1.71) |
| Medium | 1.84  (1.23-2.76) | 2.21  (1.71-2.85) | 1.98 (1.45-2.72) | 1.96 (0.94-4.10) | 2.25 (1.94-2.61) | 2.12 (1.74-2.59) | 2.68 (2.13-3.38) |
| High | 2.45  (1.19-5.05) | 2.67  (1.58-4.51) | 2.67 (1.54-4.66) | 1.78 (0.42-7.61) | 3.48 (2.73-4.42) | 2.49 (1.73-3.58) | 3.81 (2.7-5.38) |
| Very high | 1.47  (0.21-10.54) | 5.49  (2.82-10.68) | 0.67 (0.09-4.81) | 5.35 (1.23-23.10) | 6.13 (4.31-8.71) | 3.48 (1.91-6.36) | 3.83 (1.96-7.48) |
| **Smoking** |  |  |  |  |  |  |  |
| Smoker | 1.38  (1.04-1.83) | 1.18  (0.97-1.43) | 1.27 (0.99-1.62) | 1.50 (0.86-2.64) | 1.16 (1.02-1.31) | 0.99 (0.84-1.17) | 1.02 (0.80-1.29) |
| Never smoked | 0.93  (0.75-1.17) | 1.03  (0.89-1.19) | 1.02 (0.84-1.23) | 1.04 (0.66-1.63) | 0.89 (0.81-0.99) | 1.01 (0.89-1.15) | 0.95 (0.80-1.12) |
| Unknown | 1.47  (0.45-4.77) | 1.40  (0.56-3.47) | 1.11 (0.40-3.1) | 2.05 (0.26-16.12) | 0.45 (0.20-1.02) | 0.60 (0.22-1.64) | 0.79 (0.19-3.27) |
| **IMD**^6^ |  |  |  |  |  |  |  |
| 1 (most deprived) | 1.23  (0.90-1.68) | 1.03  (0.83-1.27) | 1.06 (0.82-1.38) | 0.64 (0.36-1.14) | 1.00 (0.88-1.14) | 0.95 (0.80-1.12) | 1.25 (0.99-1.58) |
| 3 | 1.05  (0.76-1.45) | 1.12  (0.91-1.37) | 0.95 (0.72-1.25) | 1.18 (0.69-2.01) | 1.02 (0.89-1.16) | 0.95 (0.80-1.14) | 0.96 (0.75-1.22) |
| 4 | 0.97  (0.70-1.36) | 0.91  (0.73-1.13) | 1.16 (0.89-1.51) | 0.44 (0.21-0.90) | 0.94 (0.82-1.08) | 0.89 (0.75-1.07) | 1.09 (0.86-1.38) |
| 5 (most affluent) | 1.06  (0.76-1.48) | 1.03  (0.83-1.28) | 1.16 (0.88-1.52) | 0.70 (0.37-1.32) | 0.91 (0.79-1.05) | 0.89 (0.73-1.07) | 0.89 (0.69-1.14) |
| Unknown | 1.43  (0.71-2.88) | 1.02  (0.61-1.71) | 1.06 (0.53-2.1) | 0.30 (0.04-2.29) | 0.94 (0.66-1.34) | 0.83 (0.52-1.33) | 1.01 (0.53-1.93) |
| **Season** |  |  |  |  |  |  |  |
| Spring | 1.26  (0.93-1.71) | 0.90 (0.74-1.09) | 0.88 (0.69-1.13) | 0.83 (0.46-1.52) | 0.91 (0.81-1.02) | 0.82 (0.70-0.97) | 0.72 (0.59-0.88) |
| Summer | 1.45  (1.07-1.98) | 0.94  (0.77-1.15) | 0.92 (0.73-1.16) | 1.19 (0.68-2.08) | 0.95 (0.85-1.07) | 0.91 (0.78-1.06) | 0.77 (0.64-0.93) |
| Winter | 1.22  (0.91-1.64) | 0.98  (0.82-1.17) | 0.90 (0.71-1.14) | 1.11 (0.65-1.90) | 0.83 (0.74-0.94) | 0.84 (0.72-0.98) | 0.65 (0.53-0.8) |
| **Region** |  |  |  |  |  |  |  |
| London | 1.11  (0.66-1.86) | 1.11  (0.74-1.66) | 1.20 (0.74-1.93) | 0.95 (0.34-2.69) | 1.45 (1.15-1.82) | 1.05 (0.74-1.5) | 1.45 (0.94-2.25) |
| North East | 1.35  (0.87-2.08) | 1.01  (0.70-1.45) | 1.40 (0.97-2.03) | 2.04 (0.89-4.71) | 1.27 (1.03-1.56) | 1.60 (1.23-2.07) | 1.53 (1.08-2.18) |
| North West | 0.94  (0.65-1.36) | 1.29  (1.02-1.64) | 1.21 (0.89-1.65) | 0.88 (0.39-2.03) | 1.29 (1.10-1.52) | 1.12 (0.89-1.41) | 1.21 (0.91-1.62) |
| West Midlands | 1.19  (0.70-2.00) | 1.55  (1.11-2.16) | 1.29 (0.83-2.03) | 1.30 (0.48-3.53) | 1.22 (0.96-1.57) | 1.38 (1.00-1.90) | 1.29 (0.79-2.09) |
| Yorkshire and The Humber | 1.06  (0.77-1.46) | 1.37  (1.11-1.68) | 1.22 (0.92-1.62) | 1.81 (0.99-3.29) | 1.33 (1.16-1.54) | 1.59 (1.32-1.91) | 1.48 (1.15-1.91) |
| South East | 0.81  (0.50-1.31) | 1.17  (0.87-1.57) | 1.27 (0.85-1.90) | 0.89 (0.30-2.64) | 1.22 (1.00-1.48) | 1.18 (0.90-1.55) | 1.30 (0.93-1.82) |
| East Midlands | 1.12  (0.81-1.53) | 1.25  (1.03-1.53) | 1.69 (1.32-2.18) | 1.59 (0.86-2.92) | 1.54 (1.34-1.76) | 1.61 (1.36-1.92) | 1.89 (1.50-2.40) |
| South West | 1.03  (0.71-1.49) | 1.15  (0.9-1.48) | 1.31 (0.94-1.82) | 1.98 (0.92-4.26) | 1.36 (1.17-1.59) | 1.72 (1.40-2.11) | 1.45 (1.10-1.92) |
| **Flu vaccination** |  |  |  |  |  |  |  |
| Yes | 1.26  (0.97-1.62) | 1.14  (0.97-1.34) | 1.14 (0.91-1.40) | 1.39 (0.83-2.32) | 1.05 (0.95-1.17) | 1.12 (0.97-1.29) | 1.03 (0.85-1.25) |
| **Count of antibiotic prescription in the one year before** | 1.26  (1.21-1.32) | 1.22  (1.19-1.26) | 1.23 (1.17-1.28) | 1.06 (0.96-1.19) | 1.29 (1.26-1.32) | 1.14 (1.10-1.18) | 1.21 (1.16-1.26) |
| ^1^ HR, hazard ratio.  ^2^ CI, confidence interval.  ^3^ ABs, antibiotics prescribed or not.  ^4^ BMI, Body Mass Index recorded in the last 5 years.  ^5^ CCI, Charlson Comorbidities Index, measured from 17 weighted conditions, including myocardial infarction, congestive heart failure, peripheral vascular disease, cerebrovascular disease, dementia, chronic pulmonary disease, Connective tissue disease, ulcer disease, mild liver disease, diabetes, hemiplegia, moderate or severe renal disease, diabetes with complications, any malignancy (including leukaemia and lymphoma), moderate or severe liver disease, metastatic solid tumour, and AIDS.  ^6^ IMD, Multiple Deprivation Index, quintile measured from patient-level address.  Reference group for variable sex is female, for age is 18-25, for BMI is healthy weight, for ethnicity is non-white, for CCI is very low, for smoking status is ex-smoker, for IMD is 2, for season is autumn, for region is east, for flu vaccination is no. | | | | | | | |

Supplementary Table 12. Adjusted hazard ratios of Cox models for hospital admissions related to the components of upper respiratory tract infections (URTI), including specific URTI, cough, cold with cough, and sore throat, using overall data (from January 2019 to August 2022).

|  | **URTI, adjusted HR^1^**  **(95% CI^2^)** | | **Cough, adjusted HR (95% CI)** | | | | **Cold with cough, adjusted HR (95% CI)** | | | | **Sore throat, adjusted HR (95% CI)** | | | |
| --- | --- | --- | --- | --- | --- | --- | --- | --- | --- | --- | --- | --- | --- | --- |
|  | **Incident** | | **Incident** | | **Prevalent** | | **Incident** | | **Prevalent** | | **Incident** | | **Prevalent** | |
|  | **With ABs^3^** | **No ABs** | **With ABs** | **No ABs** | **With ABs** | **No ABs** | **With ABs** | **No ABs** | **With ABs** | **No ABs** | **With ABs** | **No ABs** | **With ABs** | **No ABs** |
| **Sex** |  |  |  |  |  |  |  |  |  |  |  |  |  |  |
| Male | 1.25  (1.14-1.38) | 1.20  (1.13-1.28) | 1.18  (1.13-1.24) | 1.17  (1.12-1.23) | 1.22  (1.07-1.4) | 1.27  (1.11-1.45) | 1.15  (1.12-1.19) | 1.17  (1.14-1.19) | 1.22  (1.14-1.3) | 1.24  (1.17-1.31) | 1.56  (1.43-1.7) | 1.44  (1.34-1.53) | 1.40  (1.18-1.66) | 1.30  (1.05-1.59) |
| **Age** |  |  |  |  |  |  |  |  |  |  |  |  |  |  |
| 25-34 | 1.03  (0.8-1.32) | 0.86  (0.69-1.07) | 1.08  (0.88-1.32) | 1.22  (1.00-1.49) | 1.50  (0.76-2.96) | 1.21  (0.63-2.31) | 1.24  (1.08-1.44) | 1.23  (1.09-1.39) | 1.26  (0.89-1.78) | 1.43  (1.01-2.03) | 0.71  (0.63-0.8) | 0.73  (0.68-0.8) | 0.70  (0.58-0.85) | 0.59  (0.47-0.74) |
| 35-44 | 1.12  (0.87-1.43) | 0.89  (0.72-1.10) | 1.08  (0.88-1.31) | 1.12  (0.92-1.37) | 0.98  (0.5-1.95) | 0.88  (0.46-1.69) | 1.39  (1.21-1.6) | 1.18  (1.05-1.32) | 1.05  (0.75-1.48) | 1.17  (0.83-1.65) | 0.6  (0.52-0.69) | 0.64  (0.58-0.71) | 0.53  (0.41-0.67) | 0.42  (0.30-0.57) |
| 45-54 | 1.13  (0.88-1.45) | 1.08  (0.88-1.32) | 1.28  (1.06-1.54) | 1.38  (1.14-1.66) | 1.63  (0.87-3.07) | 1.20  (0.66-2.21) | 1.53  (1.34-1.75) | 1.39  (1.24-1.55) | 1.50  (1.09-2.07) | 1.45  (1.04-2.01) | 0.50  (0.43-0.59) | 0.62  (0.56-0.7) | 0.38  (0.28-0.52) | 0.39  (0.26-0.58) |
| 55-64 | 1.50  (1.18-1.91) | 1.35  (1.11-1.65) | 1.42  (1.19-1.7) | 1.54  (1.28-1.85) | 1.47  (0.79-2.75) | 0.94  (0.51-1.73) | 1.92  (1.69-2.18) | 1.70  (1.53-1.90) | 1.61  (1.18-2.21) | 1.68  (1.22-2.32) | 0.57  (0.48-0.67) | 0.66  (0.58-0.76) | 0.41  (0.28-0.59) | 0.44  (0.28-0.71) |
| 65-74 | 2.51  (1.98-3.19) | 1.83  (1.51-2.23) | 1.96  (1.64-2.34) | 2.09  (1.75-2.51) | 1.92  (1.03-3.59) | 1.65  (0.91-2.99) | 2.88  (2.54-3.27) | 2.52  (2.26-2.80) | 2.39  (1.74-3.27) | 2.18  (1.58-3.00) | 0.65  (0.54-0.78) | 0.82  (0.70-0.96) | 0.29  (0.17-0.49) | 0.95  (0.60-1.5) |
| 75+ | 4.77  (3.79-6.02) | 4.04  (3.34-4.90) | 3.81  (3.19-4.54) | 4.17  (3.49-5.00) | 3.47  (1.86-6.44) | 2.30  (1.27-4.16) | 6.17  (5.45-6.99) | 5.07  (4.56-5.63) | 4.22  (3.09-5.76) | 3.64  (2.65-5.01) | 1.26  (1.05-1.51) | 1.79  (1.54-2.07) | 0.65  (0.4-1.07) | 0.89  (0.50-1.58) |
| **BMI^4^** |  |  |  |  |  |  |  |  |  |  |  |  |  |  |
| Underweight | 1.30  (0.97-1.75) | 1.62  (1.36-1.93) | 1.53  (1.35-1.73) | 1.36  (1.19-1.56) | 1.45  (0.95-2.19) | 1.34  (0.89-2.02) | 1.40  (1.29-1.52) | 1.39  (1.29-1.49) | 1.44  (1.21-1.73) | 1.26  (1.07-1.49) | 1.31  (0.98-1.74) | 1.09  (0.87-1.37) | 1.44  (0.88-2.36) | 0.80  (0.39-1.65) |
| Overweight | 0.92  (0.81-1.05) | 0.86  (0.79-0.94) | 0.84  (0.79-0.89) | 0.83  (0.78-0.89) | 0.84  (0.70-1.01) | 0.77  (0.64-0.94) | 0.85  (0.82-0.89) | 0.85  (0.82-0.88) | 0.83  (0.76-0.91) | 0.80  (0.74-0.86) | 1.01  (0.89-1.14) | 0.91  (0.83-1.00) | 1.00  (0.78-1.28) | 0.88  (0.66-1.18) |
| Obese | 0.92  (0.81-1.05) | 0.91  (0.84-0.99) | 0.95  (0.90-1.01) | 0.91  (0.86-0.97) | 1.07  (0.90-1.28) | 0.92  (0.77-1.10) | 0.97  (0.93-1.01) | 0.89  (0.86-0.92) | 0.94  (0.86-1.02) | 0.84  (0.78-0.9) | 1.20  (1.06-1.35) | 0.97  (0.89-1.07) | 1.25  (0.99-1.58) | 1.06  (0.81-1.39) |
| Unknown | 1.08  (0.93-1.24) | 0.98  (0.89-1.09) | 1.28  (1.18-1.38) | 1.08  (1.01-1.17) | 1.24  (0.98-1.55) | 1.08  (0.85-1.36) | 1.40  (1.33-1.47) | 1.06  (1.02-1.11) | 1.22  (1.10-1.36) | 0.97  (0.88-1.06) | 1.25  (1.11-1.4) | 0.99  (0.91-1.08) | 1.16  (0.93-1.44) | 1.05  (0.81-1.36) |
| **Ethnicity** |  |  |  |  |  |  |  |  |  |  |  |  |  |  |
| White | 1.04  (0.86-1.25) | 1.17  (1.04-1.32) | 0.97  (0.86-1.08) | 1.14  (1.02-1.26) | 1.18  (0.84-1.65) | 1.14  (0.82-1.59) | 1.04  (0.97-1.12) | 1.13  (1.07-1.20) | 1.03  (0.88-1.20) | 1.23  (1.06-1.44) | 1.17  (1.01-1.36) | 1.11  (0.99-1.25) | 0.95  (0.71-1.26) | 0.92  (0.64-1.33) |
| Unknown | 1.23  (1.01-1.48) | 1.24  (1.10-1.40) | 1.12  (1.0-1.26) | 1.27  (1.14-1.42) | 1.56  (1.10-2.21) | 1.29  (0.92-1.82) | 1.19  (1.1-1.28) | 1.23  (1.16-1.31) | 1.22  (1.04-1.44) | 1.37  (1.17-1.6) | 1.12  (0.96-1.3) | 1.14  (1.01-1.28) | 1.03  (0.77-1.38) | 1.03  (0.71-1.49) |
| **CCI^5^** |  |  |  |  |  |  |  |  |  |  |  |  |  |  |
| Low | 1.57  (1.41-1.75) | 1.26  (1.17-1.35) | 1.32  (1.25-1.39) | 1.22  (1.15-1.28) | 1.47  (1.26-1.72) | 1.38  (1.17-1.62) | 1.33  (1.29-1.38) | 1.23  (1.20-1.27) | 1.4  (1.29-1.51) | 1.25  (1.17-1.34) | 1.22  (1.10-1.35) | 1.17  (1.09-1.27) | 1.43  (1.17-1.73) | 1.19  (0.95-1.50) |
| Medium | 2.09  (1.79-2.45) | 1.63  (1.47-1.8) | 1.84  (1.71-1.98) | 1.61  (1.5-1.73) | 1.80  (1.46-2.23) | 1.77  (1.44-2.19) | 1.80  (1.71-1.88) | 1.66  (1.60-1.73) | 1.90  (1.73-2.1) | 1.67  (1.53-1.82) | 1.65  (1.35-2.03) | 1.69  (1.44-2.00) | 0.83  (0.4-1.7) | 2.12  (1.27-3.55) |
| High | 3.43  (2.7-4.34) | 2.25  (1.91-2.64) | 2.55  (2.26-2.88) | 2.14  (1.90-2.42) | 1.99  (1.39-2.85) | 2.35  (1.69-3.26) | 2.37  (2.20-2.56) | 2.11  (1.98-2.25) | 1.99  (1.69-2.35) | 2.17  (1.89-2.49) | 2.22  (1.55-3.18) | 2.03  (1.48-2.78) | 1.66  (0.61-4.52) | 1.39  (0.44-4.45) |
| Very high | 4.23  (2.89-6.20) | 2.82  (2.15-3.70) | 3.05  (2.48-3.75) | 2.8  (2.31-3.39) | 2.21  (1.21-4.06) | 2.81  (1.63-4.84) | 2.98  (2.64-3.38) | 2.56  (2.31-2.84) | 2.31  (1.78-3.00) | 2.66  (2.14-3.31) | 3.58  (2.18-5.89) | 3.01  (1.89-4.81) | 3.08  (0.76-12.5) | 1.5  (0.21-10.83) |
| **Smoking** |  |  |  |  |  |  |  |  |  |  |  |  |  |  |
| Smoker | 1.24  (1.08-1.43) | 0.84  (0.76-0.93) | 0.88  (0.82-0.94) | 0.91  (0.85-0.97) | 0.75  (0.61-0.93) | 0.64  (0.52-0.80) | 0.85  (0.81-0.88) | 0.87  (0.83-0.90) | 0.92  (0.84-1.02) | 0.83  (0.76-0.91) | 1.37  (1.22-1.53) | 1.15  (1.06-1.25) | 1.13  (0.92-1.40) | 1.02  (0.79-1.32) |
| Never smoked | 0.89  (0.81-0.99) | 0.90  (0.84-0.96) | 1.16  (1.09-1.23) | 0.99  (0.94-1.04) | 0.93  (0.79-1.08) | 1.00  (0.85-1.17) | 1.19  (1.15-1.24) | 1.01  (0.98-1.04) | 0.94  (0.87-1.02) | 1.00  (0.94-1.07) | 0.92  (0.84-1.02) | 0.90  (0.84-0.96) | 0.82  (0.68-0.98) | 0.87  (0.7-1.07) |
| Unknown | 0.57  (0.25-1.28) | 1.07  (0.63-1.83) | 0.56  (0.27-1.19) | 1.40  (0.89-2.22) | 1.05  (0.14-7.57) | 1.30  (0.18-9.42) | 0.85  (0.59-1.23) | 1.08  (0.83-1.41) | 1.23  (0.58-2.60) | 1.60  (0.86-3.00) | 1.24  (0.96-1.60) | 0.84  (0.68-1.03) | 0.99  (0.62-1.59) | 0.90  (0.52-1.58) |
| **IMD^6^** |  |  |  |  |  |  |  |  |  |  |  |  |  |  |
| 1 (most deprived) | 1.08  (0.94-1.24) | 1.15  (1.05-1.25) | 1.07  (1.00-1.14) | 1.03  (0.97-1.10) | 1.11  (0.90-1.35) | 0.96  (0.78-1.17) | 1.09  (1.04-1.14) | 1.03  (0.99-1.06) | 1.09  (0.99-1.20) | 1.02  (0.93-1.11) | 0.90  (0.79-1.01) | 0.98  (0.89-1.07) | 1.17  (0.93-1.49) | 0.85  (0.64-1.13) |
| 3 | 0.88  (0.76-1.02) | 1.00  (0.91-1.10) | 0.93  (0.87-1.00) | 0.95  (0.88-1.01) | 0.9  (0.73-1.10) | 0.95  (0.77-1.16) | 0.97  (0.92-1.01) | 0.97  (0.93-1.01) | 0.89  (0.80-0.98) | 0.99  (0.91-1.08) | 1.00  (0.88-1.13) | 1.04  (0.95-1.15) | 1.08  (0.85-1.38) | 1.03  (0.78-1.35) |
| 4 | 1.02  (0.88-1.17) | 1.00  (0.91-1.11) | 0.90  (0.84-0.97) | 1.04  (0.97-1.11) | 1.13  (0.92-1.38) | 1.00  (0.81-1.23) | 0.97  (0.92-1.01) | 1.00  (0.96-1.04) | 0.98  (0.88-1.08) | 0.97  (0.88-1.06) | 0.92  (0.81-1.05) | 1.01  (0.92-1.12) | 0.95  (0.73-1.23) | 0.92  (0.69-1.23) |
| 5 (most affluent) | 0.90  (0.78-1.05) | 0.96  (0.87-1.07) | 0.84  (0.77-0.91) | 0.91  (0.84-0.98) | 0.75  (0.60-0.95) | 0.84  (0.67-1.06) | 0.89  (0.84-0.93) | 0.92  (0.88-0.96) | 0.87  (0.78-0.97) | 0.93  (0.85-1.03) | 0.92  (0.80-1.05) | 1.05  (0.95-1.16) | 1.14  (0.88-1.47) | 1.10  (0.82-1.47) |
| Unknown | 0.79  (0.51-1.23) | 0.90  (0.7-1.16) | 0.80  (0.66-0.98) | 0.92  (0.76-1.11) | 0.92  (0.52-1.62) | 0.97  (0.58-1.62) | 1.06  (0.94-1.19) | 0.96  (0.86-1.06) | 1.21  (0.95-1.53) | 1.05  (0.85-1.3) | 1.20  (0.90-1.60) | 1.04  (0.81-1.32) | 0.99  (0.53-1.84) | 1.34  (0.72-2.5) |
| **Season** |  |  |  |  |  |  |  |  |  |  |  |  |  |  |
| Spring | 0.95  (0.83-1.09) | 0.95  (0.87-1.04) | 0.92  (0.86-0.98) | 0.97  (0.91-1.03) | 0.85  (0.70-1.03) | 1.03  (0.85-1.25) | 0.93  (0.89-0.97) | 0.95  (0.91-0.98) | 0.88  (0.80-0.97) | 0.88  (0.81-0.95) | 1.14  (1.01-1.29) | 0.90  (0.83-0.99) | 1.10  (0.87-1.38) | 0.90  (0.68-1.19) |
| Summer | 1.07  (0.93-1.24) | 1.06  (0.96-1.16) | 0.90  (0.84-0.96) | 0.97  (0.9-1.04) | 0.87  (0.71-1.06) | 0.95  (0.77-1.19) | 0.97  (0.93-1.01) | 1.02  (0.98-1.06) | 0.97  (0.88-1.07) | 0.94  (0.86-1.03) | 1.30  (1.15-1.47) | 1.00  (0.92-1.10) | 1.26  (1.00-1.6) | 1.09  (0.83-1.44) |
| Winter | 0.9  (0.80-1.01) | 1.00  (0.92-1.08) | 1.01  (0.95-1.07) | 1.02  (0.96-1.09) | 1.00  (0.84-1.20) | 1.06  (0.88-1.28) | 1.06  (1.01-1.10) | 0.99  (0.96-1.03) | 1.02  (0.94-1.12) | 0.96  (0.89-1.03) | 1.05  (0.93-1.18) | 0.96  (0.88-1.04) | 1.16  (0.92-1.46) | 1.21  (0.93-1.58) |
| **Region** |  |  |  |  |  |  |  |  |  |  |  |  |  |  |
| London | 0.79  (0.62-1.02) | 0.76  (0.65-0.89) | 1.02  (0.88-1.19) | 0.97  (0.85-1.11) | 0.98  (0.64-1.49) | 0.60  (0.34-1.04) | 0.95  (0.86-1.05) | 0.98  (0.90-1.06) | 0.9  (0.74-1.09) | 0.73  (0.59-0.92) | 1.08  (0.88-1.33) | 0.99  (0.85-1.17) | 1.29  (0.90-1.85) | 1.20  (0.74-1.95) |
| North East | 1.09  (0.88-1.36) | 0.86  (0.73-1.02) | 0.99  (0.90-1.09) | 0.99  (0.89-1.10) | 1.04  (0.78-1.38) | 1.20  (0.89-1.62) | 1.03  (0.97-1.10) | 0.97  (0.91-1.03) | 1.02  (0.88-1.17) | 0.91  (0.79-1.04) | 1.19  (0.98-1.44) | 1.06  (0.91-1.23) | 1.06  (0.73-1.53) | 1.20  (0.77-1.89) |
| North West | 0.87  (0.74-1.03) | 0.97  (0.86-1.09) | 0.78  (0.72-0.85) | 0.99  (0.92-1.08) | 0.81  (0.64-1.03) | 0.96  (0.75-1.22) | 0.75  (0.71-0.80) | 1.01  (0.96-1.05) | 0.81  (0.72-0.91) | 0.94  (0.85-1.04) | 1.08  (0.93-1.26) | 1.02  (0.91-1.14) | 0.85  (0.62-1.15) | 1.29  (0.94-1.77) |
| West Midlands | 1.22  (0.98-1.52) | 0.97  (0.85-1.12) | 1.17  (1.03-1.31) | 1.11  (0.99-1.23) | 0.90  (0.62-1.29) | 1.14  (0.80-1.63) | 1.20  (1.11-1.29) | 1.08  (1.02-1.15) | 1.07  (0.90-1.26) | 1.10  (0.95-1.27) | 1.36  (1.12-1.65) | 1.19  (1.03-1.37) | 1.09  (0.74-1.61) | 1.20  (0.77-1.88) |
| Yorkshire and The Humber | 0.98  (0.84-1.14) | 0.98  (0.90-1.08) | 0.91  (0.85-0.98) | 1.02  (0.95-1.09) | 0.90  (0.73-1.11) | 1.10  (0.88-1.37) | 0.96  (0.91-1.01) | 1.00  (0.97-1.04) | 0.97  (0.87-1.07) | 0.95  (0.87-1.04) | 1.23  (1.07-1.40) | 1.05  (0.95-1.16) | 0.98  (0.76-1.26) | 1.00  (0.74-1.36) |
| South East | 0.97  (0.80-1.18) | 1.08  (0.94-1.25) | 0.76  (0.69-0.85) | 0.93  (0.84-1.03) | 0.95  (0.72-1.26) | 1.25  (0.95-1.64) | 0.92  (0.86-0.99) | 0.97  (0.92-1.03) | 0.87  (0.76-1.01) | 1.03  (0.91-1.17) | 1.15  (0.96-1.38) | 1.30  (1.14-1.48) | 1.02  (0.73-1.42) | 1.49  (1.04-2.14) |
| East Midlands | 1.08  (0.94-1.24) | 1.10  (1.00-1.20) | 1.06  (0.99-1.13) | 1.10  (1.02-1.17) | 1.01  (0.82-1.23) | 1.21  (0.98-1.49) | 1.09  (1.04-1.14) | 1.09  (1.05-1.13) | 1.07  (0.97-1.18) | 1.11  (1.02-1.21) | 1.14  (1.00-1.3) | 1.12  (1.02-1.23) | 1.01  (0.80-1.29) | 0.94  (0.70-1.25) |
| South West | 0.79  (0.67-0.95) | 1.02  (0.90-1.16) | 0.73  (0.67-0.79) | 0.95  (0.88-1.04) | 0.77  (0.59-1.00) | 1.22  (0.95-1.56) | 0.83  (0.79-0.88) | 1.05  (1.0-1.09) | 0.86  (0.76-0.97) | 1.07  (0.96-1.2) | 1.4  (1.21-1.61) | 1.37  (1.23-1.52) | 1.15  (0.87-1.50) | 1.49  (1.09-2.04) |
| **Flu vaccination** |  |  |  |  |  |  |  |  |  |  |  |  |  |  |
| Yes | 0.98  (0.87-1.09) | 1.00  (0.93-1.08) | 0.81  (0.77-0.85) | 1.05  (0.99-1.11) | 1.09  (0.92-1.28) | 1.03  (0.87-1.21) | 0.81  (0.78-0.84) | 0.97  (0.94-1.00) | 0.95  (0.88-1.02) | 0.99  (0.92-1.06) | 0.92  (0.82-1.04) | 1.04  (0.95-1.13) | 0.74  (0.57-0.96) | 0.97  (0.74-1.28) |
| **Count of antibiotic prescription in the one year before** | 1.20  (1.17-1.22) | 1.16  (1.14-1.18) | 1.15  (1.14-1.16) | 1.11  (1.10-1.12) | 1.17  (1.14-1.20) | 1.13  (1.10-1.16) | 1.12  (1.11-1.12) | 1.10  (1.10-1.11) | 1.11  (1.10-1.12) | 1.09  (1.08-1.11) | 1.18  (1.15-1.21) | 1.16  (1.14-1.19) | 1.06  (1.01-1.12) | 1.16  (1.09-1.24) |
| ^1^ HR, hazard ratio.  ^2^ CI, confidence interval.  ^3^ ABs, antibiotics prescribed or not.  ^4^ BMI, Body Mass Index recorded in the last 5 years.  ^5^ CCI, Charlson Comorbidities Index, measured from 17 weighted conditions, including myocardial infarction, congestive heart failure, peripheral vascular disease, cerebrovascular disease, dementia, chronic pulmonary disease, Connective tissue disease, ulcer disease, mild liver disease, diabetes, hemiplegia, moderate or severe renal disease, diabetes with complications, any malignancy (including leukaemia and lymphoma), moderate or severe liver disease, metastatic solid tumour, and AIDS.  ^6^ IMD, Multiple Deprivation Index, quintile measured from patient-level address.  Reference group for variable sex is female, for age is 18-25, for BMI is healthy weight, for ethnicity is non-white, for CCI is very low, for smoking status is ex-smoker, for IMD is 2, for season is autumn, for region is east, for flu vaccination is no. | | | | | | | | | | | | | | |

#### Hazard ratios of antibiotics

Supplementary Table 13. Crude hazard ratios of prescribed antibiotics as a predictor variable in Cox models for hospital admissions related to incident and prevalent other common infections, including sinusitis, otitis media, and otitis externa.

|  | **Sinusitis, crude HR^1^**  **(95% CI^2^)** | | **Otitis media, crude HR (95% CI)** | | **Otitis externa, crude HR (95% CI)** | |
| --- | --- | --- | --- | --- | --- | --- |
|  | **Incident** | **Prevalent** | **Incident** | **Prevalent** | **Incident** | **Prevalent** |
| **Antibiotic exposure** |  |  |  |  |  |  |
| No exposure | reference | reference | reference | reference | reference | reference |
| Exposed | 0.68  (0.60-0.77) | 0.69  (0.49-0.97) | 0.65  (0.58-0.72) | 0.68  (0.52-0.89) | 1.73  (1.61-1.86) | 1.00  (0.87-1.13) |
| **Antibiotic type^3^** |  |  |  |  |  |  |
| Most prescribed | reference | reference | reference | reference | reference | reference |
| Second most prescribed | 1.08  (0.91-1.28) | 1.12  (0.60-2.10) | 1.38  (3.20-5.09) | 1.58  (2.52-14.83) | 1.53  (1.32-1.78) | 1.22  (0.89-1.68) |
| Others | 1.73  (1.47-2.03) | 1.73  (0.97-3.09) | 1.85  (5.18-7.92) | 2.31  (4.48-35.28) | 0.82  (0.72-0.92) | 1.28  (0.98-1.68) |
| None | 1.45  (1.23-1.70) | 1.35  (0.74-2.44) | 1.85  (5.06-8.33) | 2.02  (3.62-24.04) | 1.69  (1.46-1.95) | 1.46  (1.09-1.97) |
| **Stratified by sex category** |  |  |  |  |  |  |
| Male | 0.66  (0.53-0.81) | 0.74  (0.41-1.36) | 0.61  (0.52-0.72) | - | 1.60  (1.43-1.80) | 0.96  (0.80-1.15) |
| Female | 0.67  (0.58-0.78) | 0.83  (0.55-1.25) | 0.70  (0.61-0.80) | 0.58  (0.41-0.82) | 1.77  (1.61-1.95) | 0.96  (0.79-1.16) |
| **Stratified by age category** |  |  |  |  |  |  |
| 18-24 | 0.69  (0.43-1.10) | 1.99  (0.04-107.40) | 1.01  (0.66-1.54) | - | 1.68  (1.28-2.21) | 1.06  (0.60-1.89) |
| 25-34 | 0.65  (0.47-0.89) | 0.35  (0.13-0.93) | 0.59  (0.46-0.77) | 0.94  (0.47-1.85) | 1.77  (1.46-2.14) | 1.08  (0.69-1.68) |
| 35-44 | 0.54  (0.40-0.73) | 0.79  (0.35-1.78) | - | - | 1.75  (1.40-2.18) | 0.67  (0.45-1.00) |
| 45-54 | 0.60  (0.44-0.82) | 0.62  (0.29-1.33) | 0.73  (0.54-0.97) | 0.42  (0.18-0.96) | 1.53  (1.25-1.85) | 0.68  (0.44-1.04) |
| 55-64 | 0.55  (0.41-0.73) | - | 0.55  (0.41-0.73) | - | 1.84  (1.50-2.25) | 1.23  (0.85-1.79) |
| 65-74 | 0.84  (0.61-1.16) | - | 0.68  (0.52-0.89) | - | 1.65  (1.37-1.98) | 1.23  (0.85-1.79) |
| 75+ | 0.68  (0.51-0.91) | 0.64  (0.27-1.49) | 0.74  (0.59-0.93) | 0.47  (0.26-0.85) | 1.62  (1.40-1.87) | 0.97  (0.78-1.19) |
| **Stratified by time** |  |  |  |  |  |  |
| Pre-pandemic | 0.68  (0.57-0.82) | - | 0.60  (0.51-0.70) | 0.73  (0.50-1.06) | 1.85  (1.65-2.06) | 0.89  (0.73-1.08) |
| Beginning and during pandemic | 0.46  (0.35-0.62) | - | 0.60  (0.47-0.76) | - | 1.46  (1.25-1.72) | 1.14  (0.85-1.52) |
| After 2^nd^ lockdown | 0.66  (0.57-0.76) | - | 0.60  (0.53-0.68) | 0.72  (0.53-0.99) | 1.69  (1.54-1.84) | 1.04  (0.89-1.21) |
| ^1^ HR, hazard ratio.  ^2^ CI, confidence interval.  ^3^ The most prescribed and the second most prescribed type of antibiotic are respectively amoxicillin and doxycycline for sinusitis, amoxicillin and clarithromycin for otitis media, and amoxicillin and flucloxacillin for otitis externa. | | | | | | |

Supplementary Table 14. Crude hazard ratios of prescribed antibiotics as a predictor variable in Cox models for hospital admissions related to incident and prevalent upper respiratory tract infections (URTI), including specific URTI, cough, cold with cough, and otitis externa.

|  | **Specific URTI,**  **crude HR^1^**  **(95% CI^2^)** | | **Cough, crude HR**  **(95% CI)** | | **Cold with cough, crude HR (95% CI)** | | **Sore throat, crude HR (95% CI)** | |
| --- | --- | --- | --- | --- | --- | --- | --- | --- |
|  | **Incident** | **Prevalent** | **Incident** | **Prevalent** | **Incident** | **Prevalent** | **Incident** | **Prevalent** |
| **Antibiotic exposure** |  |  |  |  |  |  |  |  |
| No exposure | reference | reference | reference | reference | reference | reference | reference | reference |
| Exposed | 0.79  (0.75-0.84) | 0.68  (0.55-0.84) | 1.35  (1.31-1.39) | 1.18  (1.07-1.30) | 0.90  (0.89-0.92) | 0.81  (0.78-0.85) | 0.79  (0.75-0.83) | 0.63  (0.56-0.72) |
| **Antibiotic type^3^** |  |  |  |  |  |  |  |  |
| Most prescribed | reference | reference | reference | reference | reference | reference | reference | reference |
| Second most prescribed | 0.98  (0.91-1.06) | 0.70  (0.51-0.96) | 1.00  (0.95-1.06) | 0.93  (0.79-1.11) | 0.98  (0.95-1.01) | 0.94  (0.87-1.01) | 0.86  (0.79-0.94) | 0.75  (0.61-0.93) |
| Others | 1.37  (1.29-1.46) | 1.39  (1.06-1.81) | 0.80  (0.77-0.84) | 0.91  (0.80-1.05) | 1.20  (1.17-1.22) | 1.34  (1.26-1.43) | 1.15  (1.05-1.26) | 1.39  (1.16-1.68) |
| None | 1.55  (1.44-1.66) | 1.18  (0.89-1.58) | 1.47  (1.39-1.55) | 1.36  (1.15-1.60) | 1.57  (1.52-1.61) | 1.38  (1.29-1.48) | 1.03  (0.93-1.15) | 1.09  (0.85-1.39) |
| **Stratified by sex category** |  |  |  |  |  |  |  |  |
| Male | 0.76  (0.70-0.83) | 0.66  (0.47-0.93) | 1.39  (1.32-1.46) | 1.27  (1.10-1.47) | 0.93  (0.91-0.96) | 0.81  (0.76-0.86) | 0.82  (0.75-0.89) | 0.69  (0.56-0.85) |
| Female | 0.84  (0.78-0.90) | 0.59  (0.45-0.76) | 1.34  (1.28-1.40) | 1.18  (1.04-1.35) | 0.87  (0.84-0.89) | 0.82  (0.77-0.87) | 0.82  (0.76-0.88) | 0.64  (0.55-0.74) |
| **Stratified by age category** |  |  |  |  |  |  |  |  |
| 18-24 | 0.90  (0.69-1.18) | 0.55  (0.20-1.47) | 0.98  (0.77-1.26) | 0.83  (0.33-2.08) | 0.74  (0.63-0.86) | 0.72  (0.47-1.09) | 0.68  (0.61-0.75) | 0.60  (0.49-0.73) |
| 25-34 | 0.85  (0.69-1.05) | 0.55  (0.20-1.47) | 1.06  (0.90-1.25) | 1.19  (0.73-1.95) | 0.73  (0.66-0.81) | 0.82  (0.64-1.05) | 0.74  (0.67-0.83) | 0.45  (0.35-0.56) |
| 35-44 | 0.70  (0.57-0.86) | 1.74  (0.59-5.10) | 1.03  (0.89-1.19) | 0.83  (0.51-1.35) | 0.71  (0.64-0.78) | 0.82  (0.65-1.04) | 0.81  (0.70-0.93) | 0.64  (0.43-0.94) |
| 45-54 | 0.81  (0.68-0.97) | 0.55  (0.27-1.12) | 1.25  (1.12-1.39) | 1.26  (0.94-1.68) | 0.79  (0.74-0.85) | 0.77  (0.66-0.90) | 0.93  (0.78-1.09) | 0.76  (0.47-1.21) |
| 55-64 | 0.86  (0.73-1.02) | 0.53  (0.26-1.06) | 1.25  (1.15-1.37) | - | 0.88  (0.83-0.93) | 0.87  (0.77-0.98) | 0.90  (0.74-1.08) | 0.83  (0.46-1.50) |
| 65-74 | 0.68  (0.59-0.77) | 0.51  (0.33-0.78) | 1.40  (1.31-1.49) | 1.40  (1.15-1.70) | 0.92  (0.88-0.96) | 0.78  (0.71-0.85) | 0.89  (0.73-1.08) | 1.90  (0.99-3.63) |
| 75+ | 0.86  (0.79-0.95) | 0.68  (0.50-0.93) | 1.41  (1.34-1.48) | 1.21  (1.04-1.41) | 0.90  (0.88-0.93) | 0.82  (0.77-0.87) | 1.09  (0.92-1.30) | 0.81  (0.40-1.63) |
| **Stratified by time** |  |  |  |  |  |  |  |  |
| Pre-pandemic | 0.91  (0.84-0.98) | 0.75  (0.57-0.99) | 1.41  (1.34-1.48) | 1.05  (0.91-1.22) | 0.88  (0.86-0.91) | 0.81  (0.76-0.86) | 0.93  (0.86-1.01) | 0.70  (0.59-0.85) |
| Beginning and during pandemic | 0.74  (0.62-0.88) | 0.64  (0.33-1.25) | 1.53  (1.41-1.65) | 1.49  (1.19-1.87) | 1.05  (1.00-1.10) | 0.96  (0.86-1.08) | 0.81  (0.71-0.92) | 0.64  (0.47-0.88) |
| After 2^nd^ lockdown | 0.89  (0.83-0.95) | 0.74  (0.58-0.93) | 1.46  (1.40-1.52) | 1.18  (1.05-1.32) | 0.94  (0.92-0.96) | 0.84  (0.80-0.88) | 0.90  (0.84-0.96) | 0.66  (0.57-0.76) |
| ^1^ HR, hazard ratio.  ^2^ CI, confidence interval.  ^3^ The most prescribed and the second most prescribed type of antibiotic are respectively amoxicillin and doxycycline for URTI, cough, and cold with cough, and phenoxymethylpenicillin and clarithromycin for sore throat. | | | | | | | | |

### Cox models with pre-pandemic data

Due to the fluctuations of counts of hospital admissions particularly related to UTI (shown in Error: Reference source not found), we split data into four periods regarding COVID-19 status (introduced in the Methods section). Supplementary Table 15 shows C-statistics of all converged Cox models without extreme HRs using development and validation splits of pre-pandemic data for hospital admission related to common infectionsError: Reference source not found. The models with pre-pandemic data showed strong association in age category 75+ and CCI category very high with infection-related complication. Supplementary Tables 16-19 present HRs of Cox models for infection-related hospital admissions using pre-pandemic data. The calibration plots of these Cox models are displayed in Supplementary Figures 1-10. These figures indicate that models for LRTI, URTI, and UTI were well-calibrated, but those for other common infections were less well calibrated.

#### Performance

Supplementary Table 15. C-statistics of Cox models for hospital admissions related to common infections, including lower respiratory tract infection, upper respiratory tract infection, urinary tract infection (UTI), sinusitis, otitis media, and otitis externa, using pre-pandemic data (from January 2019 to December 2019).

|  | | | **C-statistics** | |
| --- | --- | --- | --- | --- |
|  |  |  | **Development dataset** | **Validation dataset** |
| LRTI | Incident | No ABs^1^ | 0.68 | 0.67 |
|  |  | With ABs | 0.72 | 0.72 |
|  | Prevalent | No ABs | 0.67 | 0.67 |
|  |  | With ABs | 0.68 | 0.67 |
| URTI | Incident | No ABs | 0.73 | 0.72 |
|  |  | With ABs | 0.71 | 0.71 |
|  | Prevalent | No ABs | 0.71 | 0.72 |
|  |  | With ABs | 0.69 | 0.68 |
| UTI | Incident | No ABs | 0.73 | 0.73 |
|  |  | With ABs | 0.75 | 0.75 |
|  | Prevalent | No ABs | 0.70 | 0.71 |
|  |  | With ABs | 0.69 | 0.70 |
| Sinusitis | Incident | No ABs | 0.73 | 0.64 |
|  |  | With ABs | - | - |
|  | Prevalent | No ABs | - | - |
|  |  | With ABs | - | - |
| Otitis media | Incident | No ABs | 0.71 | 0.64 |
|  |  | With ABs | 0.67 | 0.68 |
|  | Prevalent | No ABs | - | - |
|  |  | With ABs | - | - |
| Otitis externa | Incident | No ABs | 0.71 | 0.70 |
|  |  | With ABs | - | - |
|  | Prevalent | No ABs | 0.76 | 0.72 |
|  |  | With ABs | - | - |
| URTI components | |  |  |  |
| Specific URTI | Incident | No ABs | 0.75 | 0.76 |
|  |  | With ABs | 0.72 | 0.73 |
|  | Prevalent | No ABs | - | - |
|  |  | With ABs | - | - |
| Cough | Incident | No ABs | 0.73 | 0.73 |
|  |  | With ABs | 0.71 | 0.71 |
|  | Prevalent | No ABs | 0.75 | 0.70 |
|  |  | With ABs | 0.70 | 0.68 |
| Cold with cough | Incident | No ABs | 0.74 | 0.74 |
|  |  | With ABs | 0.71 | 0.71 |
|  | Prevalent | No ABs | 0.72 | 0.71 |
|  |  | With ABs | 0.68 | 0.69 |
| Sore throat | Incident | No ABs | 0.64 | 0.63 |
|  |  | With ABs | 0.62 | 0.59 |
|  | Prevalent | No ABs | 0.67 | 0.61 |
|  |  | With ABs | 0.68 | 0.67 |
| ^1^ABs, antibiotics prescribed or not. | | | | |

#### Hazard ratios

Supplementary Table 16. Adjusted hazard ratios of Cox models for hospital admissions related to incident common infections, including lower respiratory tract infection (LRTI), upper respiratory tract infection (URTI), and urinary tract infection (UTI), using pre-pandemic data (from January 2019 to December 2019).

|  | **LRTI, adjusted HR^1^ (95% CI^2^)** | **URTI, adjusted HR**  **(95% CI)** | **UTI, adjusted HR (95% CI)** |
| --- | --- | --- | --- |
| **Sex** |  |  |  |
| Male | 1.17 (1.10-1.23) | 1.14 (1.10-1.19) | 1.60 (1.51-1.71) |
| **Age** |  |  |  |
| 25-34 | 1.13 (0.86-1.48) | 0.98 (0.87-1.09) | 0.82 (0.65-1.04) |
| 35-44 | 1.31 (1.01-1.70) | 0.89 (0.79-1.00) | 0.99 (0.78-1.25) |
| 45-54 | 1.42 (1.10-1.82) | 0.93 (0.83-1.04) | 1.39 (1.12-1.72) |
| 55-64 | 1.97 (1.55-2.51) | 1.14 (1.02-1.26) | 1.72 (1.40-2.10) |
| 65-74 | 2.97 (2.34-3.77) | 1.66 (1.50-1.84) | 2.93 (2.42-3.54) |
| 75+ | 4.63 (3.67-5.86) | 3.63 (3.28-4.01) | 5.22 (4.35-6.27) |
| **BMI^3^** |  |  |  |
| Underweight | 1.16 (1.01-1.33) | 1.45 (1.32-1.60) | 1.26 (1.06-1.49) |
| Overweight | 0.89 (0.82-0.96) | 0.82 (0.78-0.87) | 0.90 (0.83-0.97) |
| Obese | 0.99 (0.92-1.06) | 0.91 (0.86-0.95) | 1.15 (1.06-1.25) |
| Unknown | 1.14 (1.05-1.24) | 1.27 (1.20-1.35) | 1.16 (1.06-1.27) |
| **Ethnicity** |  |  |  |
| White | 0.96 (0.84-1.09) | 0.91 (0.84-0.99) | 1.21 (1.03-1.41) |
| Unknown | 1.01 (0.88-1.15) | 1.05 (0.97-1.14) | 1.29 (1.09-1.51) |
| **CCI^4^** |  |  |  |
| Low | 1.26 (1.18-1.35) | 1.44 (1.38-1.50) | 1.35 (1.26-1.45) |
| Medium | 1.54 (1.42-1.67) | 2.05 (1.94-2.18) | 1.55 (1.41-1.70) |
| High | 1.65 (1.44-1.89) | 2.72 (2.48-2.98) | 2.00 (1.75-2.29) |
| Very high | 2.14 (1.78-2.58) | 3.47 (3.02-3.98) | 2.48 (2.05-3.00) |
| **Smoking status** |  |  |  |
| Smoker | 1.14 (1.05-1.24) | 0.99 (0.94-1.05) | 1.20 (1.08-1.33) |
| Never smoked | 0.95 (0.90-1.02) | 1.08 (1.03-1.13) | 1.00 (0.94-1.07) |
| Unknown | 1.03 (0.62-1.69) | 0.97 (0.72-1.3) | 0.89 (0.51-1.54) |
| **IMD^5^** |  |  |  |
| 1 (most deprived) | 1.13 (1.04-1.22) | 1.04 (0.99-1.10) | 1.08 (0.99-1.18) |
| 3 | 0.93 (0.86-1.01) | 0.86 (0.82-0.91) | 0.90 (0.82-0.98) |
| 4 | 0.92 (0.85-1.01) | 0.91 (0.86-0.96) | 0.93 (0.85-1.02) |
| 5 (most affluent) | 0.84 (0.77-0.92) | 0.80 (0.76-0.86) | 0.77 (0.70-0.85) |
| Unknown | 1.19 (0.97-1.47) | 0.96 (0.82-1.11) | 0.97 (0.76-1.23) |
| **Season** |  |  |  |
| Spring | 0.91 (0.84-0.98) | 0.99 (0.94-1.05) | 0.90 (0.83-0.98) |
| Summer | 1.03 (0.95-1.11) | 0.94 (0.89-1.00) | 0.96 (0.88-1.04) |
| Winter | 1.00 (0.93-1.08) | 1.09 (1.04-1.14) | 1.02 (0.94-1.10) |
| **Region** |  |  |  |
| London | 0.77 (0.64-0.91) | 0.91 (0.81-1.02) | 1.02 (0.86-1.21) |
| North East | 0.99 (0.88-1.12) | 0.88 (0.81-0.96) | 1.00 (0.88-1.15) |
| North West | 0.83 (0.75-0.92) | 0.78 (0.73-0.83) | 0.84 (0.75-0.94) |
| West Midlands | 0.92 (0.80-1.06) | 1.09 (0.99-1.19) | 1.01 (0.86-1.18) |
| Yorkshire and The Humber | 0.92 (0.84-1.00) | 0.95 (0.90-1.01) | 1.09 (1.00-1.20) |
| South East | 1.00 (0.89-1.13) | 0.91 (0.84-0.98) | 0.90 (0.79-1.02) |
| East Midlands | 1.11 (1.02-1.20) | 1.05 (1.00-1.11) | 1.07 (0.98-1.17) |
| South West | 0.86 (0.78-0.95) | 0.82 (0.76-0.87) | 0.77 (0.69-0.86) |
| **Flu vaccination** |  |  |  |
| Yes | 0.92 (0.87-0.98) | 0.81 (0.78-0.85) | 0.88 (0.83-0.94) |
| **Count of antibiotic prescription in the one year before** | 1.04 (1.03-1.05) | 1.15 (1.14-1.16) | 1.02 (1.01-1.03) |
| ^1^ HR, hazard ratio.  ^2^ CI, confidence interval.  ^3^ BMI, Body Mass Index recorded in the last 5 years.  ^4^ CCI, Charlson Comorbidities Index, measured from 17 weighted conditions, including myocardial infarction, congestive heart failure, peripheral vascular disease, cerebrovascular disease, dementia, chronic pulmonary disease, Connective tissue disease, ulcer disease, mild liver disease, diabetes, hemiplegia, moderate or severe renal disease, diabetes with complications, any malignancy (including leukaemia and lymphoma), moderate or severe liver disease, metastatic solid tumour, and AIDS.  ^5^ IMD, Multiple Deprivation Index, quintile measured from patient-level address.  Reference group for variable sex is female, for age is 18-25, for BMI is healthy weight, for ethnicity is non-white, for CCI is very low, for smoking status is ex-smoker, for IMD is 2, for season is autumn, for region is east, for flu vaccination is no. | | | |

Supplementary Table 17. Adjusted hazard ratios of Cox models for hospital admissions related to common infections, including lower respiratory tract infection (LRTI), upper respiratory tract infection (URTI), urinary tract infection (UTI), sinusitis, otitis media, and otitis externa, using pre-pandemic data (from January 2019 to December 2019).

|  | **LRTI, adjusted HR (95% CI)** | | | **URTI, adjusted HR (95% CI)** | | | **UTI, adjusted HR (95% CI)** | | |
| --- | --- | --- | --- | --- | --- | --- | --- | --- | --- |
|  | **Incident** | **Prevalent** | | **Incident** | **Prevalent** | | **Incident** | **Prevalent** | |
|  | **With ABs^1^** | **No ABs** | **With ABs** | **No ABs** | **With ABs^1^** | **No ABs** | **With ABs** | **No ABs** | **With ABs^1^** |
| **Sex** |  |  |  |  |  |  |  |  |  |
| Male | 1.12  (1.09-1.16) | 1.17  (1.06-1.3) | 1.21  (1.11-1.31) | 1.13  (1.1-1.17) | 1.14  (1.06-1.24) | 1.23  (1.15-1.32) | 2.15  (2.06-2.23) | 1.74  (1.56-1.94) | 1.95  (1.79-2.11) |
| **Age** |  |  |  |  |  |  |  |  |  |
| 25-34 | 1.06  (0.9-1.24) | 1.52  (0.81-2.86) | 0.81  (0.47-1.38) | 0.86  (0.79-0.93) | 0.63  (0.5-0.79) | 0.58  (0.45-0.73) | 1.04  (0.9-1.19) | 1.03  (0.68-1.56) | 1.16  (0.83-1.62) |
| 35-44 | 1.03  (0.88-1.2) | 1.23  (0.66-2.29) | 0.87  (0.53-1.43) | 0.75  (0.69-0.82) | 0.46  (0.36-0.58) | 0.4  (0.32-0.52) | 1.01  (0.88-1.17) | 0.89  (0.57-1.38) | 1.23  (0.88-1.71) |
| 45-54 | 1.12  (0.96-1.3) | 1.26  (0.69-2.31) | 1.14  (0.71-1.84) | 0.81  (0.75-0.88) | 0.53  (0.43-0.66) | 0.59  (0.48-0.73) | 1.16  (1.02-1.32) | 1.0  (0.67-1.5) | 1.13  (0.82-1.55) |
| 55-64 | 1.41  (1.22-1.63) | 1.57  (0.87-2.84) | 1.35  (0.85-2.16) | 0.99  (0.91-1.06) | 0.5  (0.41-0.62) | 0.64  (0.53-0.79) | 1.48  (1.31-1.68) | 1.12  (0.76-1.64) | 1.27  (0.94-1.72) |
| 65-74 | 1.91  (1.66-2.21) | 2.61  (1.46-4.68) | 1.77  (1.12-2.82) | 1.31  (1.22-1.42) | 0.77  (0.63-0.94) | 0.88  (0.73-1.07) | 2.13  (1.89-2.41) | 1.82  (1.27-2.61) | 1.69  (1.26-2.27) |
| 75+ | 3.96  (3.43-4.57) | 3.58  (2.01-6.39) | 2.83  (1.79-4.49) | 2.75  (2.56-2.96) | 1.32  (1.09-1.6) | 1.44  (1.19-1.73) | 4.41  (3.92-4.96) | 2.77  (1.95-3.94) | 3.04  (2.28-4.06) |
| **BMI^2^** |  |  |  |  |  |  |  |  |  |
| Underweight | 1.46  (1.34-1.6) | 1.24  (0.96-1.6) | 1.38  (1.12-1.69) | 1.48  (1.37-1.6) | 1.5  (1.22-1.85) | 1.37  (1.13-1.66) | 1.41  (1.26-1.58) | 1.21  (0.86-1.7) | 1.2  (0.93-1.55) |
| Overweight | 0.8  (0.77-0.84) | 0.93  (0.81-1.06) | 0.73  (0.66-0.82) | 0.82  (0.79-0.86) | 0.87  (0.78-0.97) | 0.8  (0.73-0.88) | 0.91  (0.87-0.96) | 0.99  (0.86-1.14) | 0.95  (0.85-1.05) |
| Obese | 0.85  (0.81-0.89) | 0.83  (0.72-0.95) | 0.83  (0.75-0.92) | 0.89  (0.86-0.93) | 0.91  (0.82-1.02) | 0.85  (0.78-0.94) | 1.03  (0.98-1.08) | 1.19  (1.04-1.37) | 0.97  (0.87-1.07) |
| Unknown | 1.03  (0.98-1.09) | 1.1  (0.93-1.29) | 1.01  (0.89-1.15) | 1.06  (1.02-1.11) | 1.31  (1.16-1.47) | 1.09  (0.97-1.22) | 1.08  (1.02-1.14) | 1.23  (1.04-1.45) | 1.08  (0.96-1.22) |
| **Ethnicity** |  |  |  |  |  |  |  |  |  |
| White | 1.02  (0.94-1.1) | 0.94  (0.73-1.22) | 1.06  (0.87-1.3) | 1.07  (1.01-1.14) | 1.06  (0.88-1.27) | 1.06  (0.89-1.25) | 1.02  (0.93-1.12) | 0.85  (0.65-1.1) | 0.87  (0.72-1.06) |
| Unknown | 1.13  (1.05-1.23) | 1.12  (0.86-1.46) | 1.24  (1.01-1.53) | 1.19  (1.12-1.27) | 1.22  (1.01-1.46) | 1.16  (0.98-1.38) | 1.14  (1.04-1.25) | 1.0  (0.77-1.31) | 0.96  (0.78-1.17) |
| **CCI^3^** |  |  |  |  |  |  |  |  |  |
| Low | 1.32  (1.27-1.37) | 1.18  (1.04-1.34) | 1.24  (1.13-1.37) | 1.3  (1.26-1.35) | 1.48  (1.35-1.63) | 1.31  (1.21-1.42) | 1.49  (1.43-1.56) | 1.41  (1.24-1.61) | 1.47  (1.34-1.61) |
| Medium | 1.73  (1.64-1.82) | 1.63  (1.39-1.9) | 1.67  (1.48-1.88) | 1.76  (1.68-1.84) | 2.0  (1.77-2.26) | 1.7  (1.53-1.89) | 1.81  (1.71-1.92) | 2.03  (1.75-2.37) | 1.74  (1.54-1.96) |
| High | 2.27  (2.09-2.47) | 1.78  (1.4-2.28) | 1.82  (1.49-2.21) | 2.1  (1.95-2.27) | 2.14  (1.75-2.6) | 2.14  (1.81-2.54) | 2.22  (2.03-2.43) | 1.9  (1.51-2.4) | 2.1  (1.76-2.52) |
| Very high | 2.66  (2.33-3.04) | 2.21  (1.59-3.07) | 2.74  (2.09-3.59) | 2.85  (2.55-3.19) | 2.44  (1.8-3.31) | 3.09  (2.42-3.93) | 2.77  (2.4-3.19) | 2.19  (1.55-3.12) | 2.62  (1.97-3.47) |
| **Smoking** |  |  |  |  |  |  |  |  |  |
| Smoker | 0.89  (0.84-0.94) | 0.98  (0.83-1.17) | 0.9  (0.79-1.03) | 0.93  (0.9-0.97) | 0.92  (0.82-1.05) | 0.85  (0.77-0.95) | 1.2  (1.12-1.27) | 1.09  (0.9-1.32) | 1.25  (1.09-1.43) |
| Never smoked | 0.98  (0.95-1.02) | 0.9  (0.8-1.01) | 1.0  (0.91-1.09) | 0.98  (0.95-1.02) | 0.92  (0.84-1.0) | 0.96  (0.88-1.04) | 0.99  (0.95-1.03) | 0.92  (0.82-1.03) | 0.99  (0.91-1.07) |
| Unknown | 0.93  (0.65-1.35) | 0.32  (0.04-2.28) | 1.1  (0.41-2.96) | 1.01  (0.83-1.23) | 1.16  (0.69-1.96) | 1.26  (0.76-2.09) | 0.92  (0.63-1.35) | 0.74  (0.28-2.0) | 0.64  (0.21-2.0) |
| **IMD^4^** |  |  |  |  |  |  |  |  |  |
| 1 (most deprived) | 1.05  (1.0-1.11) | 1.26  (1.08-1.47) | 1.13  (1.01-1.28) | 1.02  (0.98-1.07) | 1.17  (1.04-1.32) | 1.07  (0.96-1.19) | 1.08  (1.02-1.14) | 1.09  (0.92-1.28) | 1.03  (0.91-1.16) |
| 3 | 0.97  (0.92-1.03) | 0.88  (0.75-1.04) | 1.03  (0.91-1.16) | 0.98  (0.94-1.02) | 0.91  (0.8-1.02) | 1.0  (0.9-1.11) | 0.91  (0.86-0.96) | 0.89  (0.76-1.04) | 0.86  (0.77-0.97) |
| 4 | 0.97  (0.92-1.02) | 0.8  (0.68-0.95) | 0.99  (0.88-1.12) | 0.99  (0.95-1.04) | 0.97  (0.86-1.1) | 0.98  (0.88-1.09) | 0.86  (0.82-0.91) | 0.76  (0.64-0.89) | 0.77  (0.68-0.87) |
| 5 (most affluent) | 0.89  (0.84-0.94) | 0.9  (0.76-1.06) | 0.98  (0.86-1.12) | 0.92  (0.87-0.96) | 0.87  (0.77-1.0) | 0.97  (0.87-1.09) | 0.83  (0.78-0.88) | 0.69  (0.58-0.82) | 0.73  (0.65-0.83) |
| Unknown | 0.84  (0.72-0.97) | 1.23  (0.83-1.82) | 0.97  (0.69-1.35) | 0.95  (0.85-1.06) | 1.21  (0.9-1.61) | 1.07  (0.82-1.39) | 0.96  (0.83-1.11) | 1.02  (0.69-1.52) | 0.93  (0.69-1.26) |
| **Season** |  |  |  |  |  |  |  |  |  |
| Spring | 0.94  (0.9-0.99) | 0.94  (0.81-1.09) | 0.87  (0.78-0.98) | 0.97  (0.94-1.01) | 0.93  (0.83-1.04) | 0.88  (0.8-0.97) | 0.96  (0.91-1.01) | 1.02  (0.89-1.18) | 0.95  (0.86-1.06) |
| Summer | 0.98  (0.93-1.03) | 1.02  (0.87-1.19) | 0.85  (0.76-0.96) | 0.98  (0.94-1.02) | 0.91  (0.81-1.02) | 0.93  (0.83-1.03) | 0.98  (0.93-1.03) | 1.01  (0.87-1.17) | 0.94  (0.84-1.04) |
| Winter | 0.94  (0.9-0.98) | 0.91  (0.79-1.04) | 0.86  (0.78-0.95) | 0.95  (0.92-0.98) | 0.99  (0.9-1.1) | 0.94  (0.86-1.03) | 0.95  (0.9-0.99) | 1.02  (0.88-1.17) | 0.95  (0.85-1.05) |
| **Region** |  |  |  |  |  |  |  |  |  |
| London | 1.01  (0.9-1.12) | 1.06  (0.75-1.49) | 0.95  (0.69-1.32) | 0.89  (0.82-0.97) | 1.17  (0.91-1.49) | 0.78  (0.59-1.03) | 1.07  (0.95-1.19) | 0.96  (0.7-1.32) | 1.1  (0.83-1.45) |
| North East | 0.97  (0.9-1.06) | 0.99  (0.79-1.24) | 0.88  (0.73-1.06) | 0.97  (0.91-1.04) | 0.86  (0.72-1.03) | 0.95  (0.81-1.12) | 1.01  (0.92-1.11) | 0.96  (0.74-1.24) | 1.1  (0.91-1.33) |
| North West | 1.02  (0.96-1.08) | 0.88  (0.73-1.05) | 0.9  (0.78-1.03) | 1.06  (1.01-1.11) | 0.79  (0.69-0.91) | 0.93  (0.82-1.05) | 1.0  (0.94-1.07) | 0.85  (0.7-1.03) | 0.94  (0.82-1.08) |
| West Midlands | 1.11  (1.02-1.21) | 1.04  (0.81-1.34) | 0.94  (0.76-1.16) | 1.13  (1.06-1.21) | 1.14  (0.95-1.38) | 1.21  (1.03-1.44) | 0.96  (0.87-1.05) | 1.09  (0.83-1.43) | 1.02  (0.83-1.26) |
| Yorkshire and The Humber | 0.97  (0.92-1.02) | 0.89  (0.76-1.04) | 0.87  (0.77-0.99) | 1.03  (0.98-1.07) | 0.99  (0.87-1.11) | 0.95  (0.85-1.06) | 1.05  (0.99-1.11) | 0.95  (0.81-1.12) | 1.12  (1.0-1.26) |
| South East | 1.02  (0.95-1.1) | 0.87  (0.68-1.1) | 0.91  (0.76-1.1) | 0.99  (0.93-1.06) | 0.9  (0.76-1.07) | 1.07  (0.92-1.25) | 0.91  (0.84-0.98) | 0.67  (0.51-0.88) | 0.89  (0.74-1.07) |
| East Midlands | 1.08  (1.03-1.14) | 0.97  (0.83-1.14) | 1.07  (0.95-1.2) | 1.12  (1.07-1.17) | 1.08  (0.96-1.21) | 1.11  (1.0-1.23) | 0.95  (0.9-1.0) | 1.13  (0.97-1.32) | 0.99  (0.88-1.11) |
| South West | 1.02  (0.96-1.09) | 0.89  (0.73-1.08) | 0.97  (0.83-1.13) | 1.05  (1.0-1.11) | 0.83  (0.72-0.97) | 1.02  (0.9-1.16) | 0.84  (0.79-0.9) | 0.95  (0.79-1.15) | 0.92  (0.8-1.07) |
| **Flu vaccination** |  |  |  |  |  |  |  |  |  |
| Yes | 0.95  (0.91-0.99) | 0.92  (0.81-1.03) | 0.96  (0.87-1.05) | 0.96  (0.92-0.99) | 0.92  (0.84-1.01) | 0.97  (0.89-1.05) | 0.9  (0.86-0.94) | 0.9  (0.8-1.02) | 0.9  (0.82-0.99) |
| **Count of antibiotic prescription in the one year before** | 1.11  (1.1-1.11) | 1.05  (1.03-1.07) | 1.09  (1.07-1.1) | 1.13  (1.12-1.14) | 1.14  (1.13-1.16) | 1.11  (1.1-1.13) | 1.05  (1.05-1.06) | 1.03  (1.02-1.04) | 1.03  (1.02-1.04) |
| ^1^ HR, hazard ratio.  ^2^ CI, confidence interval.  ^3^ ABs, antibiotics prescribed or not.  ^4^ BMI, Body Mass Index recorded in the last 5 years.  ^5^ CCI, Charlson Comorbidities Index, measured from 17 weighted conditions, including myocardial infarction, congestive heart failure, peripheral vascular disease, cerebrovascular disease, dementia, chronic pulmonary disease, Connective tissue disease, ulcer disease, mild liver disease, diabetes, hemiplegia, moderate or severe renal disease, diabetes with complications, any malignancy (including leukaemia and lymphoma), moderate or severe liver disease, metastatic solid tumour, and AIDS.  ^6^ IMD, Multiple Deprivation Index, quintile measured from patient-level address.  Reference group for variable sex is female, for age is 18-25, for BMI is healthy weight, for ethnicity is non-white, for CCI is very low, for smoking status is ex-smoker, for IMD is 2, for season is autumn, for region is east, for flu vaccination is no. | | | | | | | | | |

Supplementary Table 18. Adjusted hazard ratios of Cox models for hospital admissions related to other common infections, including sinusitis, otitis media, and otitis externa, using pre-pandemic data (from January 2019 to December 2019).

|  | **Sinusitis, adjusted HR^1^ (95% CI^2^)** | **Otitis media, adjusted HR (95% CI)** | | **Otitis externa, adjusted HR (95% CI)** | |
| --- | --- | --- | --- | --- | --- |
|  | **Incident** | **Incident** | | **Incident** | **Prevalent** |
|  | **No ABs^3^** | **No ABs** | **With ABs** | **No ABs** | **No ABs** |
| **Sex** |  |  |  |  |  |
| Male | 1.41  (1.03-1.95) | 1.41  (1.08-1.83) | 1.17  (0.97-1.42) | 1.18  (1.04-1.35) | 1.36  (1.08-1.71) |
| **Age** |  |  |  |  |  |
| 25-34 | 0.75  (0.38-1.48) | 1.49  (0.80-2.79) | 1.06  (0.75-1.51) | 1.05  (0.75-1.47) | 0.59  (0.32-1.10) |
| 35-44 | 0.72  (0.36-1.42) | 1.26  (0.66-2.40) | 0.86  (0.59-1.24) | 1.04  (0.74-1.45) | 0.68  (0.37-1.24) |
| 45-54 | 0.59  (0.29-1.19) | 1.09  (0.57-2.10) | 0.76  (0.51-1.11) | 1.03  (0.74-1.43) | 0.65  (0.36-1.16) |
| 55-64 | 0.81  (0.41-1.60) | 1.40  (0.74-2.65) | 0.78  (0.53-1.16) | 0.85  (0.61-1.19) | 0.83  (0.47-1.47) |
| 65-74 | 0.41  (0.18-0.91) | 1.21  (0.62-2.38) | 1.02  (0.68-1.54) | 1.03  (0.73-1.45) | 0.78  (0.43-1.42) |
| 75+ | 1.74  (0.83-3.62) | 2.36  (1.23-4.52) | 1.99  (1.33-2.98) | 2.34  (1.69-3.25) | 2.88  (1.66-4.98) |
| **BMI**^2^ |  |  |  |  |  |
| Underweight | 1.51  (0.54-4.21) | 0.95  (0.30-3.08) | 0.88  (0.38-2.0) | 1.19  (0.72-1.99) | 0.47  (0.12-1.94) |
| Overweight | 0.71  (0.46-1.10) | 1.21  (0.83-1.75) | 0.81  (0.62-1.05) | 0.88  (0.73-1.06) | 0.97  (0.71-1.33) |
| Obese | 0.87  (0.57-1.34) | 1.26  (0.87-1.82) | 1.08  (0.84-1.38) | 1.12  (0.94-1.34) | 1.29  (0.95-1.75) |
| Unknown | 1.12  (0.71-1.77) | 1.21  (0.80-1.83) | 0.90  (0.68-1.19) | 0.99  (0.81-1.22) | 0.76  (0.51-1.15) |
| **Ethnicity** |  |  |  |  |  |
| White | 0.76  (0.43-1.34) | 0.87  (0.53-1.42) | 1.12  (0.80-1.58) | 1.00  (0.77-1.29) | 1.23  (0.72-2.12) |
| Unknown | 0.71  (0.39-1.30) | 0.78  (0.47-1.29) | 1.06  (0.74-1.51) | 1.09  (0.84-1.43) | 1.62  (0.94-2.82) |
| **CCI**^3^ |  |  |  |  |  |
| Low | 1.21  (0.84-1.74) | 1.55  (1.15-2.09) | 1.34  (1.09-1.66) | 1.56  (1.34-1.81) | 1.60  (1.23-2.08) |
| Medium | 2.03  (1.12-3.68) | 1.96  (1.23-3.14) | 2.01  (1.43-2.83) | 2.76  (2.24-3.40) | 2.58  (1.84-3.61) |
| High | 2.10  (0.64-6.86) | 2.32  (0.99-5.42) | 1.92  (0.94-3.94) | 3.50  (2.46-4.98) | 2.84  (1.64-4.91) |
| Very high | 2.50  (0.34-18.29) | 1.20  (0.17-8.74) | 4.50  (1.84-11.03) | 4.61  (2.58-8.24) | 1.51  (0.37-6.20) |
| **Smoking** |  |  |  |  |  |
| Smoker | 0.85  (0.53-1.36) | 1.42  (0.99-2.02) | 0.98  (0.76-1.28) | 1.05  (0.86-1.27) | 1.03  (0.73-1.45) |
| Never smoked | 0.72  (0.51-1.02) | 1.04  (0.77-1.39) | 1.12  (0.91-1.36) | 0.87  (0.76-1.01) | 0.86  (0.66-1.11) |
| Unknown | 1.72  (0.39-7.46) | 0.80  (0.11-6.06) | 0.27  (0.04-1.94) | 0.38  (0.09-1.53) | 0.71  (0.09-5.29) |
| **IMD**^4^ |  |  |  |  |  |
| 1 (most deprived) | 1.50  (0.91-2.47) | 0.84  (0.57-1.23) | 1.10  (0.84-1.42) | 1.16  (0.95-1.41) | 1.49  (1.07-2.08) |
| 3 | 1.18  (0.71-1.96) | 0.88  (0.60-1.31) | 1.06  (0.81-1.40) | 1.15  (0.94-1.40) | 0.86  (0.59-1.25) |
| 4 | 1.00  (0.59-1.71) | 0.9  (0.60-1.34) | 0.92  (0.68-1.24) | 1.04  (0.85-1.28) | 0.90  (0.63-1.29) |
| 5 (most affluent) | 1.26  (0.75-2.12) | 0.89  (0.59-1.34) | 1.02  (0.75-1.38) | 1.00  (0.80-1.24) | 0.93  (0.64-1.35) |
| Unknown | 0.80  (0.19-3.41) | 1.47  (0.63-3.44) | 1.13  (0.57-2.24) | 1.34  (0.83-2.16) | 1.03  (0.37-2.86) |
| **Season** |  |  |  |  |  |
| Spring | 1.71  (1.09-2.67) | 0.70  (0.49-0.99) | 0.78  (0.61-0.99) | 0.93  (0.78-1.1) | 0.73  (0.54-0.98) |
| Summer | 1.38  (0.84-2.27) | 0.77  (0.55-1.09) | 0.84  (0.66-1.07) | 0.92  (0.78-1.09) | 0.75  (0.56-0.99) |
| Winter | 1.40  (0.89-2.19) | 0.74  (0.53-1.04) | 0.76  (0.6-0.97) | 0.86  (0.72-1.02) | 0.75  (0.55-1.00) |
| **Region** |  |  |  |  |  |
| London | 1.23  (0.55-2.74) | 0.35  (0.12-0.98) | 1.06  (0.62-1.82) | 1.37  (0.98-1.92) | 2.12  (1.12-4.02) |
| North East | 1.38  (0.69-2.76) | 1.36  (0.80-2.31) | 1.21  (0.77-1.9) | 1.17  (0.86-1.58) | 2.21  (1.39-3.53) |
| North West | 1.01  (0.57-1.79) | 0.88  (0.55-1.4) | 1.40  (1.02-1.94) | 1.14  (0.90-1.45) | 1.47  (0.95-2.29) |
| West Midlands | 0.99  (0.41-2.41) | 1.12  (0.60-2.12) | 1.45  (0.94-2.26) | 1.19  (0.84-1.68) | 1.79  (0.95-3.35) |
| Yorkshire and The Humber | 1.42  (0.88-2.30) | 0.81  (0.53-1.24) | 1.33  (0.99-1.77) | 1.20  (0.97-1.47) | 1.53  (1.03-2.27) |
| South East | 1.25  (0.66-2.39) | 1.03  (0.56-1.87) | 0.99  (0.62-1.61) | 0.97  (0.71-1.31) | 1.32  (0.77-2.26) |
| East Midlands | 0.84  (0.48-1.46) | 1.33  (0.92-1.91) | 1.47  (1.12-1.93) | 1.20  (0.99-1.47) | 1.95  (1.35-2.83) |
| South West | 1.41  (0.82-2.40) | 1.01  (0.62-1.63) | 1.62  (1.17-2.25) | 1.32  (1.06-1.65) | 1.70  (1.12-2.58) |
| **Flu vaccination** |  |  |  |  |  |
| Yes | 1.42  (0.95-2.10) | 1.35  (0.98-1.87) | 1.09  (0.87-1.36) | 1.07  (0.91-1.26) | 0.70  (0.54-0.92) |
| **Count of antibiotic prescription in the one year before** | 1.33  (1.24-1.44) | 1.27  (1.19-1.36) | 1.22  (1.16-1.28) | 1.27  (1.22-1.32) | 1.23  (1.16-1.31) |
| ^1^ HR, hazard ratio.  ^2^ CI, confidence interval.  ^3^ ABs, antibiotics prescribed or not.  ^4^ BMI, Body Mass Index recorded in the last 5 years.  ^5^ CCI, Charlson Comorbidities Index, measured from 17 weighted conditions, including myocardial infarction, congestive heart failure, peripheral vascular disease, cerebrovascular disease, dementia, chronic pulmonary disease, Connective tissue disease, ulcer disease, mild liver disease, diabetes, hemiplegia, moderate or severe renal disease, diabetes with complications, any malignancy (including leukaemia and lymphoma), moderate or severe liver disease, metastatic solid tumour, and AIDS.  ^6^ IMD, Multiple Deprivation Index, quintile measured from patient-level address.  Reference group for variable sex is female, for age is 18-25, for BMI is healthy weight, for ethnicity is non-white, for CCI is very low, for smoking status is ex-smoker, for IMD is 2, for season is autumn, for region is east, for flu vaccination is no. | | | | | |

Supplementary Table 19. Adjusted hazard ratios of Cox models for hospital admissions related to upper respiratory tract infection (URTI), cough, cold with cough, and sore throat, using pre-pandemic data (from January 2019 to December 2019).

|  | **URTI, adjusted HR^1^ (95% CI^2^)** | | **Cough, adjusted HR (95% CI)** | | | | **Cough, adjusted HR (95% CI)** | | | | **Sore throat, adjusted HR (95% CI)** | | | |
| --- | --- | --- | --- | --- | --- | --- | --- | --- | --- | --- | --- | --- | --- | --- |
|  | **Incident** | | **Incident** | | **Prevalent** | | **Incident** | | **Prevalent** | | **Incident** | | **Prevalent** | |
|  | **No ABs^3^** | **With ABs** | **No ABs** | **With ABs** | **No ABs** | **With ABs** | **No ABs** | **With ABs** | **No ABs** | **With ABs** | **No ABs** | **With ABs** | **No ABs** | **With ABs** |
| **Sex** |  |  |  |  |  |  |  |  |  |  |  |  |  |  |
| Male | 1.30  (1.15-1.48) | 1.14  (1.05-1.24) | 1.12  (1.04-1.20) | 1.12  (1.05-1.20) | 1.13  (0.92-1.39) | 1.08  (0.88-1.34) | 1.12  (1.07-1.18) | 1.13  (1.09-1.17) | 1.17  (1.07-1.29) | 1.17  (1.08-1.26) | 1.50  (1.32-1.71) | 1.31  (1.19-1.44) | 1.33  (1.02-1.72) | 1.42  (1.08-1.88) |
| **Age** |  |  |  |  |  |  |  |  |  |  |  |  |  |  |
| 25-34 | 0.97  (0.7-1.34) | 0.89  (0.66-1.20) | 0.98  (0.73-1.34) | 1.13  (0.84-1.53) | 1.35  (0.53-3.43) | 2.18  (0.64-7.45) | 1.36  (1.09-1.68) | 1.12  (0.95-1.32) | 1.47  (0.87-2.49) | 0.85  (0.53-1.35) | 0.70  (0.59-0.84) | 0.75  (0.67-0.84) | 0.57  (0.42-0.78) | 0.6  (0.44-0.81) |
| 35-44 | 1.07  (0.78-1.49) | 0.95  (0.71-1.27) | 0.95  (0.70-1.28) | 1.06  (0.79-1.41) | 1.04  (0.42-2.61) | 1.10  (0.31-3.85) | 1.33  (1.07-1.64) | 1.05  (0.89-1.23) | 0.99  (0.58-1.69) | 0.69  (0.44-1.08) | 0.60  (0.49-0.74) | 0.63  (0.54-0.72) | 0.43  (0.29-0.65) | 0.29  (0.18-0.46) |
| 45-54 | 0.99  (0.71-1.37) | 1.02  (0.77-1.36) | 1.09  (0.83-1.45) | 1.19  (0.90-1.57) | 1.36  (0.57-3.24) | 1.74  (0.53-5.69) | 1.63  (1.33-2.00) | 1.17  (1.00-1.37) | 1.41  (0.86-2.31) | 1.04  (0.69-1.59) | 0.54  (0.43-0.68) | 0.62  (0.53-0.74) | 0.38  (0.23-0.61) | 0.47  (0.29-0.78) |
| 55-64 | 1.33  (0.97-1.83) | 1.30  (0.99-1.71) | 1.23  (0.94-1.61) | 1.36  (1.03-1.78) | 0.79  (0.33-1.90) | 1.16  (0.36-3.81) | 1.94  (1.59-2.36) | 1.45  (1.25-1.69) | 1.51  (0.93-2.45) | 1.12  (0.74-1.69) | 0.60  (0.47-0.77) | 0.60  (0.49-0.73) | 0.59  (0.35-0.97) | 0.33  (0.16-0.71) |
| 65-74 | 2.40  (1.75-3.29) | 1.63  (1.24-2.14) | 1.65  (1.26-2.15) | 1.81  (1.38-2.36) | 1.35  (0.57-3.17) | 1.97  (0.61-6.36) | 2.96  (2.44-3.60) | 1.99  (1.72-2.31) | 2.41  (1.49-3.90) | 1.44  (0.96-2.16) | 0.72  (0.55-0.95) | 0.66  (0.52-0.84) | 0.28  (0.12-0.67) | 0.66  (0.33-1.31) |
| 75+ | 4.52  (3.33-6.12) | 3.51  (2.69-4.57) | 3.38  (2.60-4.39) | 3.59  (2.76-4.68) | 2.18  (0.94-5.09) | 2.84  (0.89-9.11) | 6.48  (5.35-7.85) | 4.10  (3.55-4.74) | 3.96  (2.45-6.38) | 2.39  (1.60-3.59) | 1.10  (0.83-1.46) | 1.52  (1.23-1.89) | 0.33  (0.12-0.85) | 0.63  (0.27-1.47) |
| **BMI**^4^ |  |  |  |  |  |  |  |  |  |  |  |  |  |  |
| Underweight | 1.47  (1.02-2.11) | 1.79  (1.43-2.23) | 1.41  (1.15-1.72) | 1.41  (1.17-1.7) | 1.83  (1.06-3.17) | 1.59  (0.92-2.75) | 1.41  (1.24-1.59) | 1.44  (1.31-1.58) | 1.63  (1.28-2.06) | 1.37  (1.11-1.68) | 1.06  (0.68-1.66) | 1.17  (0.85-1.59) | 2.33  (1.23-4.44) | 1.15  (0.49-2.68) |
| Overweight | 0.94  (0.79-1.11) | 0.82  (0.73-0.92) | 0.82  (0.74-0.90) | 0.82  (0.75-0.9) | 0.84  (0.63-1.12) | 0.76  (0.57-1.01) | 0.84  (0.78-0.89) | 0.8  (0.77-0.84) | 0.83  (0.73-0.94) | 0.77  (0.7-0.86) | 0.84  (0.69-1.01) | 0.96  (0.84-1.10) | 1.26  (0.85-1.87) | 0.93  (0.62-1.38) |
| Obese | 0.91  (0.77-1.08) | 0.88  (0.79-0.99) | 0.86  (0.78-0.95) | 0.93  (0.85-1.01) | 0.96  (0.73-1.26) | 0.83  (0.63-1.09) | 0.9  (0.84-0.96) | 0.86  (0.82-0.9) | 0.91  (0.80-1.03) | 0.79  (0.71-0.87) | 1.10  (0.92-1.32) | 0.92  (0.81-1.05) | 1.19  (0.80-1.77) | 0.99  (0.68-1.44) |
| Unknown | 1.07  (0.89-1.29) | 1.05  (0.92-1.20) | 1.23  (1.08-1.40) | 1.07  (0.96-1.20) | 1.37  (0.98-1.92) | 1.28  (0.91-1.81) | 1.32  (1.22-1.42) | 1.06  (1.00-1.12) | 1.29  (1.11-1.49) | 0.97  (0.86-1.11) | 1.19  (1.00-1.41) | 0.92  (0.81-1.04) | 1.47  (1.03-2.10) | 1.10  (0.78-1.55) |
| **Ethnicity** |  |  |  |  |  |  |  |  |  |  |  |  |  |  |
| White | 1.02  (0.80-1.30) | 1.14  (0.97-1.35) | 0.89  (0.75-1.06) | 1.02  (0.88-1.18) | 1.04  (0.66-1.65) | 1.19  (0.71-2.00) | 0.98  (0.87-1.09) | 1.03  (0.95-1.11) | 1.04  (0.83-1.31) | 1.12  (0.92-1.38) | 1.09  (0.88-1.35) | 1.03  (0.87-1.22) | 0.9  (0.57-1.41) | 1.09  (0.64-1.86) |
| Unknown | 1.15  (0.89-1.48) | 1.22  (1.03-1.44) | 1.03  (0.86-1.23) | 1.17  (1.00-1.36) | 1.28  (0.80-2.05) | 1.24  (0.73-2.11) | 1.11  (0.99-1.25) | 1.13  (1.05-1.23) | 1.20  (0.95-1.51) | 1.25  (1.02-1.53) | 0.97  (0.78-1.21) | 1.13  (0.95-1.33) | 0.95  (0.60-1.52) | 1.34  (0.78-2.29) |
| **CCI**^5^ |  |  |  |  |  |  |  |  |  |  |  |  |  |  |
| Low | 1.59  (1.38-1.84) | 1.31  (1.19-1.45) | 1.50  (1.37-1.64) | 1.28  (1.19-1.39) | 1.71  (1.33-2.19) | 1.61  (1.25-2.08) | 1.42  (1.34-1.50) | 1.32  (1.26-1.37) | 1.48  (1.32-1.66) | 1.28  (1.16-1.41) | 1.40  (1.20-1.63) | 1.09  (0.97-1.22) | 1.52  (1.13-2.06) | 1.11  (0.80-1.54) |
| Medium | 2.01  (1.63-2.48) | 1.8  (1.57-2.06) | 2.15  (1.92-2.42) | 1.69  (1.52-1.88) | 2.60  (1.90-3.58) | 2.06  (1.48-2.86) | 1.92  (1.78-2.06) | 1.74  (1.65-1.84) | 2.09  (1.82-2.41) | 1.68  (1.5-1.89) | 1.89  (1.4-2.56) | 1.61  (1.27-2.06) | 0.87  (0.27-2.83) | 2.29  (1.14-4.59) |
| High | 3.53  (2.62-4.76) | 2.30  (1.85-2.86) | 3.06  (2.55-3.68) | 1.93  (1.61-2.32) | 2.49  (1.51-4.11) | 2.27  (1.36-3.80) | 2.46  (2.18-2.77) | 2.27  (2.08-2.47) | 2.13  (1.70-2.67) | 1.99  (1.64-2.40) | 1.21  (0.57-2.57) | 1.54  (0.93-2.55) | 3.58  (1.28-10.06) | 1.01  (0.14-7.42) |
| Very high | 3.20  (1.86-5.49) | 3.54  (2.57-4.88) | 4.01  (3.03-5.29) | 2.95  (2.28-3.81) | 1.27  (0.40-4.02) | 4.83  (2.32-10.04) | 3.41  (2.89-4.04) | 2.88  (2.52-3.28) | 3.03  (2.23-4.12) | 3.03  (2.32-3.95) | 3.97  (1.96-8.04) | 3.41  (1.87-6.22) | 4.78  (0.66-34.92) | 5.53  (1.29-23.58) |
| **Smoking** |  |  |  |  |  |  |  |  |  |  |  |  |  |  |
| Smoker | 1.16  (0.96-1.41) | 0.94  (0.82-1.07) | 0.93  (0.83-1.04) | 0.89  (0.81-0.99) | 0.69  (0.48-0.98) | 0.56  (0.39-0.80) | 0.95  (0.89-1.02) | 0.88  (0.84-0.93) | 0.87  (0.75-1.02) | 0.87  (0.77-0.99) | 1.47  (1.24-1.75) | 1.17  (1.04-1.32) | 1.06  (0.76-1.48) | 0.88  (0.62-1.26) |
| Never smoked | 0.88  (0.77-1.01) | 0.93  (0.85-1.03) | 1.17  (1.06-1.28) | 1.01  (0.94-1.09) | 0.95  (0.75-1.19) | 0.98  (0.77-1.24) | 1.11  (1.05-1.17) | 1.00  (0.96-1.04) | 0.94  (0.85-1.05) | 1.00  (0.91-1.09) | 0.95  (0.82-1.10) | 0.89  (0.80-0.98) | 0.79  (0.59-1.05) | 0.82  (0.61-1.09) |
| Unknown | 0.33  (0.08-1.32) | 0.52  (0.19-1.40) | 0.58  (0.19-1.83) | 1.14  (0.56-2.30) | 1.56  (0.21-11.32) | 1.84  (0.25-13.43) | 1.33  (0.86-2.06) | 1.07  (0.75-1.53) | 1.10  (0.41-2.95) | 0.72  (0.23-2.24) | 1.03  (0.67-1.56) | 0.84  (0.63-1.12) | 1.06  (0.55-2.03) | 0.66  (0.32-1.37) |
| **IMD**^6^ |  |  |  |  |  |  |  |  |  |  |  |  |  |  |
| 1 (most deprived) | 1.02  (0.84-1.24) | 1.04  (0.92-1.18) | 0.98  (0.88-1.09) | 1.09  (0.99-1.21) | 1.07  (0.79-1.45) | 0.95  (0.70-1.30) | 1.10  (1.02-1.18) | 1.05  (1.00-1.11) | 1.12  (0.98-1.29) | 1.09  (0.97-1.22) | 0.93  (0.77-1.12) | 1.08  (0.94-1.24) | 1.41  (0.96-2.07) | 1.08  (0.73-1.6) |
| 3 | 0.88  (0.72-1.06) | 0.95  (0.84-1.08) | 0.82  (0.73-0.92) | 0.96  (0.87-1.07) | 0.78  (0.57-1.07) | 0.86  (0.63-1.17) | 0.88  (0.81-0.94) | 0.99  (0.94-1.05) | 0.87  (0.76-1.01) | 1.00  (0.89-1.13) | 0.96  (0.79-1.16) | 1.11  (0.97-1.27) | 1.21  (0.82-1.78) | 1.05  (0.72-1.54) |
| 4 | 1.00  (0.83-1.2) | 0.96  (0.85-1.10) | 0.81  (0.72-0.91) | 1.00  (0.90-1.11) | 0.81  (0.58-1.11) | 0.94  (0.68-1.29) | 0.94  (0.87-1.02) | 1.00  (0.95-1.06) | 0.93  (0.81-1.08) | 0.93  (0.82-1.05) | 0.94  (0.77-1.15) | 1.06  (0.92-1.22) | 1.14  (0.76-1.71) | 0.91  (0.61-1.36) |
| 5 (most affluent) | 0.85  (0.7-1.05) | 0.92  (0.8-1.06) | 0.70  (0.62-0.80) | 0.88  (0.79-0.99) | 0.80  (0.57-1.11) | 0.75  (0.52-1.08) | 0.83  (0.77-0.90) | 0.89  (0.84-0.94) | 0.79  (0.68-0.93) | 1.01  (0.89-1.14) | 1.04  (0.85-1.28) | 1.11  (0.96-1.29) | 1.10  (0.73-1.67) | 1.19  (0.80-1.75) |
| Unknown | 1.13  (0.69-1.86) | 0.70  (0.48-1.03) | 0.77  (0.56-1.07) | 1.04  (0.79-1.35) | 0.89  (0.38-2.04) | 1.06  (0.51-2.21) | 1.15  (0.96-1.39) | 0.92  (0.79-1.06) | 1.04  (0.72-1.51) | 1.13  (0.84-1.51) | 1.39  (0.91-2.13) | 1.18  (0.83-1.67) | 1.16  (0.46-2.94) | 1.20  (0.48-3.03) |
| **Season** |  |  |  |  |  |  |  |  |  |  |  |  |  |  |
| Spring | 1.05  (0.88-1.24) | 0.88  (0.78-1.00) | 1.00  (0.90-1.11) | 1.00  (0.91-1.1) | 0.72  (0.54-0.96) | 0.90  (0.68-1.19) | 0.96  (0.9-1.03) | 0.95  (0.90-1.00) | 0.83  (0.73-0.95) | 0.93  (0.83-1.04) | 1.05  (0.88-1.25) | 0.97  (0.86-1.09) | 1.27  (0.91-1.78) | 0.78  (0.54-1.12) |
| Summer | 0.92  (0.75-1.13) | 0.95  (0.83-1.08) | 0.82  (0.73-0.92) | 0.91  (0.82-1.01) | 0.77  (0.57-1.04) | 0.80  (0.57-1.10) | 0.98  (0.91-1.05) | 0.94  (0.89-0.99) | 0.92  (0.8-1.06) | 0.91  (0.80-1.03) | 1.22  (1.02-1.46) | 0.92  (0.81-1.04) | 1.19  (0.83-1.7) | 1.05  (0.74-1.5) |
| Winter | 0.99  (0.85-1.15) | 1.02  (0.92-1.13) | 1.11  (1.01-1.22) | 0.97  (0.89-1.05) | 0.93  (0.72-1.20) | 0.81  (0.62-1.06) | 1.12  (1.05-1.19) | 0.93  (0.89-0.98) | 0.98  (0.87-1.10) | 0.97  (0.88-1.07) | 1.05  (0.89-1.25) | 0.86  (0.76-0.97) | 1.12  (0.79-1.58) | 1.06  (0.76-1.48) |
| **Region** |  |  |  |  |  |  |  |  |  |  |  |  |  |  |
| London | 0.86  (0.61-1.21) | 0.76  (0.61-0.94) | 0.90  (0.7-1.16) | 0.98  (0.80-1.19) | 1.40  (0.75-2.62) | 0.77  (0.33-1.79) | 0.96  (0.82-1.13) | 1.00  (0.9-1.12) | 1.14  (0.84-1.55) | 0.96  (0.70-1.31) | 1.14  (0.83-1.56) | 1.12  (0.89-1.39) | 1.12  (0.59-2.13) | 0.85  (0.38-1.88) |
| North East | 1.03  (0.75-1.42) | 1.00  (0.81-1.24) | 0.88  (0.75-1.03) | 0.97  (0.83-1.13) | 1.00  (0.64-1.56) | 1.10  (0.70-1.73) | 0.88  (0.79-0.98) | 0.95  (0.88-1.04) | 0.93  (0.76-1.14) | 1.05  (0.88-1.25) | 1.22  (0.92-1.63) | 1.03  (0.82-1.28) | 1.16  (0.67-2.00) | 0.92  (0.48-1.74) |
| North West | 0.84  (0.66-1.07) | 1.09  (0.93-1.27) | 0.74  (0.65-0.85) | 1.08  (0.96-1.22) | 0.82  (0.57-1.18) | 1.17  (0.82-1.65) | 0.78  (0.72-0.85) | 1.00  (0.94-1.07) | 0.82  (0.70-0.97) | 0.95  (0.83-1.09) | 1.07  (0.84-1.34) | 1.29  (1.10-1.51) | 0.96  (0.60-1.53) | 0.84  (0.52-1.36) |
| West Midlands | 1.21  (0.88-1.65) | 1.06  (0.88-1.28) | 0.97  (0.80-1.18) | 1.25  (1.07-1.46) | 1.05  (0.62-1.79) | 1.52  (0.92-2.52) | 1.10  (0.97-1.23) | 1.09  (1.00-1.18) | 1.13  (0.89-1.43) | 1.10  (0.90-1.34) | 1.21  (0.9-1.62) | 1.26  (1.02-1.56) | 1.00  (0.55-1.81) | 1.47  (0.88-2.48) |
| Yorkshire and The Humber | 1.25  (1.02-1.52) | 1.01  (0.89-1.15) | 0.84  (0.74-0.95) | 1.03  (0.92-1.15) | 1.05  (0.76-1.45) | 0.91  (0.64-1.29) | 0.93  (0.87-1.01) | 0.98  (0.92-1.03) | 0.98  (0.85-1.13) | 0.99  (0.87-1.11) | 1.19  (0.97-1.45) | 1.03  (0.89-1.19) | 0.90  (0.60-1.36) | 0.99  (0.66-1.47) |
| South East | 1.20  (0.94-1.54) | 1.05  (0.87-1.28) | 0.74  (0.63-0.88) | 0.86  (0.74-1.01) | 1.11  (0.73-1.67) | 0.88  (0.55-1.42) | 0.90  (0.81-0.99) | 0.93  (0.86-1.01) | 0.83  (0.68-1.03) | 1.00  (0.84-1.19) | 1.34  (1.04-1.74) | 1.39  (1.15-1.67) | 0.85  (0.49-1.47) | 1.46  (0.93-2.31) |
| East Midlands | 1.24  (1.03-1.49) | 1.05  (0.92-1.19) | 0.94  (0.84-1.06) | 1.22  (1.1-1.35) | 1.24  (0.91-1.68) | 1.25  (0.91-1.71) | 1.09  (1.01-1.17) | 1.09  (1.04-1.15) | 1.12  (0.97-1.28) | 1.12  (1.00-1.26) | 1.16  (0.95-1.40) | 1.03  (0.90-1.19) | 1.12  (0.78-1.62) | 0.86  (0.59-1.27) |
| South West | 0.95  (0.75-1.19) | 1.07  (0.91-1.27) | 0.68  (0.59-0.78) | 1.05  (0.93-1.18) | 0.65  (0.42-1.00) | 1.12  (0.78-1.62) | 0.8  (0.73-0.87) | 0.98  (0.92-1.05) | 0.82  (0.68-0.98) | 1.00  (0.86-1.16) | 1.23  (0.99-1.53) | 1.46  (1.25-1.69) | 1.25  (0.83-1.86) | 1.25  (0.82-1.90) |
| **Flu vaccination** |  |  |  |  |  |  |  |  |  |  |  |  |  |  |
| Yes | 0.84  (0.73-0.98) | 1.00  (0.91-1.11) | 0.87  (0.79-0.95) | 0.98  (0.90-1.06) | 1.00  (0.78-1.28) | 1.05  (0.82-1.35) | 0.81  (0.76-0.85) | 0.92  (0.88-0.96) | 0.89  (0.80-0.99) | 0.99  (0.90-1.08) | 0.87  (0.73-1.04) | 1.10  (0.97-1.26) | 0.80  (0.54-1.21) | 1.22  (0.84-1.77) |
| **Count of antibiotic prescription in the one year before** | 1.20  (1.17-1.23) | 1.17  (1.15-1.19) | 1.18  (1.16-1.19) | 1.14  (1.12-1.15) | 1.20  (1.16-1.25) | 1.13  (1.09-1.18) | 1.12  (1.12-1.13) | 1.11  (1.11-1.12) | 1.11  (1.09-1.13) | 1.09  (1.07-1.1) | 1.19  (1.14-1.24) | 1.17  (1.14-1.21) | 1.02  (0.94-1.11) | 1.20  (1.10-1.31) |
| ^1^ HR, hazard ratio.  ^2^ CI, confidence interval.  ^3^ ABs, antibiotics prescribed or not.  ^4^ BMI, Body Mass Index recorded in the last 5 years.  ^5^ CCI, Charlson Comorbidities Index, measured from 17 weighted conditions, including myocardial infarction, congestive heart failure, peripheral vascular disease, cerebrovascular disease, dementia, chronic pulmonary disease, Connective tissue disease, ulcer disease, mild liver disease, diabetes, hemiplegia, moderate or severe renal disease, diabetes with complications, any malignancy (including leukaemia and lymphoma), moderate or severe liver disease, metastatic solid tumour, and AIDS.  ^6^ IMD, Multiple Deprivation Index, quintile measured from patient-level address.  Reference group for variable sex is female, for age is 18-25, for BMI is healthy weight, for ethnicity is non-white, for CCI is very low, for smoking status is ex-smoker, for IMD is 2, for season is autumn, for region is east, for flu vaccination is no. | | | | | | | | | | | | | | |

#### Calibration plots

| A  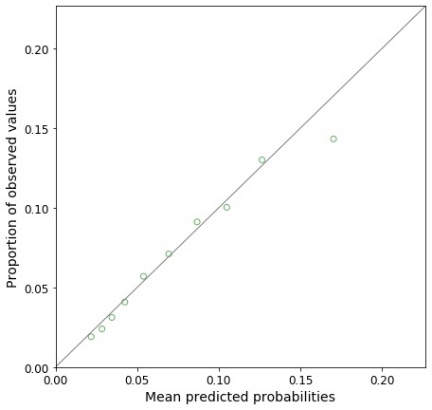 | B  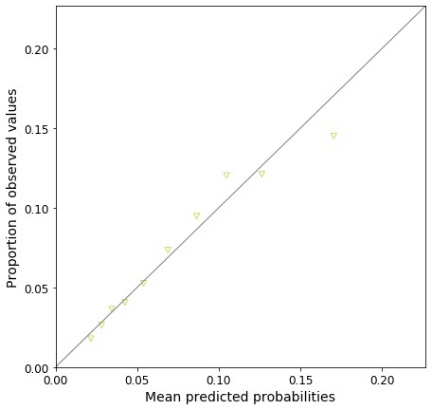 |
| --- | --- |
| C  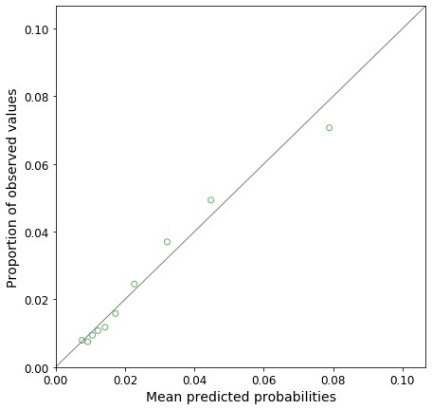 | D  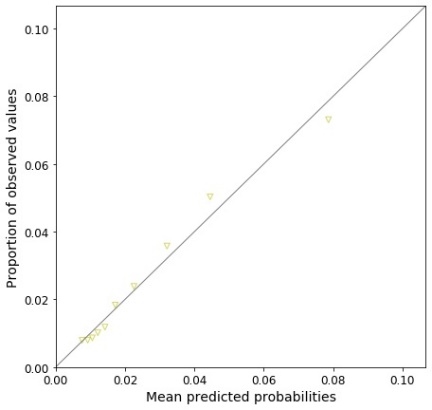 |
| E  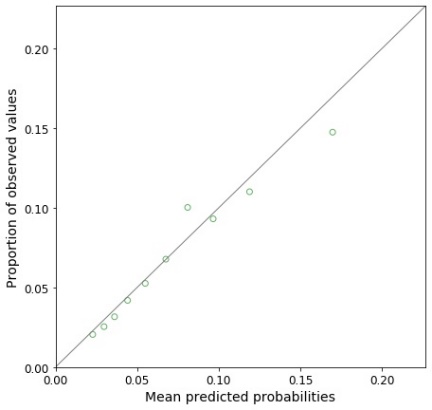 | F  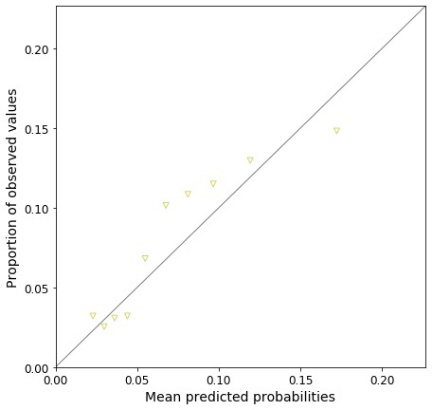 |
| G  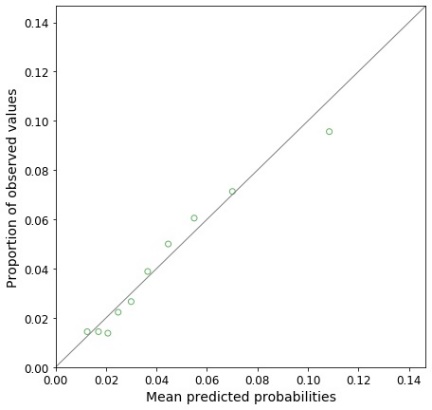 | H  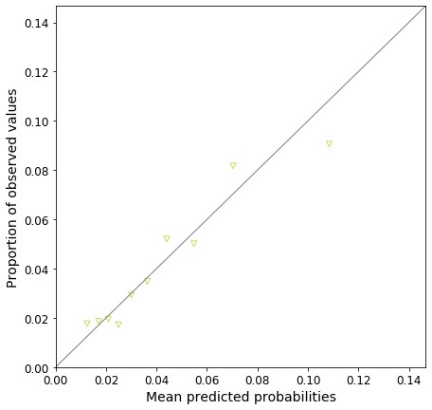 |
| *Supplementary Figure 1. Calibration plots of Cox models for infection-related hospital admission following a lower respiratory tract infection (LRTI), developed and validated with pre-pandemic data (from January 2019 to December 2019): (A) incident LRTI with no antibiotics using development dataset, (B) incident LRTI with no antibiotics using validation dataset, (C) incident LRTI with antibiotics using development dataset, (D) incident LRTI with antibiotics using validation dataset, (E) prevalent LRTI with no antibiotics using development dataset, (F) prevalent LRTI with no antibiotics using validation dataset, (G) prevalent LRTI with antibiotics using development dataset, (H) prevalent LRTI with antibiotics using validation dataset.* | |

| A  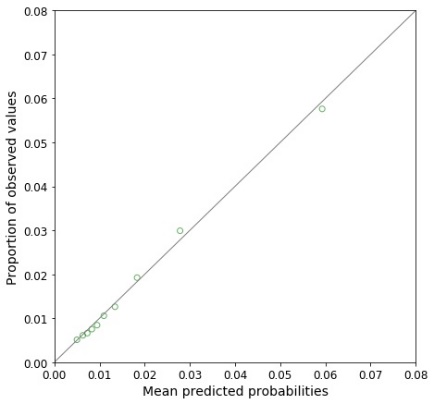 | B  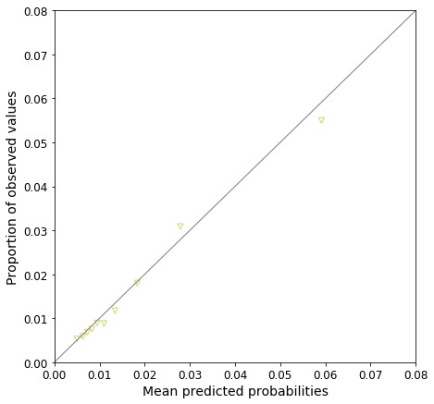 |
| --- | --- |
| C  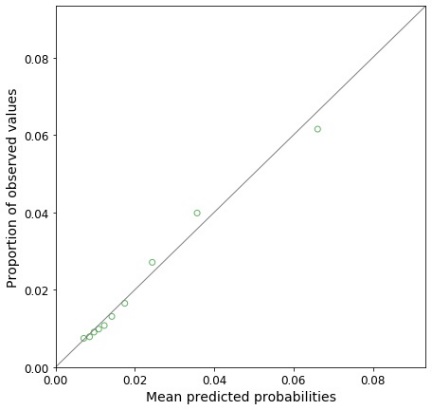 | D  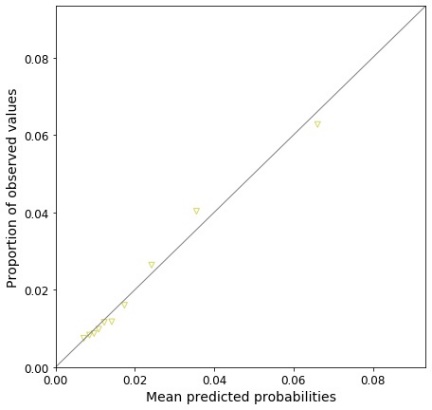 |
| E  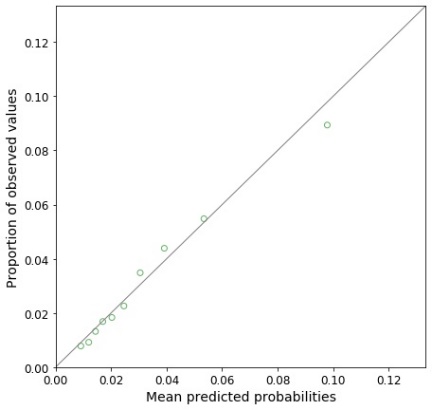 | F  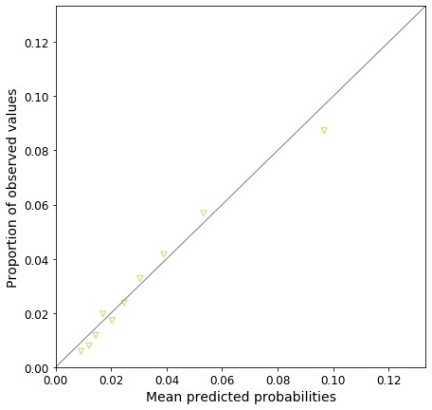 |
| G  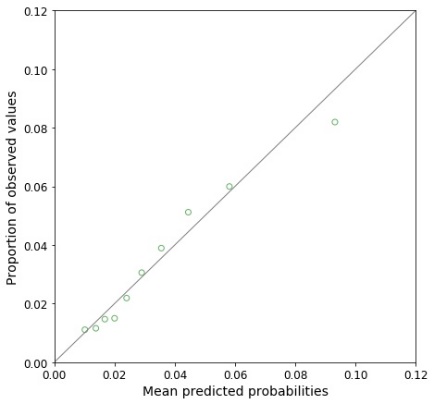 | H  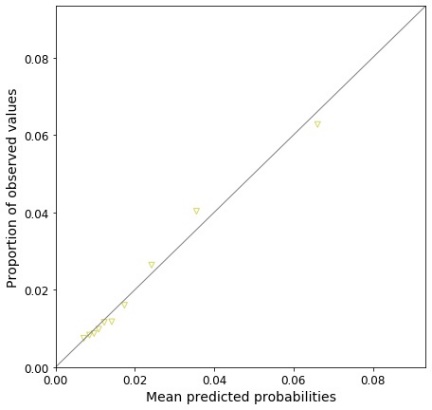 |
| Supplementary Figure 2. *Calibration plots of Cox models for infection-related hospital admission following* upper respiratory tract infection (URTI; including URTI, cough, cold with cough, and sore throat*), developed and validated with pre-pandemic data (from January 2019 to December 2019): (A) incident URTI with no antibiotics using development dataset, (B) incident URTI with no antibiotics using validation dataset, (C) incident URTI with antibiotics using development dataset, (D) incident URTI with antibiotics using validation dataset, (E) prevalent URTI with no antibiotics using development dataset, (F) prevalent URTI with no antibiotics using validation dataset, (G) prevalent URTI with antibiotics using development dataset, (H) prevalent URTI with antibiotics using validation dataset.* | |

| A  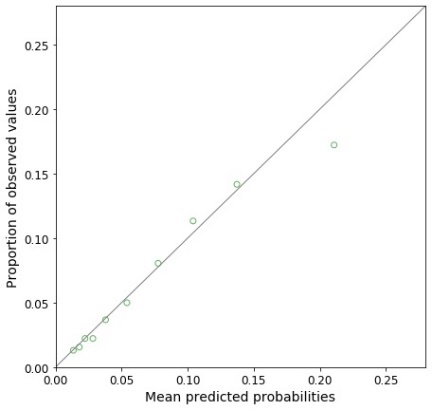 | B  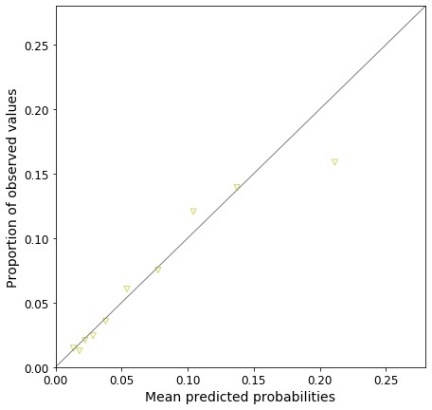 |
| --- | --- |
| C  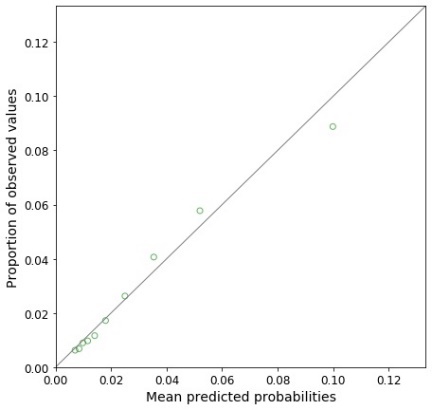 | D  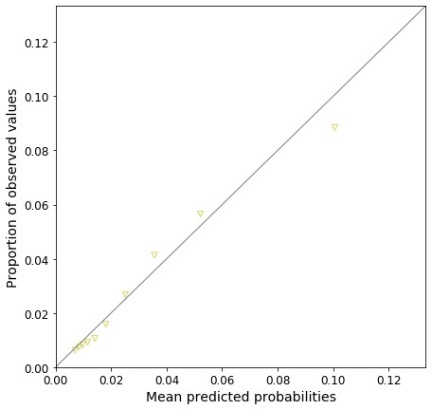 |
| E  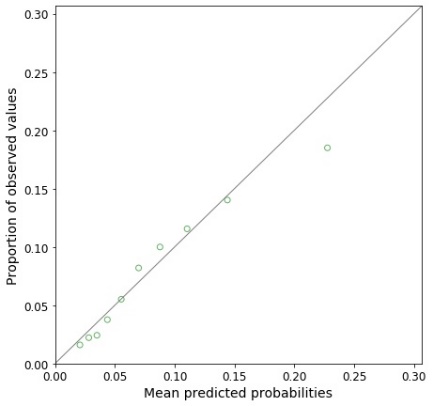 | F  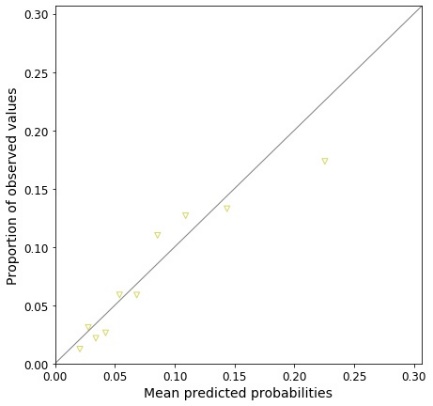 |
| G  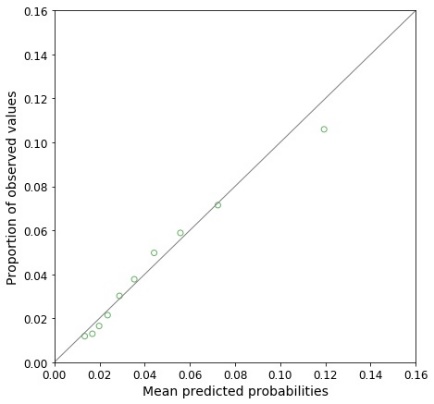 | H  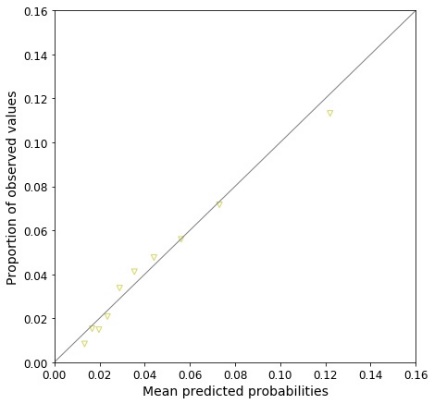 |
| *Supplementary Figure 3. Calibration plots of Cox models for infection-related hospital admission following a urinary tract infection (UTI) , developed and validated with pre-pandemic data (from January 2019 to December 2019): (A) incident UTI with no antibiotics using development dataset, (B) incident UTI with no antibiotics using validation dataset, (C) incident UTI with antibiotics using development dataset, (D) incident UTI with antibiotics using validation dataset, (E) prevalent UTI with no antibiotics using development dataset, (F) prevalent UTI with no antibiotics using validation dataset, (G) prevalent UTI with antibiotics using development dataset, (H) prevalent UTI with antibiotics using validation dataset.* | |

| A  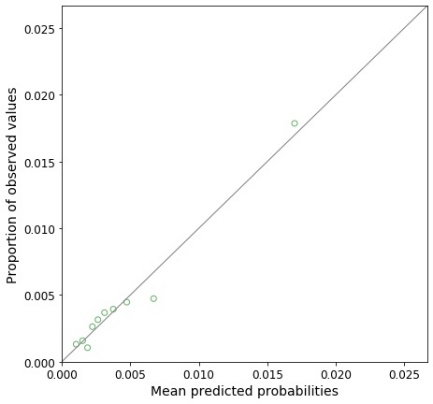 | B  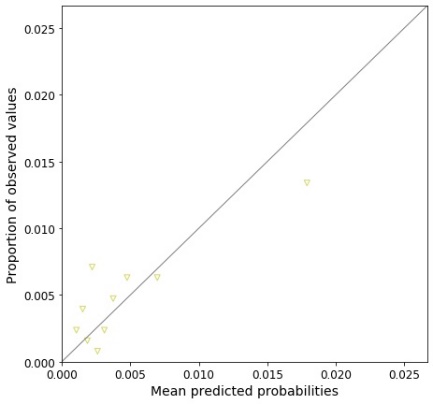 |
| --- | --- |
| *Supplementary Figure 4. Calibration plots of Cox models for infection-related hospital admission following a sinusitis, developed and validated with pre-pandemic data (from January 2019 to December 2019): (A) incident sinusitis with no antibiotics using development dataset, (B) incident sinusitis with no antibiotics using validation dataset.* | |

| A  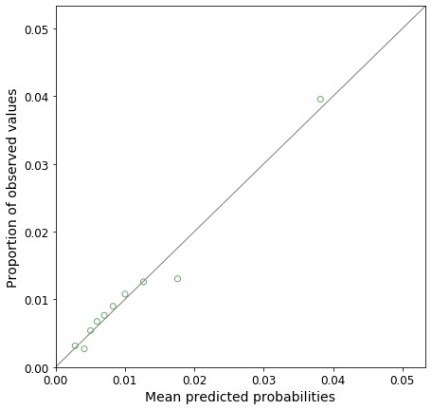 | B  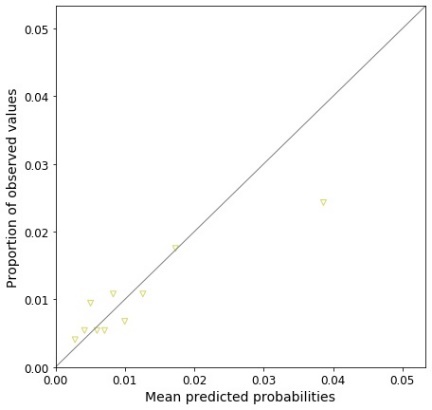 |
| --- | --- |
| C  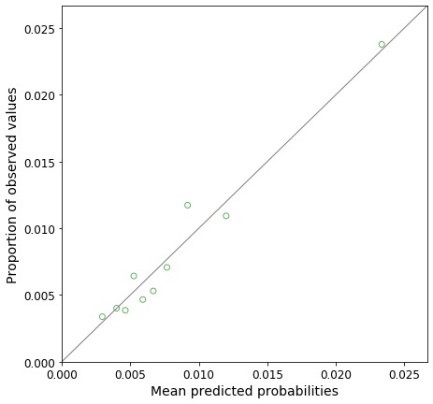 | D  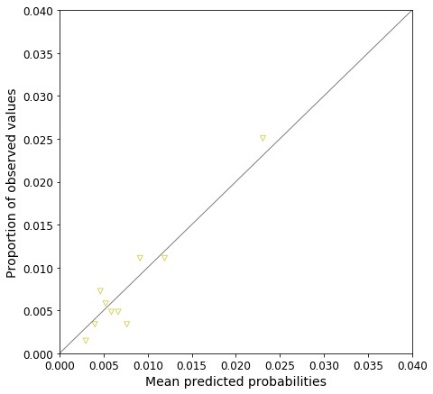 |
| *Supplementary Figure 5. Calibration plots of Cox models for infection-related hospital admission following an otitis media, developed and validated with pre-pandemic data (from January 2019 to December 2019): (A) incident otitis media with no antibiotics using development dataset, (B) incident otitis media with no antibiotics using validation dataset, (C) incident otitis media with antibiotics using development dataset, (D) incident otitis media with antibiotics using validation dataset.* | |

| A  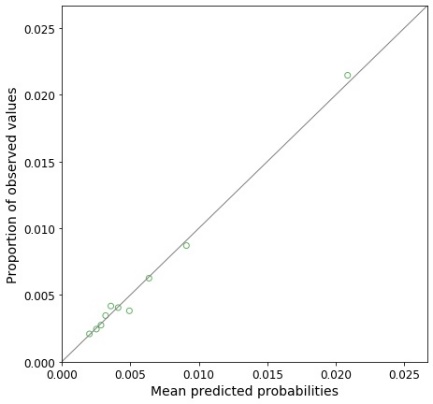 | B  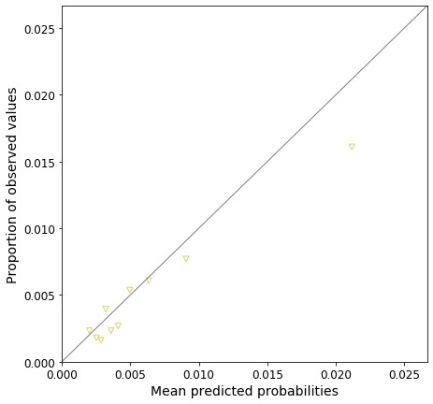 |
| --- | --- |
| C  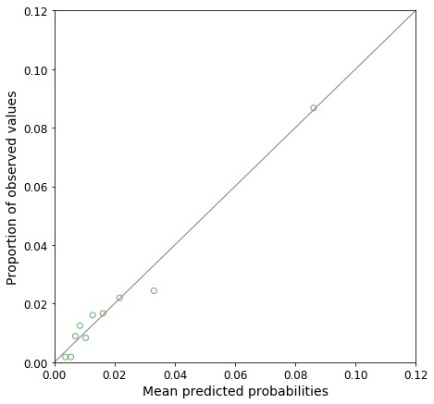 | D  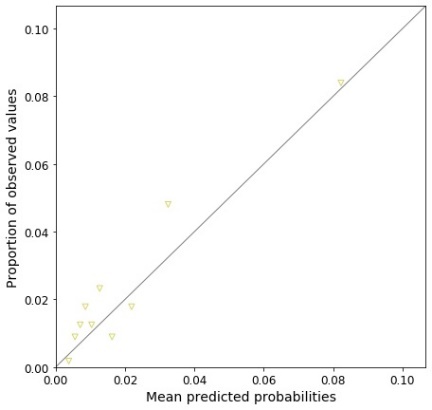 |
| *Supplementary Figure 6. Calibration plots of Cox models for infection-related hospital admission following an otitis externa, developed and validated with pre-pandemic data (from January 2019 to December 2019): (A) incident otitis externa with no antibiotics using development dataset, (B) incident otitis externa with no antibiotics using validation dataset, (C) prevalent otitis externa with no antibiotics using development dataset, (D) prevalent otitis externa with no antibiotics using validation dataset.* | |

| A  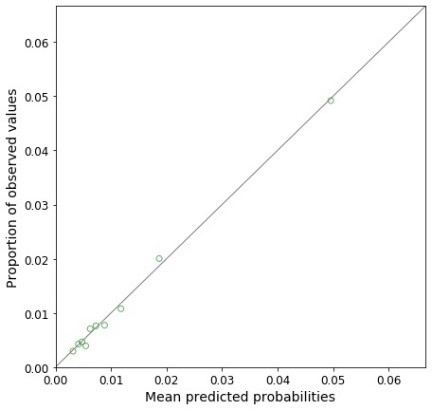 | B  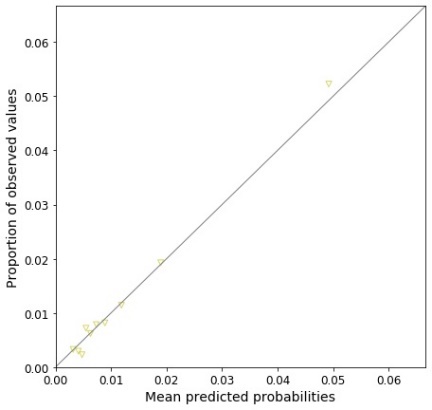 |
| --- | --- |
| C  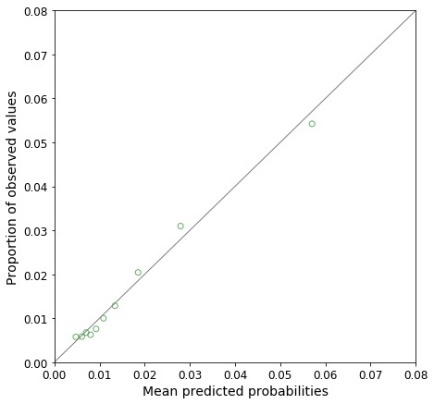 | D  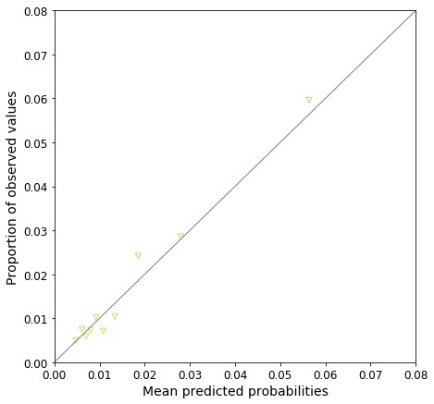 |
| *Supplementary Figure 7. Calibration plots of Cox models for infection-related hospital admission following an upper respiratory tract infection (URTI), developed and validated with pre-pandemic data (from January 2019 to December 2019): (A) incident URTI with no antibiotics using development dataset, (B) incident URTI with no antibiotics using validation dataset, (C) incident URTI with antibiotics using development dataset, (D) incident URTI with antibiotics using validation dataset.* | |

| A  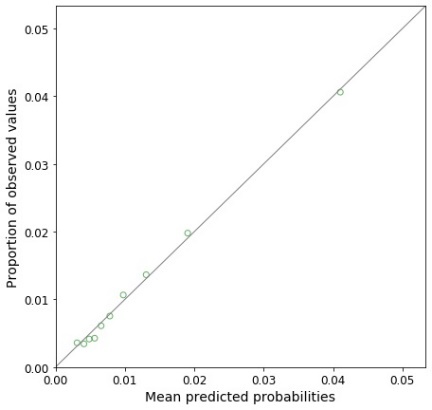 | B  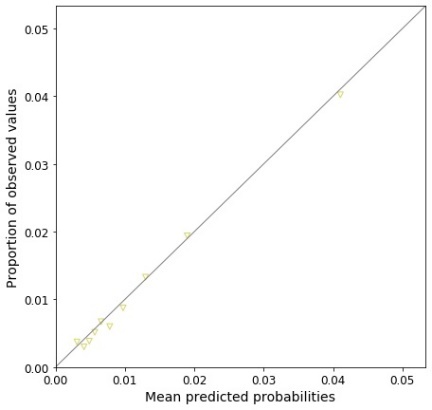 |
| --- | --- |
| C  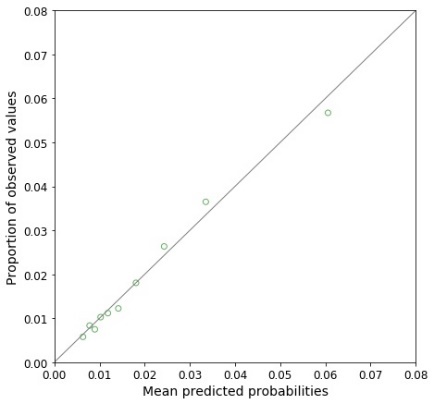 | D  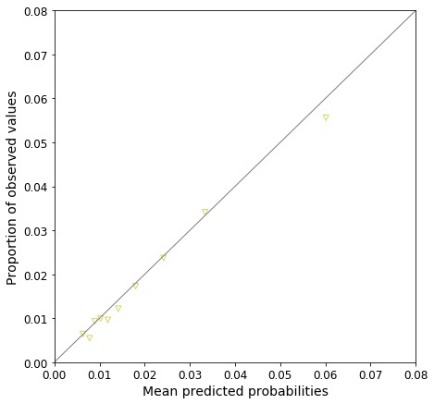 |
| E  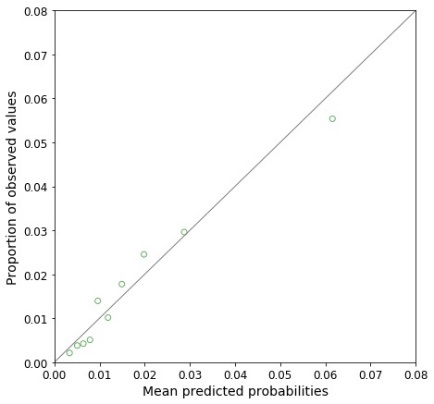 | F  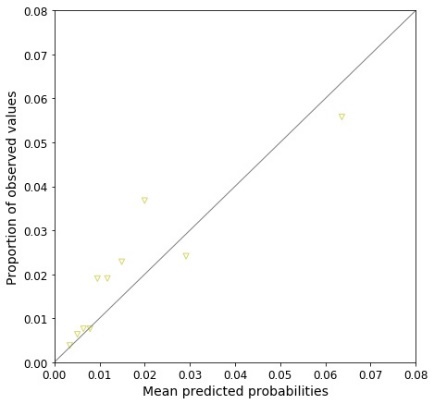 |
| G  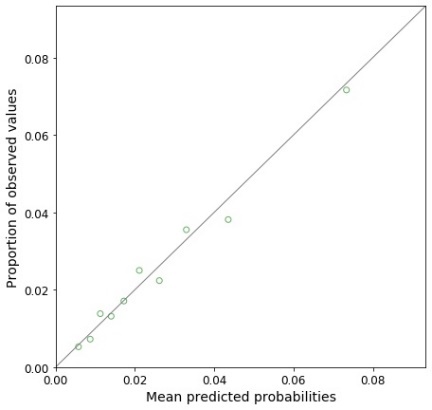 | H  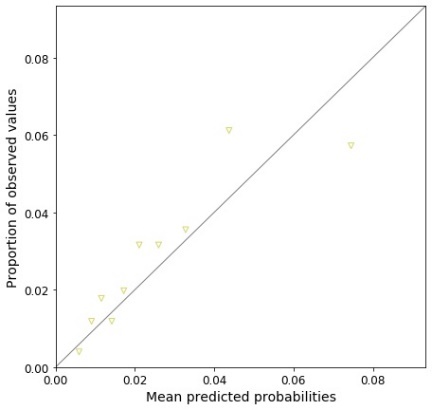 |
| *Supplementary Figure 8. Calibration plots of Cox models for infection-related hospital admission following a cough, developed and validated with pre-pandemic data (from January 2019 to December 2019): (A) incident cough with no antibiotics using development dataset, (B) incident cough with no antibiotics using validation dataset, (C) incident cough with antibiotics using development dataset, (D) incident cough with antibiotics using validation dataset, (E) prevalent cough with no antibiotics using development dataset, (F) prevalent cough with no antibiotics using validation dataset, (G) prevalent cough with antibiotics using development dataset, (H) prevalent cough with antibiotics using validation dataset.* | |

| A  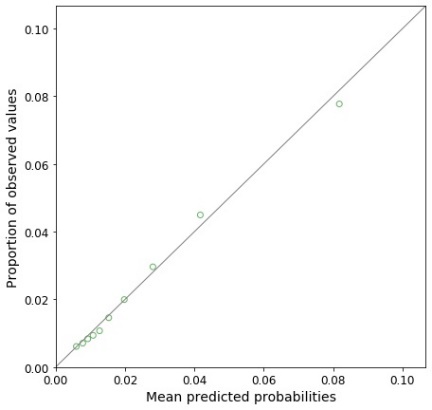 | B  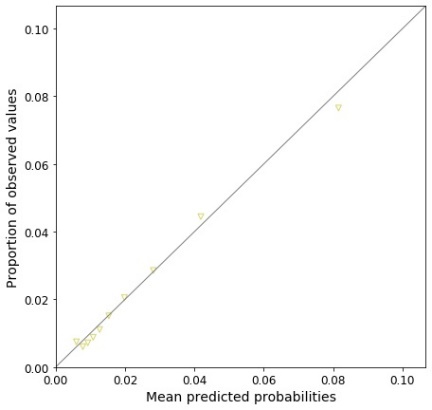 |
| --- | --- |
| C  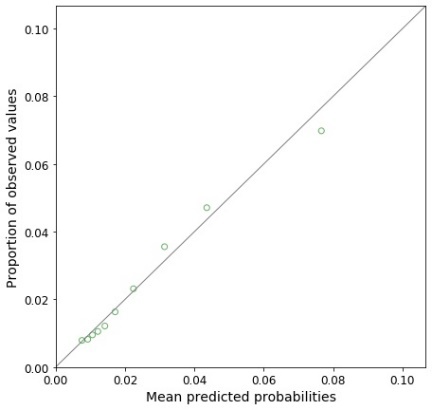 | D  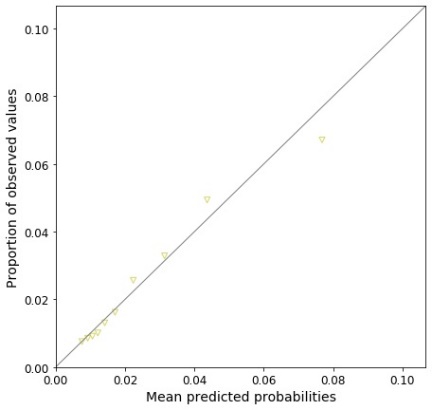 |
| E  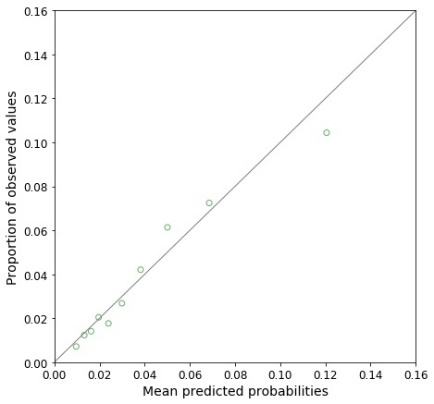 | F  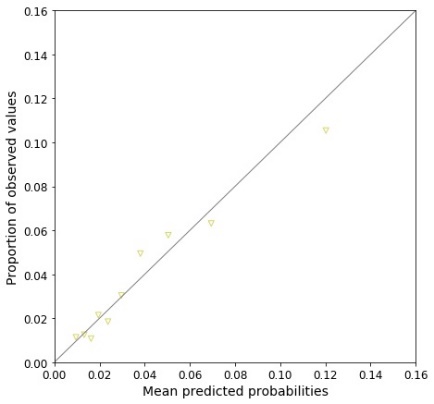 |
| G  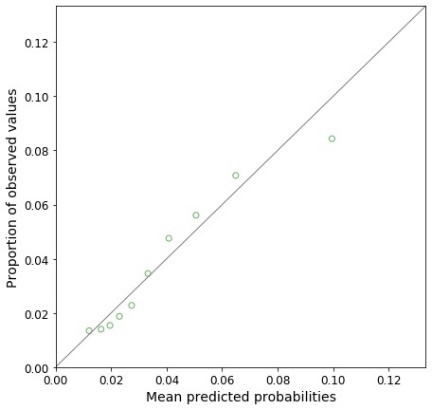 | H  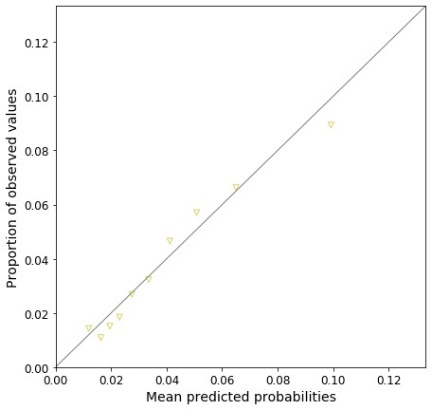 |
| *Supplementary Figure 9. Calibration plots of Cox models for infection-related hospital admission following a cold with cough, developed and validated with pre-pandemic data (from January 2019 to December 2019): (A) incident cold with cough without antibiotics using development dataset, (B) incident cold with cough without antibiotics using validation dataset, (C) incident cold with cough with antibiotics using development dataset, (D) incident cold with cough with antibiotics using validation dataset, (E) prevalent cold with cough without antibiotics using development dataset, (F) prevalent cold with cough without antibiotics using validation dataset, (G) prevalent cold with cough with antibiotics using development dataset, (H) prevalent cold with cough with antibiotics using validation dataset.* | |

| A  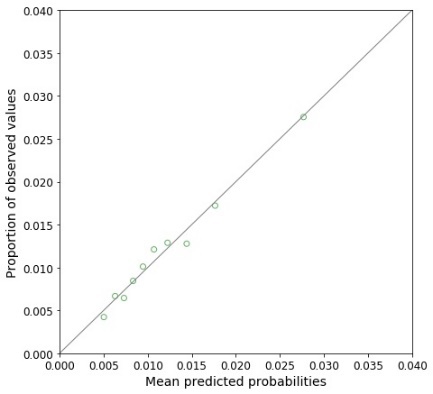 | B  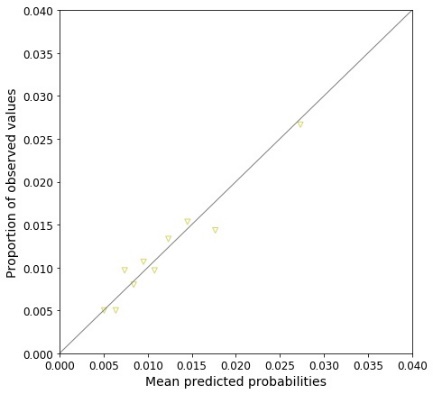 |
| --- | --- |
| C  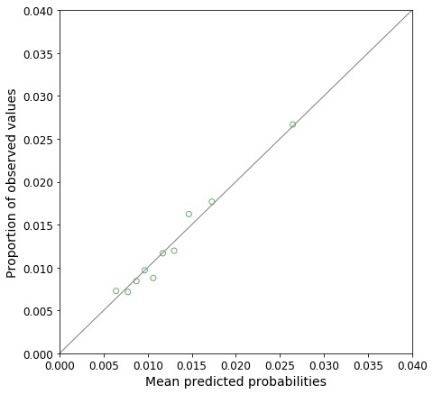 | D  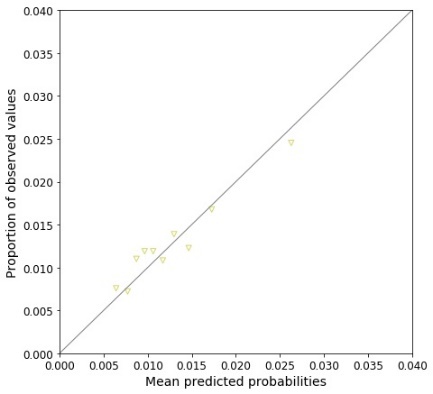 |
| E  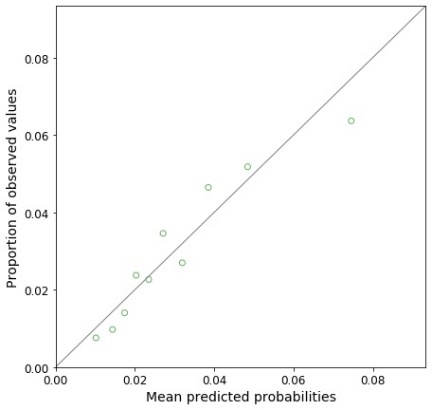 | F  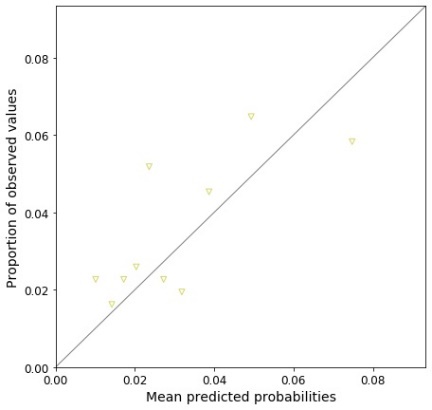 |
| G  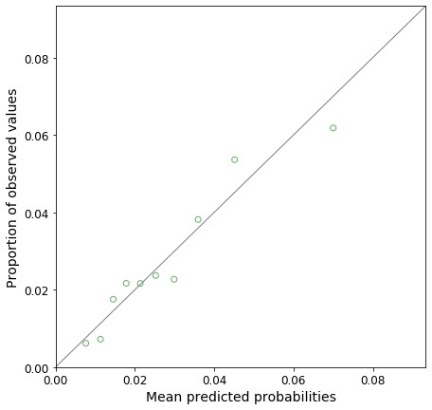 | H  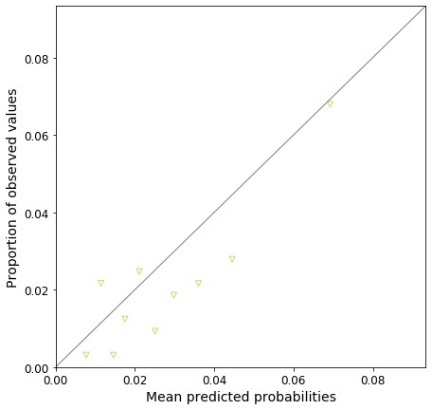 |
| *Supplementary Figure 10. Calibration plots of Cox models for infection-related hospital admission following a sore throat, developed and validated with pre-pandemic data (from January 2019 to December 2019): (A) incident sore throat with no antibiotics using development dataset, (B) incident sore throat with no antibiotics using validation dataset, (C) incident sore throat with antibiotics using development dataset, (D) incident sore throat with antibiotics using validation dataset, (E) prevalent sore throat with no antibiotics using development dataset, (F) prevalent sore throat with no antibiotics using validation dataset, (G) prevalent sore throat with antibiotics using development dataset, (H) prevalent sore throat with antibiotics using validation dataset.* | |
